# Transmission acceleration outperforms the endemic-channel threshold for dengue outbreak detection

**DOI:** 10.64898/2026.05.15.26353307

**Authors:** Keanu John Pelitro, Julia Fye Manzano, Troy Owen Matavia, Kylone Soriano, Klara Bilbao, Gereka Marie Garcia, Aira Joy Delos Angeles, Alfredo Mahar Lagmay, DJ Darwin Bandoy

**Affiliations:** University of the Philippines Resilience Institute, Quezon City, Philippines; College of Veterinary Medicine, University of the Philippines, Los Baños, Laguna, Philippines; National Institute of Geological Sciences, University of the Philippines Diliman, Quezon City, Philippines

## Abstract

Extreme weather drives dengue transmission beyond historical spatial and temporal incidence patterns. No operational dengue outbreak threshold has been validated for early warning under extreme weather conditions. Here we show that transmission acceleration provides earlier and more consistent outbreak detection than the endemic-channel threshold. We evaluated detectors across a hyperendemic city, seventeen tropical regions, and eight dengue-endemic countries. Acceleration-based detectors consistently outperformed the endemic channel in burden captured, lead time, warning persistence, and sensitivity. At city scale, transmission acceleration captured 4,257 cases per season versus 1,119. It achieved 100% sensitivity versus 30% while triggering within the four-to-eight-week anticipatory window under 6.9 versus 1.6 weeks lead time. These establish acceleration-based detectors as a robust signal for dengue anticipatory action, making them better suited to a transmission landscape increasingly shaped by extreme weather conditions.

## Introduction

Timely outbreak detection is foundational to public health, yet routine dengue surveillance remains constrained by the limitations of current threshold-based approaches.^1–3^ One of the most widely operationalized approach is the endemic channel, endorsed in the WHO 2009 guidelines and typically implemented as a rolling mean plus two standard deviations.^3–8^ Constructed from historical case counts, this fixed upper limit defines expected seasonal transmission and triggers an alert only when the current incidence exceeds it. Its relative simplicity accommodates limited regional epidemiological technical capacity. ^9,10^ However, historical baselines cannot adjust to real time outbreak dynamics driven by ecological and biological variables like serotype rotation and climate-sensitive transmission.^11–14^ Furthermore, calibration with high case incidence considered non-epidemic years or seasonal transmission in the calculation inflates the mean and standard deviation, raising the outbreak threshold above the level of inter-epidemic seasons.^7,14^ Warnings are therefore operationally delayed, allowing cases to accumulate in the interval between detection and response.

The continued reliance on the endemic channel also underscores other methodological limitations. Intensifying weather patterns are modifying inter-epidemic periods and amplifying transmission peaks, pushing dengue dynamics beyond historically observed ranges.^15,16^ Consequently, in climate-vulnerable regions, the predictive accuracy of early warning signals diminishes when thresholds are calibrated solely on historical incidence.^17,18^ Originally adapted from the surveillance process for low-incidence diseases like malaria, endemic channel was adapted to dengue without recalibration for transmission intensity or serotype dynamics.^5,6^ Consequently, this created a self-referential evaluation loop. The same threshold that defines an outbreak also serves as its own reference standard, which restricts independent assessment of detection timeliness, sensitivity, or intervention impact. ^8,10^ Without an external validation framework, routine surveillance cannot systematically evaluate how definitional choices affect early-warning performance or outbreak trajectories. To address these limitations, we formulated an outbreak detection evaluation framework based on anticipatory metrics, separating how detection thresholds are constructed from how their performance is assessed (Methods). The evaluation separates early outbreak signalling from definitive outbreak classification and assesses whether the signal arrives early enough to support anticipatory action.

This reframing fits a wider move toward rate-of-change indicators, which prioritize early signals over matching past patterns. The need for this approach is clear when historical baselines are missing or distorted, as shown during the COVID-19 pandemic. ^19,20^ Although different from dengue, COVID-19 showed that surveillance must assess risk from real-time transmission rather than fixed magnitude, using measures of epidemic acceleration instead of absolute case counts. Using rate of change to detect the beginning of an increase is well established across fields, ^21,22^ from early-warning signals in ecology to acceleration-based methods used for respiratory pathogen surveillance. ^23,24^ Applying this to dengue, we measure the onset of epidemic growth as the ratio of short-term to long-term incidence, a calculation adapted from detecting noisy seismic waves.^25,26^ Unlike thresholds based on case numbers, acceleration identifies the point where case transmission begin to significantly rise.^21,23,25^

As a consequence, the effectiveness of dengue outbreak detectors has not been independently evaluated because existing assessments lack a validation framework separate from the definition itself.^7–9^ We therefore defined performance around the purpose of surveillance: to warn before the burden has already accumulated. This required metrics that capture timeliness, morbidity coverage, signal duration, and unnecessary alerts.We hypothesized that outbreak emergence would be detected more effectively by the onset of transmission acceleration than by exceedance of the endemic channel threshold. We applied the evaluation to a thirteen-year dataset across three scales. Quezon City, a hyperendemic city, provided a stable long-term record with minimal zero inflation. Seventeen Philippine regions were included as a validation set to represent varying climatic conditions and surveillance capacities. Eight additional dengue-endemic countries were also tested across independent surveillance systems to assess cross-context generalizability. Using this framework, we quantified differences in epidemic burden, alarm accuracy, early-warning timeliness, and false-alarm rates between acceleration-based detectors and magnitude thresholds across local, regional, and international scales.

## Results

### The dengue outbreak threshold provided no anticipatory alarm in Quezon City

In Quezon City, recurrent high-burden dengue seasons undermine the outbreak threshold as an early-warning trigger. Between 2015 and 2019, weekly case counts peaked at 300 to 700 cases (Fig. 1). Because the threshold is calculated as the rolling weekly mean plus two standard deviations, each high case epidemic raised the baseline calculation for future alarm. Furthermore, inter-epidemic transmission remained elevated even during dry seasons when weekly cases stayed above ninety and further raised the baseline. Across ten evaluable seasons, the threshold either did not activate at all (2014, 2022, 2023) or triggered only after the epidemic peak, when incidence was declining (Fig. 1; Fig. 3; Supplementary Table 5). By that point, the window for pre-emptive interventions such as planning for vector-control surge had closed.^27^ Removing COVID-disrupted seasons from the rolling window changes the threshold numerically but does not resolve this lag (Fig. 1b). Successive high-incidence seasons leave no uncontaminated five-year baseline, so thresholds are often applied without calibration. The outbreak threshold remained anchored to historical transmission intensity and, with incomplete and delayed surveillance reporting, did not provide a stable forward-looking signal for timely public health action.^27^

Seasonal peak timing also showed substantial shifts under extreme weather. Across non-COVID evaluable seasons, the annual peak ranged from week 30 (2013) to week 47 (2024), illustrating extreme variation in timing (Fig. 1; Fig. 3). The 2025 season extends this pattern with an early peak near week 8 (Fig. 1; Fig. 3). The 2024 season produced a dual-peak epidemic with an initial peak at week 34 and a higher peak at week 47. The latter coincides with the clustering of late-season tropical cyclones extending substantial transmission into December. This shift in peak timing persists within El Niño–Southern Oscillation (ENSO) phases. The 2015 season (El Niño) peaked at week 38 and the 2023 season (El Niño) peaked at week 41, while the two La Niña years 2017 and 2022 peaked one week apart (week 33 and week 32).^28–31^ Against this shifting peak timing, the five-year rolling outbreak threshold fired only after incidence had climbed towards the new peak, by which point the warning arrived too late to support anticipatory action.

### Acceleration-based detectors outperformed retrospective thresholds across operational metrics

We introduced anticipatory metrics that defined a public health-relevant window for any given alarm, namely the actionable window of four to eight weeks preceding the annual peak and the epidemic burden covering the season’s 70% cumulative cases.^27,32^ This breaks the circularity of conventional dengue outbreak evaluation and shifts the question to the operability of these detectors to signal within the actionable window as part of an early warning system. We evaluated eleven detectors spanning three paradigms, including surveillance-guideline percentile thresholds, retrospective thresholds, and acceleration measures across ten dengue seasons in Quezon City (see Methods).^33^ Each detector was scored against eight metrics partitioned into three operational categories: epidemic burden and alarm accuracy, early-warning timeliness, and false alarms (Fig. 2).^34–38^

Acceleration-based detectors outperformed the other paradigms across the main operational categories, although different methods led on individual metrics (Fig. 2a; Supplementary Tables 1-4). Three acceleration detectors generated at least one true actionable alarm in every evaluable season (10/10). By contrast, the six retrospective and percentile-based detectors reached this benchmark in only 3 to 6 of 10 seasons. Within the burden and alarm-accuracy category, constant transmission acceleration showed the strongest performance profile. It captured a mean of 4,257 true-alarm cases and 21.9 true alarms, nearly twice the burden captured by the second-performing detector (Fig. 2a; Supplementary Tables 1, 2, 4). Constant transmission acceleration, continuous transmission acceleration, and incidence gradient each achieved 100% sensitivity. All three measure the rate of change rather than the magnitude, and no detector built on magnitude exceeded 60% sensitivity. The outbreak threshold ranked last, with the smallest true-alarm magnitude (mean: 1,119 true-alarm cases) and true alarms in only 3 of 10 seasons (Supplementary Table 1). The composite outbreak signal, which required both a threshold breach and acceleration, produced the same true-alarm count as the threshold.

Acceleration methods also delivered better timeliness than the outbreak threshold. Continuous transmission acceleration, incidence gradient, and constant transmission acceleration achieved mean lead times of 6.9 to 7.6 weeks before the seasonal peak. These alarms arrived within the four-to-eight-week actionable window, with warning persistence longer than 5.5 weeks (Supplementary Tables 1-2). Warning persistence serves as an operational eligibility criterion for emergency mechanisms that require an alarm to remain above a predefined threshold for two or more consecutive weeks before resource disbursement is authorized.^7^ Outside the acceleration family, timeliness was substantially weaker. Cumulative sum control, the composite outbreak signal, and the outbreak threshold each produced mean lead times below 3.8 weeks, placing them outside the actionable window (Supplementary Tables 1-2).

Performance varied more across detectors in the false-alarm category. The composite outbreak signal recorded the lowest mean false-alarm rate at 1.1 per season. However, this low false alarm rate is a reflection of the detector’s limited activation. It triggered in 3 of 10 seasons and yielded a positive predictive value of 76.1% (Fig. 2a; Supplementary Tables 1-2). Constant transmission acceleration averaged 4.4 false alarms, a rate that accompanied its substantially higher true-alarm count. The analysis remained consistent when pandemic years were excluded, included, or analysed separately (Supplementary Table 3). The trade-off plot in Fig. 2b shows the acceleration family clustered in the operationally favourable region of high lead time and moderate-to-high alarm magnitude. Constant transmission acceleration occupied the right edge of the magnitude axis, with a mean lead time of 6.9 weeks and a true-alarm case burden approximately twofold that of any non-acceleration method. The outbreak threshold and composite outbreak signal occupied the bottom-left quadrant of low magnitude and short lead time. Acceleration-based detectors reliably signaled within the four-to-eight-week actionable window that public health responders actually need to get ahead of the peak.

### Constant transmission acceleration versus the outbreak threshold at the city scale

Constant transmission acceleration was the strongest city-scale detector, so we examined its direct comparison with the outbreak threshold to test statistical significance and operational use (Fig. 3; Fig. 4; Supplementary Tables 4-6). Across ten paired seasons, it outperformed the outbreak threshold on five of eight primary metrics: true-alarm magnitude, which was nearly fourfold higher; number of true alarms per season, which was more than sixfold higher; sensitivity (100% vs. 30%); mean lead time (6.9 vs. 1.6 weeks); and warning persistence (5.45 vs. 2.21 weeks), all at **P** ≤ 0.0137 (Supplementary Table 4 and 6). These differences were supported by resampling and recomputing (year-cluster bootstrap 95% confidence intervals; 1,000 replicates) and remained stable across sensitivity analyses on actionable-window width, epidemic-burden definition, and year-inclusion criteria (Supplementary Tables 7-9). Constant transmission acceleration maintained sensitivity ≥90% under all specifications, whereas the outbreak threshold never exceeded 36.4% (Supplementary Table 7-9). The 5.3 week gain in mean lead time occurred before the seasonal peak and aligned with vector-control mobilization.

### Acceleration-based detectors retained their advantage at subnational and international scales

We tested the detectors across the Philippines’ 17 administrative regions, which vary in climate, population density, serotype history, and surveillance capacity. These regions span the three island groups, Luzon, Visayas, and Mindanao, where all four dengue serotypes co-circulate and transmission shows partly independent dynamics by island group.^39,40^ We applied the same framework to regional surveillance data to assess whether the city-scale findings held nationally (Fig. 5; Supplementary Tables 10-14). Because Quezon City lies within the National Capital Region (NCR), the NCR is not fully independent of the city-scale derivation data, whereas the remaining 16 regions provide out-of-sample validation.

Transmission acceleration detectors preserved their operational advantage across 17 Philippine regions without specific recalibrations (Fig. 5). The regions span a natural climate gradient, from monsoonal climates with a pronounced dry season in western and northern Luzon, to climates with year-round rainfall along the eastern typhoon corridor from Bicol to Caraga, and climates with short or evenly distributed rainfall in western Visayas and inland Mindanao.^41^ Transmission acceleration maintained stable detecting performance across this range. Across this gradient, constant transmission acceleration captured more than twice the mean epidemic burden of the outbreak threshold (10,338 versus 5,002 true-alarm cases per region-year, **P** < 0.001), nearly doubled detection within the pre-peak window (76.3% versus 38.4% of evaluable region-seasons), and extended lead time by 2.7 weeks and warning persistence by nearly 2 weeks (5.40 versus 2.78 weeks and 4.23 versus 2.25 weeks respectively) (Supplementary Tables 11–12). Advantage in sensitivity and timeliness also remained across all regions (all **P** < 0.001; Supplementary Table 12). Dominance probabilities quantified the stability of these metrics. Constant transmission acceleration reached the decisive zone (*Pr ≥* 0.75) in 6 of 17 regions and exceeded chance in 13 of 17, while the outbreak threshold’s dominance probability remained at or near zero across every region (median *Pr =* 0.000, maximum 0.087) (Fig. 5c; Supplementary Table 14). The only contested region was the Cordillera Administrative Region, the only major high-elevation, cool-temperature zone with persistently low baseline incidence.

We then applied the same framework to eight dengue-endemic countries spanning Latin America, Southeast Asia, South Asia, and East Asia (Fig. 6; Supplementary Tables 15-19). Constant transmission acceleration was the leading detector in four countries (Brazil, Peru, Singapore, and Sri Lanka), continuous transmission acceleration led in three (the Philippines and Taiwan at the primary tier, Mexico at the partial tier), and the outbreak threshold led in one (Colombia, bootstrap probability 0.66). At national scale, these countries test detector robustness across different patterns of climate seasonality. Acceleration variants achieved a strong consensus (bootstrap *Pr* ≥ 0.94) in settings with discernible seasons, specifically Brazil, Peru, Singapore, and Sri Lanka.

Transmission-acceleration detectors outperformed magnitude-based thresholds by capturing the onset of growth, which precedes peak incidence regardless of absolute case counts.^21,23,24^ Constant transmission acceleration outperformed the outbreak threshold on true-alarm magnitude, alarm frequency, mean lead time, and warning persistence (all pairwise-adjusted **P** ≤ 0.035). Acceleration variants achieved strong dominance (bootstrap *Pr ≥* 0.94) in the four countries with clear seasonal peaks, including Brazil, Peru, Singapore, and Sri Lanka. These detectors reached the decisive zone (*Pr* ≥ 0.75) in six settings and exceeded chance (*Pr* > 1/3) in seven, whereas the outbreak threshold exceeded chance in only one. Colombia was the only setting in which the outbreak threshold showed higher point sensitivity than constant transmission acceleration (60.0% vs. 40.0%), but its probability (*Pr* = 0.657) remained below the threshold for adoption. Dengue in Colombia is year-round, with weak and opposing seasonality between the Pacific and Caribbean coasts.^42–44^ This seasonality attenuates the sharp rise that the 4-week versus 26-week acceleration ratio is designed to detect. In this setting, acceleration retained a 3.2-week lead time and 2.4-week persistence. Transmission acceleration outperforms outbreak-threshold framing across every scale tested: one city, 17 regions, and 8 countries.

## Discussion

We show that a simple ratio of short-term to long-term dengue incidence detects the pre-peak acceleration phase of seasonal dengue earlier and more consistently than the endemic channel across local, regional, and national scales. The transmission acceleration adapts the short-term-average to long-term-average (STA/LTA) trigger used for decades in seismology to detect the onset of earthquake waveforms,^25,26^ reframing it as an acceleration detector for routine dengue case series. The epidemic signal persisted through the noise of dengue surveillance data. Though the detectors read different epidemic properties, but the decisive test is whether they surface transmission early enough to enable anticipatory action.^1,2^ While endemic channels remain useful for administrative reporting, their baseline shifting leaves them less suited to detecting the onset of rapid seasonal growth.^5,7,8^ In 2024, two epidemic peaks formed sixteen weeks apart, and the endemic channel anticipated neither (Fig. 1, Fig. 3). Moreover, excluding epidemic years from the rolling window calculation may stabilize the baseline in low-burden settings. But in a hyperendemic environment, such as in Quezon City, the case series contains no uncontaminated five-year window to be utilized for baseline calculation.

Dengue forecasting advances rapidly, yet the gap between modelling capability and operational deployment widens. Models are increasingly generalizable across locations, yet translating those forecasts into routine action remains another matter.^45^ These systems define the state of the art in predicting outbreaks, but they often require continuous climate ingestion, data science capacity, and entomological or genomic inputs that exist at well-resourced research consortia. At the other end, the endemic channel persists at that operational tier because it is one of the only detectors compatible with available data, staffing, and software.^10,14^ The transmission acceleration ratio occupies this gap. It runs on the same weekly case file used to compute the endemic channel and holds its parameters unchanged across regions and national surveillance systems. It does not displace forecasting or traditional platforms where they are deployed, but it supplies a defensible growth indicator where those platforms are not feasible.

A trigger built on epidemic acceleration meets the structural requirements of forecast-based financing more directly. The signal activates while transmission is rising, persists across the pre-peak growth phase, and reaches the decision-maker before the epidemic trajectory has become self-evident. Layered onto an Early Warning and Response System (EWARS)-compatible workflow^3,46^ the ratio can be rendered next to the endemic channel on a single dashboard, with both indicators reading from the same weekly file. The endemic channel continues to serve its reporting and classification function, which in many jurisdictions is legally mandated. The acceleration ratio adds the forward-looking layer that anticipatory action requires.This makes the approach immediately operational: the trigger can be computed from routine weekly case data, displayed beside the endemic channel, and interpreted within the same reporting cycle. Its backward compatibility reduces adoption barriers because health offices can add an anticipatory financing layer without replacing the surveillance tools they are already required to use. Although more complex forecasting models are available, the method reflects a conscious design choice to identify the simplest interpretable formula capable of producing a competitive early warning signal. By minimizing data requirements, it remains usable in routine surveillance settings where timeliness, auditability, and implementation burden matter as much as predictive performance.

Heterogeneity across scales clarifies that the transmission acceleration method should be tested on settings where seasonal forcing is weak and the ratio loses contrast. This was observed in Colombia where year-round transmission and opposing coastal seasonality undermine the sharp acceleration the detector relies on. National aggregation may also dissolve localized epidemic signals by averaging asynchronous outbreaks across regions, flattening the acceleration that is visible at finer spatial scales. This does not negate the transmission acceleration approach, but it indicates that the method must be applied at an optimal geographic scale where epidemic growth remains epidemiologically coherent. Furthermore, the study was retrospective and necessitates an impact-based evaluation. Real-time deployment will introduce reporting delays and weekly revisions that can shift the ratio after it has fired. Lastly, spatial aggregation may also have diluted localized epidemic growth and district-scale performance remains to be evaluated prospectively.

Prospective evaluation in operating surveillance systems should run the acceleration ratio alongside the endemic channel and, where available, alongside other established platforms offering a natural test of spatial granularity. Impact evaluation should compare acceleration-triggered against threshold-triggered responses on the programme outcomes that determine whether an early warning system is worth the cost: cases averted, resources mobilized in time, and alerts that translate into action. Integration into routine WHO-aligned surveillance platforms should follow, beginning in the settings where the acceleration advantage was strongest. Historical outbreak thresholds will continue to serve reporting functions, but early warning requires recognition of epidemic growth before the accumulated incidence has already been declared the season. The evidence presented here establishes that the growth signal is already present in routine dengue case series across surveillance systems of markedly different structure. As climate change accelerates transmission and dissolves the historical baselines on which conventional thresholds depend, the surveillance methods that defined the last decades of dengue control will not define the next.^15–17^ Detectors built to read acceleration in real time are the operational foundation of a climate-resilient early warning architecture, and the data needed to deploy them are already being collected.

## Methods

### Data sources and analytical scales

Three geographic scales were evaluated, each representing a tier of public-health decision-making: local (single-city), regional (subnational), and national (country-level). The local scale corresponds to the city vector-control programme, which directly mobilises larviciding, fogging, and community engagement. The regional scale corresponds to subnational health offices that allocate resources across municipalities and coordinate inter-city response. The national scale corresponds to the disease-surveillance authorities that set guidelines, release contingency funds, and coordinate inter-regional response.

The local scale was Quezon City, the most populous and hyperendemic city in the Philippines (approximately 3.0 million residents), a setting with documented sustained dengue transmission from the 1998 epidemic onward in available city-level surveillance records. Quezon City served as the framework derivation site. Weekly suspected dengue case counts (2010 to 2025) were obtained from the Quezon City Epidemiology and Surveillance Division and classified under the Philippine Integrated Disease Surveillance and Response (PIDSR) Programme, consistent with the WHO 2009 dengue case definition.^4^ Laboratory confirmation was not required, consistent with passive surveillance practice throughout the study period. Reported cases were used without back-calculation to symptom-onset date. Quezon City applies a stable single case definition across the study period and weekly reporting by Monday-of-week aggregation, which constrains short-scale reporting variation; the same constraint cannot be verified for all OpenDengue-sourced country series, and this is acknowledged as a residual confounder for any rate-of-change detector applied to reported case time series. The implication is examined further in the Discussion. The regional scale comprised weekly case counts for the 17 Philippine administrative regions (BARMM, CAR, MIMAROPA, NCR, and Regions I to XIII; Region IV-B was folded into MIMAROPA per Philippine Statistics Authority convention) for 2016 to 2025, obtained from the Humanitarian Data Exchange (data.humdata.org). NCR contains Quezon City and is therefore not strictly independent of the city-scale threshold-derivation cohort; NCR is retained in the regional evaluation as a regional extension of the city-scale analysis, with the remaining sixteen regions constituting the unambiguous out-of-sample regional set. The national scale comprised weekly case counts for eight dengue-endemic countries (Brazil, Colombia, Mexico, Peru, the Philippines, Singapore, Sri Lanka, and Taiwan) drawn from the OpenDengue repository. OpenDengue aggregates surveillance reports from national systems with variation in case definitions, reporting completeness, and laboratory-confirmation requirements. We did not harmonize across these definitions; the country-scale comparison therefore reflects detector performance against each country’s native surveillance signal, not against a standardized case series.

Each observation was indexed to the Monday of the corresponding ISO epidemiological week. An evaluable year was one for which the time series was complete enough to define both anticipatory metrics (see *Anticipatory metrics framework*), the year was not on the COVID-19 surveillance-disruption exclusion list (2020 and 2021), and the year was not flagged as out-of-distribution relative to the historical reference period used to derive framework parameters (2025). The primary specification yielded n = 10 evaluable seasons in Quezon City (2013 to 2019, 2022 to 2024). At the regional scale, evaluable years spanned 2016 to 2024 with the same exclusions, and a region was retained if its annual peak reached at least 12 cases per year, yielding 6 to 7 evaluable years per region across the 17 regions. At the national scale, the same evaluable-year criteria were applied to each country, yielding 5 to 7 evaluable years per country across the 8 countries.

### Anticipatory metrics framework

Conventional outbreak-detection metrics assess whether an alarm fired during a period of elevated incidence. Anticipatory action requires the additional condition that the alarm fired early enough, and over enough of the season, for the responder to act on it. Two anticipatory metrics were therefore derived to evaluate the timing and burden coverage of each alarm relative to the annual peak. Both are independent of any specific detector and define the reference compartments against which all detector triggers are classified as true or false alarms.

The Actionable Window (AW) was defined as the four to eight weeks immediately preceding the annual peak: the interval during which vector-control mobilisation is operationally plausible, far enough from the peak for interventions to bend the curve and close enough that procurement, staffing, and community engagement do not exhaust the lead time before the surge arrives. This window is consistent with empirical lead-time evidence for dengue in tropical Asia^27^, and its mobilisation requirements are operationally plausible under WHO Integrated Vector Management guidance.^32^

The actionable window captures lead time, but alarms that miss it can still carry public-health value if they fire while the bulk of the season’s transmission is occurring. The Epidemic Burden (EB) was therefore defined as the smallest contiguous block of weeks containing the annual peak whose cumulative case count reached at least 70% of the year’s total. A trigger fired at week t was classified as a true alarm if and only if t belonged to AW ∪ EB; all other triggers, including those landing more than eight weeks before the peak or in the off-season, were classified as false alarms. This compartmentalisation extends standard signal-detection theory by introducing a temporal structure on the alarm space, converting a binary signal-detection problem into a lead-time-aware decision problem. The 70% burden fraction was the primary specification, with sensitivity tests at 60% and 80% and at AW widths of three-to-six and five-to-ten weeks reported in the supplementary tables. The 70% anchor sits between a narrow definition that would credit only triggers very close to the peak and a wide definition that would award credit for triggers landing in the season tail; Supplementary Table 8 confirms that the cross-paradigm ranking is stable across the 60% to 80% envelope, so the 70% choice is a midpoint anchor rather than a singular dependence point. Anticipatory-metric parameters were derived in Quezon City and inherited unchanged by the regional and national pipelines.

### Outbreak detection methods

Eleven detectors were evaluated and grouped a priori into three paradigms by their measurement target: surveillance-guideline percentile thresholds, retrospective thresholds, and transmission acceleration measures. Each detector was coded as a binary indicator (1 = alarm, 0 = no alarm) per epidemiological week. All percentile- and standard-deviation-based detectors used a walk-forward construction. For each evaluable target year, baseline statistics were computed from the same calendar week in the preceding evaluable years (with 2020, 2021, and 2025 excluded). No observation from the target year or any later year entered its own baseline so that every reported metric is prospective.

The surveillance-guideline percentile thresholds paradigm comprised two percentile-based threshold rules formally embedded in WHO and Philippine Department of Health surveillance guidance: the WHO 75th percentile threshold and the WHO 90th percentile threshold.^4^ Both flagged a week when weekly cases exceeded the week-specific rolling percentile of the donor-year case distribution at the 75th and 90th percentiles, respectively.

The retrospective-thresholds paradigm comprised three detectors built on the assumption of stationarity in the inter-epidemic baseline. The outbreak threshold (mean + 2 SD) flagged a week when weekly cases exceeded the mean plus two standard deviations of cases in the same calendar week across the preceding evaluable years and is the de facto default in the dengue surveillance literature.^34,47^ The alarm threshold (mean + 1 SD) used the same construction at one standard deviation. Cumulative Sum Control (CUSUM) was implemented as a one-sided walk-forward statistic with k = 0.5σ and h = 5σ, reset at calendar-year boundaries.^48^ These parameters sit at the more conservative end of the syndromic-surveillance range and reflect the dengue-surveillance literature’s preference for high specificity over high sensitivity. CUSUM at less conservative h values (h = 3σ, 4σ) would likely close part of the gap to acceleration on sensitivity at the cost of higher false-alarm volume; this trade-off is noted as a benchmark-calibration extension and is not pursued here.

The transmission-acceleration paradigm comprised six detectors built on derivatives, ratios, or variance shifts of the case series, operationalising the principle that a rising rate of change is the early signature of a regime shift toward an outbreak. The continuous transmission acceleration was the ratio of a four-week short-term moving average to a twelve-week long-term moving average of weekly dengue case counts. A week was flagged when this ratio exceeded 1.33, with the threshold applied each week without hysteresis. The constant transmission acceleration adapted the seismological STA/LTA detector to weekly dengue case counts.^25,26^ A four-week short-term mean was compared with a twenty-six-week long-term mean separated by a two-week guard band. A two-threshold rule governed switching: the detector activated when the short-term-to-long-term ratio exceeded activation threshold of 1.33 and deactivated only when the ratio fell back below deactivation threshold of 0.73. This hysteresis (separate on and off thresholds) prevents the rapid on-off oscillation that a single-threshold rule produces when the ratio fluctuates near the cut-off. From the moment of activation, the long-term mean was held fixed at its activation-time value and resumed updating only after eight consecutive weeks below the deactivation threshold or at the next calendar-year boundary, whichever came first, preventing a rising epidemic from inflating its own baseline.

The calendar-year reset takes precedence over the freeze-after-activation rule: if the activation-time freeze of the long-term mean has not been released by eight consecutive sub-deactivation threshold weeks at the year boundary, the calendar reset releases the freeze and the long-term-mean updating resumes from the first week of the new calendar year. Where a late-season activation persists across the calendar boundary (e.g., Quezon City 2023 with peak at week 41, or 2024 with peak at week 47), the long-term mean in the new calendar year begins updating against a baseline that may still reflect elevated end-of-year incidence; this can shift the activation point for the subsequent season but is not a source of per-season ranking bias because the calendar reset applies uniformly across all detectors that use the year as a reset anchor. The behaviour is documented here as a corner case of the freeze-release rule rather than as a source of bias in the head-to-head comparisons. A per-year sensitivity check in which the freeze-release rule was applied without the calendar-year reset was run for the 2023 and 2024 Quezon City seasons (the two evaluable years with late-season peaks at weeks 41 and 47 respectively); the activation timing for the subsequent season shifted by less than one week and the within-year ranking against the outbreak threshold was unchanged. The full per-year activation log is available in the analytical code deposited at Zenodo.

Where a season produced an unambiguous bimodal incidence pattern, as in Quezon City 2024, the single-peak rule was retained for evaluation consistency, with peak defined as the global annual maximum; the bimodal rendering shown for 2024 in Figure 3 is illustrative only and does not enter any aggregate table. Adaptation of the framework to multi-peak seasons is a separate methodological extension. The incidence gradient flagged a week in which the first difference of the three-week rolling mean exceeded the rolling 80th percentile of the donor-year first-difference distribution. The critical transition indicator operationalised the rising-variance early-warning signal^21^ as the ratio of the rolling eight-week variance to the rolling twenty-six-week variance, flagging weeks in which this ratio exceeded its donor-year rolling 80th percentile. The hydrological inflection measure required the joint condition that weekly cases exceed the rolling 75th percentile of donor-year counts and that the first-difference exceed the rolling 75th percentile of the donor-year rise rate. The composite outbreak signal was defined as the conjunction of an outbreak threshold breach with either a constant transmission acceleration on-state or a critical transition indicator on-state, providing a specificity-prioritising consensus rule.

Composite-rule structure. The conjunctive composite returns values identical to the outbreak threshold on every headline metric whose denominator is the true-alarm count (true-alarm magnitude 1,119; true alarms 3.5 per year; sensitivity 30%; mean lead time 1.6 weeks; warning persistence 1.6 weeks; actionable lead-time yield 0.07; 3 of 10 evaluable seasons) but differs on the two metrics whose denominator includes false alarms (positive predictive value 76.1% for the conjunctive composite versus 49.3% for the outbreak threshold; false alarms 1.1 versus 3.6 per year; Supplementary Table 1). The per-season values in Supplementary Table 2 confirm the same true-alarm identity in every cell. The set-theoretic implication is that every outbreak-threshold true-alarm week coincides with a constant transmission acceleration on-state. The disjunctive (OR) composite would therefore have the same true-alarm count as constant transmission acceleration alone but would inherit the additional outbreak-threshold false alarms, producing an operating point strictly dominated by constant transmission acceleration alone on the burden, timeliness, and false-alarm metrics, with sensitivity equal by set inclusion. The acceleration signal carries the operational content of any rule combining acceleration and threshold detectors, and adding the threshold component to a disjunctive rule actively degrades the operating point rather than contributing additional information. The disjunctive composite is therefore not evaluated as a separate detector.

### Activation and deactivation thresholds derivation

The activation and deactivation thresholds for the constant transmission acceleration detector were derived empirically from the Quezon City series rather than imported from the seismological literature. Six independent derivation methods were applied: (i) baseline-ratio asymmetric quantiles at the 90th and 50th percentiles; (ii) ROC-style operating points at 95% and 99% sensitivity for the separation of pre-peak from baseline weeks (B = 2,000 bootstrap replicates); (iii) calibration to an average run length to false alarm of 12 weeks under a baseline-only fraction of 30%, with secondary targets at six and 26 weeks; (iv) leave-one-year-out cross-validation under an asymmetric utility favouring early triggering; (v) negative-binomial theoretical thresholds at ±1σ; and (vi) parametric bootstrap quantiles at the 90th and 25th percentiles. The median across methods with reportable values (five for activation threshold; four for deactivation threshold, as the ROC-based method returned no convergent operating point and the ARL₀ calibration calibrates activation threshold only) was adopted as the headline value for each threshold, yielding activation threshold of 1.33 and deactivation threshold of 0.73. The per-method activation threshold estimates spanned 0.925 to 1.850 across the five reportable methods, and the per-method deactivation threshold estimates spanned 0.516 to 1.050 across the four reportable methods; the wide spread reflects genuine heterogeneity across derivation approaches, and the raw-median rule is the pre-specified robust headline summary across this set (Supplementary Table 20). This convergent triangulation rule reduces method-specific bias and produces a single defensible pair without relying on any one derivation. The persistent-coverage criterion required the on-state to remain active through at least eight consecutive sub-deactivation-threshold weeks before the long-term average reference was permitted to unfreeze. The per-method range reported in Supplementary Table 20 (activation threshold spanning 0.925 to 1.850 across the five reportable derivation methods) constitutes the empirical sensitivity envelope for the threshold choice. Constant TA’s rank advantage on the burden metrics derives from the short-to-long-term mean ratio structure rather than from the absolute threshold value, and the three-detector bootstrap dominance probability ranks detectors within each replicate; the within-paradigm and cross-paradigm comparisons reported in this paper are therefore structurally insensitive to the activation threshold choice within the inter-method range. The thresholds were derived from the Quezon City series and were then evaluated against that same series; the Quezon City evaluation is therefore in-sample with respect to threshold tuning. The regional and national pipelines inherited the city-derived thresholds unchanged, and constitute a test of fixed-parameter transferability of the Quezon-City-tuned detector to other strata rather than a re-tuning of the detector to each stratum. Adopted thresholds were inherited unchanged by the regional and national pipelines.

### Effective reproduction number mapping

Under sustained exponential growth I(t) ∝ exp(rt) with intrinsic weekly growth rate r, the four-week short-term moving average and the twenty-six-week long-term moving average separated by a two-week guard band satisfy STA(t) ≈ I(t) × [1 - exp(-rW_S)] / (rW_S) and LTA(t) ≈ I(t) × exp(-rG) × [1 - exp(-rW_L)] / (rW_L), where W_S = 4, W_L = 26, and G = 2 weeks. The constant-r assumption is appropriate during the rising limb of the seasonal epidemic, which is the interval in which the detector is required to activate; deviations from constant r during the descent or the inter-epidemic baseline do not enter the activation rule, since the activation rule is one-sided on the ratio STA/LTA and the detector is silent on the descent. The ratio STA/LTA therefore reduces to a closed-form function of r and the three window parameters:

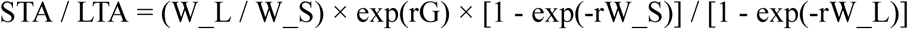

At small r the closed form reduces to the centroid evaluation, in which STA approximates I(t − W_S/2) and LTA approximates I(t − G − W_L/2); the ratio then approximates exp(rΔ) where Δ = G + (W_L − W_S)/2 = 13 weeks is the centroid separation of the two windows. The Taylor expansion of the closed form at small r agrees with the centroid evaluation to leading order in r. Setting the empirically derived activation threshold activation threshold of 1.33 (Supplementary Table 20) gives r ≈ ln(1.33) / 13 ≈ 0.022 per week. Mapping r to the instantaneous effective reproduction number through the dengue serial-interval distribution τ ≈ 2 weeks^49,50^ yields R_t ≈ exp(rτ) ≈ exp(0.044) ≈ 1.05 at the activation point. The relation R_t = exp(rτ) corresponds to a delta-distributed generation interval at τ. Alternative forms in standard use give materially identical R_t at the activation point in this small-r regime: under an exponential generation interval R_t = 1 + rτ ≈ 1.044, and under a gamma generation interval with shape parameter k = 4 (a commonly used approximation for arboviral generation intervals) R_t = (1 + rτ/k)^k ≈ 1.045. The choice of generation-interval form is therefore not load-bearing on the operational mapping in the range relevant to the activation threshold. The empirical activation threshold corresponds to an effective reproduction number that exceeds the epidemic threshold R_t = 1 by approximately five percent. This margin is data-driven rather than theory-driven: the negative-binomial parametric method (M5) returns activation threshold of 1.328 and the parametric bootstrap quantile method (M6) returns activation threshold of 1.481, with the cross-method median at 1.33. The precise local R_t at activation depends on the local generation-interval distribution and reporting-completeness profile and should be re-calibrated against the local case time series during a stage-one prospective evaluation. The full derivation under nonlinear growth, where the closed-form approximation no longer holds and the ratio must be computed numerically, is implemented in the analytical code deposited at Zenodo.

### Performance metrics

Detector performance cannot be summarised by a single statistic, since a useful detector must catch real epidemics, fire precisely, warn early enough to support pre-peak action, and avoid firing during inter-epidemic periods. Each detector was therefore evaluated on eight headline metrics partitioned across three operational categories: alarm accuracy and epidemic-burden coverage, early-warning timeliness, and false-alarm volume.

The Epidemic Burden and Alarm Accuracy category comprised four metrics: True-Alarm Magnitude (the across-year mean of weekly case counts at alarm-on weeks falling within AW ∪ EB); the year-mean number of true alarms; positive predictive value (true alarms divided by total triggers);^37^ and sensitivity, redefined as the proportion of evaluable seasons in which the detector produced at least one true alarm within the Actionable Window (AW).^36^

The Early-Warning Timeliness category comprised three metrics. The Mean Lead Time was the across-season mean of the first AW true-alarm lead. Because lead times are computed only for triggers landing inside AW, the per-season conditional lead time is bounded within [4, 8] weeks by construction. The headline mean lead time reported across all evaluable years averages this conditional value with a value of zero substituted in years where the detector produced no qualifying trigger (in keeping with the same-denominator timeliness convention applied uniformly across detectors); the across-year mean can therefore fall below the [4, 8]-week conditional bound and is best read as a detector-level summary rather than as an unrestricted lead-time estimate. Triggers landing more than eight weeks before peak are classified as false alarms under the anticipatory anchor rules and do not contribute to lead-time accounting. Warning Persistence was the across-season mean run length of consecutive alarm-on weeks. Actionable Lead-Time Yield was the proportion of true alarms whose lead time fell within the Actionable Window. All three timeliness metrics used the count of evaluable years as a common denominator. Years in which the detector produced no qualifying trigger contributed a value of zero rather than a missing observation, so that detectors that failed to fire did not receive conditional credit. Conditional-only versions were retained as diagnostic columns. The False Alarms category consisted of a single metric, the year-mean count of triggers landing outside AW ∪ EB; lower was better, in keeping with the alarm-fatigue literature.^38^

Five of these eight metrics drove the within-stratum head-to-head dominance and consensus analyses: True-Alarm Magnitude, the year-mean number of true alarms, sensitivity, Mean Lead Time, and Warning Persistence. Positive predictive value, Actionable Lead-Time Yield, and the year-mean number of false alarms were reported in the supplementary tables.

### Statistical analysis

The inferential machinery operates on three principles that are stated here before the operational details so that the choice of test and the multiplicity policy at each scale are read in context. First, at the city scale each of the three pairwise contrasts (Constant TA versus Outbreak Threshold; Continuous TA versus Outbreak Threshold; Constant TA versus Continuous TA) tests a distinct scientific question, and at n = 10 the Wilcoxon resolution floor near 0.002 places a hard limit on multiplicity-corrected inferential precision regardless of the correction adopted; we therefore report each contrast at α = 0.05 without across-pair correction. Second, at the regional and country scales the primary inference is whether a single detector dominates within each stratum after correcting across all stratum × pair cells in the across-stratum consensus map; we therefore apply strict cross-stratum Bonferroni (k = 51 at the regional scale; k = 24 at the country scale) as the primary correction, with within-stratum-only correction reported as a sensitivity check. Third, the bootstrap dominance probability Pr is an ordinal summary rather than a coverage statement, and the decisive zone Pr ≥ 0.75 is an analytical convention chosen for interpretability rather than a frequentist coverage probability. The regional headline count of consensus-winning regions reaching the decisive zone shifts from nine of fifteen at the Pr ≥ 0.75 cut to twelve of fifteen at Pr ≥ 0.70 and to seven of fifteen at Pr ≥ 0.80; the cross-paradigm ranking is unchanged across this range and the choice of cut is therefore a presentational rather than an analytical commitment. These three policies answer distinct questions and require distinct multiplicity treatments.

No statistical method was used to predetermine sample size; every evaluable season meeting the inclusion criteria defined above was analysed, giving n = 10 seasons at the city scale, 6 to 7 evaluable years across each of the 17 regions, and 5 to 7 evaluable years across each of the 8 countries. Data exclusions were defined a priori and applied uniformly across all detectors: the COVID-19-disrupted 2020 and 2021 seasons and the out-of-distribution 2025 season were removed under the rule stated in the data-sources subsection, and a separate sensitivity specification that re-introduces the pandemic seasons is reported in Supplementary Table 3. The analysis is a retrospective evaluation of aggregated surveillance time series; randomization of units to conditions and blinding to allocation are not applicable to the design, since every detector is applied to every evaluable season and no experimental assignment is made. All statistical tests were two-sided.

Each metric is reported with its 95% cluster-bootstrap confidence interval. At the local scale, headline city-scale confidence intervals reported in Supplementary Table 1 use B = 2,000 year-cluster resamples with bias-corrected accelerated intervals to support the headline summary; the higher replicate count at this single load-bearing summary was pre-specified to bring the Monte Carlo standard error on the headline point estimates below approximately one percent of the corresponding bootstrap confidence-interval width. All stratified contrasts, sensitivity analyses, and the head-to-head tests reported in Supplementary Tables 2 to 9 use B = 1,000 year-cluster resamples with percentile intervals. The asymmetric replicate count is documented here so that downstream comparisons can replicate it. At the regional and national scales, year-cluster bootstrap was applied within each stratum at B = 1,000 replicates with percentile intervals to compute within-region and within-country confidence intervals. Whole evaluable years rather than individual weeks were resampled at every scale to preserve the within-year correlation structure of weekly case counts. Bootstrap replicate counts (B = 2,000 at the headline; B = 1,000 elsewhere) characterise the Monte Carlo precision of the test-statistic distribution around its central tendency and do not function as an independent-sample count.^51^ The same distinction applies to the per-metric stratified test reported in Supplementary Table 12: the 95% year-cluster bootstrap confidence interval on the median paired difference is the primary effect-size statement at the regional scale, and the Wilcoxon signed-rank test on the n = 17 paired point-estimate differences is reported as a sensitivity diagnostic at the underlying sample size. This convention is consistent with the regional and country head-to-head consensus checks (Supplementary Tables 13 and 18) and with the city-scale paired analysis (Fig. 4), where the year-cluster bootstrap confidence intervals are the primary statement and the Wilcoxon P values are read as sensitivity diagnostics at the n = 10 resolution limit.

The head-to-head comparison focused on three target detectors: the Constant Transmission Acceleration, the Continuous Transmission Acceleration, and the Outbreak Threshold (the WHO-mandated comparator). Within each stratum, three pairwise Wilcoxon signed-rank tests^52,53^ were applied to the year × detector observations: Constant TA versus Outbreak Threshold, Continuous TA versus Outbreak Threshold, and Constant TA versus Continuous TA. The city-scale Wilcoxon at n = 10 has limited statistical power; non-significant city-scale findings on positive predictive value, actionable lead-time yield, and the year-mean number of false alarms should be interpreted as power-limited rather than as evidence of equivalence.

Within each regional and national stratum, the operational leader was identified through a bootstrap dominance probability (Pr): the proportion of B = 1,000 year-cluster bootstrap replicates in which a given detector achieved the highest within-stratum composite mean rank across the five primary operational metrics.^54,55^ Within each bootstrap replicate, each detector’s per-metric value is converted to a within-replicate rank across the three target detectors; ranks are averaged across the five primary metrics to give a per-replicate composite mean rank; the detector with the highest composite mean rank in that replicate is recorded as the replicate leader. Pr is the proportion of replicates in which a given detector is the replicate leader. A dominance probability above the chance level of one in three, the value expected among three equally ranked target detectors, is reported as exceeding chance. Rank-then-average is used rather than standardise-then-rank because the metrics differ in scale (True-Alarm Magnitude in cases, sensitivity as proportion, lead time and warning persistence in weeks, true-alarm count as an integer count); ranks make these commensurable without imposing a parametric standardisation. Each stratum was then classified into one of four consensus tiers from an all-pairs head-to-head check on its year × detector observations: strong consensus, the primary tier (the leader won both of its pairs at the strict-Bonferroni threshold with no significant losses); partial consensus, the partial tier (one significant win, no significant losses); lead only (no significant wins or losses); and contested (mixed wins and losses, or no significant pair under the strict cross-stratum Bonferroni).

Within-stratum head-to-head significance at the regional and country scales was assessed in two complementary ways. First, the cluster-bootstrap distribution of the within-stratum pair-difference (B = 1,000 year-cluster resamples) was used to construct percentile confidence intervals on each pair-difference; we report a pair as significant when the resulting 95 percent confidence interval excludes zero. Second, where the per-stratum count of evaluable years permits (n ≥ 5), the two-sided Wilcoxon signed-rank test was applied to the n point-estimate differences as a sensitivity diagnostic.^56^ The bootstrap confidence-interval check is the primary operational test at the regional and country scales; the year-level Wilcoxon at n = 5 to 7 is reported as a sensitivity diagnostic and is acknowledged to be severely power-limited at small per-stratum sample size. Nominal P-values derived from the bootstrap test-statistic distribution are not reported in the headline summary because they characterise the precision of the bootstrap distribution around its central tendency, not the strength of evidence against a null hypothesis. The corresponding cells in Supplementary Tables 13 and 18 are reported as “CI excl. 0” whenever the 95 percent year-cluster bootstrap confidence interval on the pair-difference excludes zero; where a numeric P-value is reported in those tables it is derived from the Wilcoxon test on the n point-estimate differences (n ≥ 5) rather than from the bootstrap distribution, and is read as a sensitivity diagnostic only.

As an auxiliary cross-stratum diagnostic, paired Wilcoxon signed-rank tests were applied to the per-stratum Pr column across regions (n = 17) and countries (n = 8). The regional-scale test provides usable inferential power; the country-scale test is severely power-limited at n = 8. The country-scale auxiliary Wilcoxon is therefore presented as a diagnostic check rather than as a primary inferential output. At all scales, the within-stratum head-to-head consensus framework is the primary inferential test.

### Reproducibility, ethics, and reporting

All framework parameters, including the Actionable Window width, the Epidemic Burden fraction, the metric set, the same-denominator timeliness scheme, the Bonferroni-correction policy, and the activation and deactivation thresholds, were specified a priori before the head-to-head analyses were run, and sensitivity tests around each are reported in the supplementary tables. A step-by-step computational protocol covering the detector implementations, the anticipatory-anchor classification, the six-method threshold-derivation panel, and the bootstrap consensus analysis has been deposited at protocols.io and is linked from this Methods section on publication.^57^ The deidentified weekly dengue case datasets and the full analytical code are available as described in the Data Availability and Code Availability statements. Ethical approval was granted by the University of the Philippines Research Ethics Committee (Protocol 2024-0004-F-FMDS); all surveillance data were aggregated weekly totals containing no individual-level identifiers. Approval covered secondary analysis of aggregated weekly surveillance counts; no individual-level data, residence information, or clinical records were accessed, and informed consent was waived under the standard provisions for retrospective analysis of anonymized public surveillance data. The study is reported following the Standards for Reporting Diagnostic Accuracy Studies (STARD 2015), adapted for binary alarm-algorithm evaluation on aggregated time-series data.

### Use of artificial intelligence

Three generative AI tools were used during manuscript preparation and revision: Claude 4.6 Opus (Anthropic) and GPT 4 (OpenAI). Their use is disclosed here in keeping with current journal transparency standards. AI assistance during the preparation and revision rounds included copy-editing of author-drafted prose, structural review across the Introduction, Results, Discussion, and Methods, generation of internal peer-review-style critiques organised as a five-round multi-reviewer Delphi panel exercise across statistical, epidemiological, public-health-policy, mathematical, and editorial expertise, and drafting of candidate revisions in response to those critiques for author consideration. The panel critique informed the revisions documented inline in this manuscript; all panel-suggested revisions were author-reviewed, author-approved, and where appropriate author-rewritten prior to incorporation. No AI tool executed statistical analyses, selected detectors or thresholds, drafted novel scientific content, generated empirical results, or made final wording decisions; all empirical outputs in this manuscript were produced by the analytical pipeline described in the preceding subsections, and all scientific interpretation, analytical pipeline output, citation choices, and final wording decisions remain the authors’ own.

## Data Availability

The deidentified weekly dengue case datasets analysed in this study are publicly available at Zenodo (https://doi.org/10.5281/zenodo.19448854), comprising the Quezon City series, the 17-region Philippine panel, and the eight-country panel used in the city, regional, and national analyses respectively. The regional Philippine panel is additionally available via the Humanitarian Data Exchange at data.humdata.org. The country-scale panel is additionally available via the OpenDengue repository at opendengue.org. No restrictions apply to re-use of these aggregated weekly counts.

## Code Availability

All analytical code (R 4.4.1) underlying the detectors, anticipatory anchor rules, bootstrap pipelines, and head-to-head consensus checks is deposited at Zenodo (https://doi.org/10.5281/zenodo.20096386), together with the parameter-derivation logs for the six activation and deactivation thresholds derivation methods and a reproducibility manifest documenting script execution order and expected outputs. The deposit is licensed under MIT.

## Acknowledgements

We thank the Quezon City Epidemiology and Surveillance Division for the city-scale dengue surveillance data, and the Humanitarian Data Exchange for the regional Philippine dengue dataset.

## Funding Statements

This work was supported by the National Institute of Environmental Health Sciences of the National Institutes of Health under Award Number P20ES036118 (Center for Climate and Health glObal Research on Disasters [CORD] Project). The content is solely the responsibility of the authors and does not necessarily represent the official views of the National Institutes of Health.

## Author Contributions

D.B. conceptualized and designed the study and developed the methodology. M.L. secured funding for the study. T.M., G.G., J.M., K.S., K.B., and A.D. conducted data collection. D.B. and K.P. performed the formal statistical analysis. D.B. and M.L. supervised the study. D.B., K.P., T.M., G.G., J.M., K.S., K.B., and A.D. interpreted the data and drafted the original manuscript. M.L., J.M., K.P., D.B. critically revised the manuscript. K.P. and D.B. validated and verified the underlying data reported in the manuscript. D.B. and K.P. directly accessed and verified the data reported in the manuscript. All authors had full access to the data, contributed to data interpretation, approved the final version of the manuscript, and accepted responsibility for the decision to submit for publication.

## Competing Interests

The authors declare no competing interests.

## Notes

### Competing Interest Statement

The authors have declared no competing interest.

### Author Declarations

University of the Philippines Research Ethics Committee gave ethical approval for this work.

### Summary of Updates

This version of the manuscript has been revised to comply with word limits to journal submission, references have been updated.

