## Supplementary material for "Transmission acceleration outperforms the endemic-channel threshold for dengue outbreak detection": Figures and Supplemental Tables

**Fig. 1 | Shifting alarm and outbreak thresholds in Quezon City, 2013–2025.**

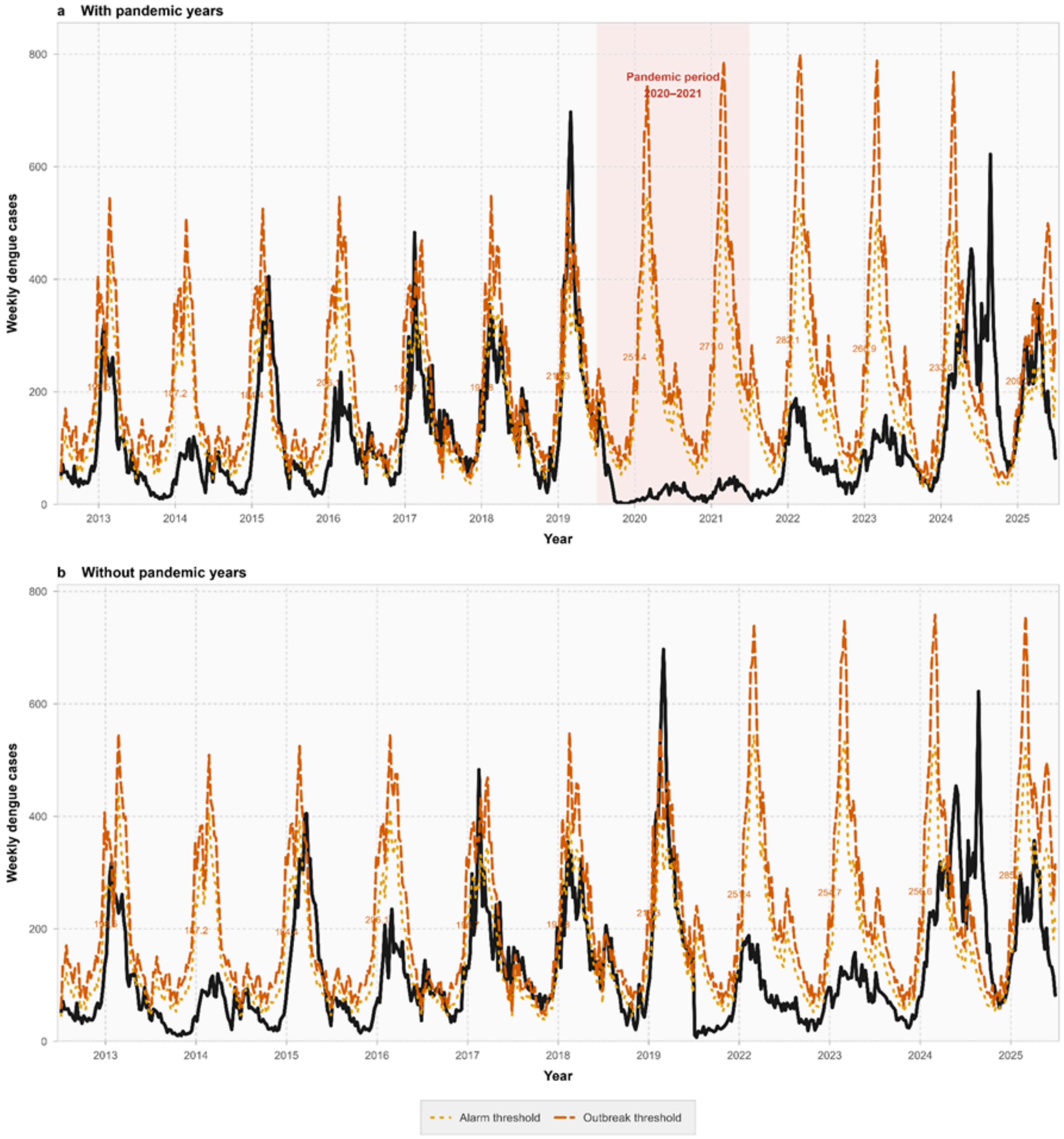


Weekly dengue case counts (solid black line) are shown against two rolling, week-specific surveillance thresholds: the alarm threshold (dotted orange line) and the outbreak threshold (dashed orange line). a, Thresholds derived from prior-year reference windows that retain the pandemic years; the shaded vertical band labelled "Pandemic period 2020–2021" marks those years. b, Thresholds derived from prior-year reference windows that omit 2020 and 2021. The numerals printed beside each season give the annual mean outbreak-threshold value assigned to that year. Values are descriptive weekly counts and modelled thresholds; no error bars are shown and no statistical test is applied to this panel.

**Fig. 2 | Operational comparison of 11 dengue outbreak-detection methods in Quezon City, Philippines, 2013-2024.**

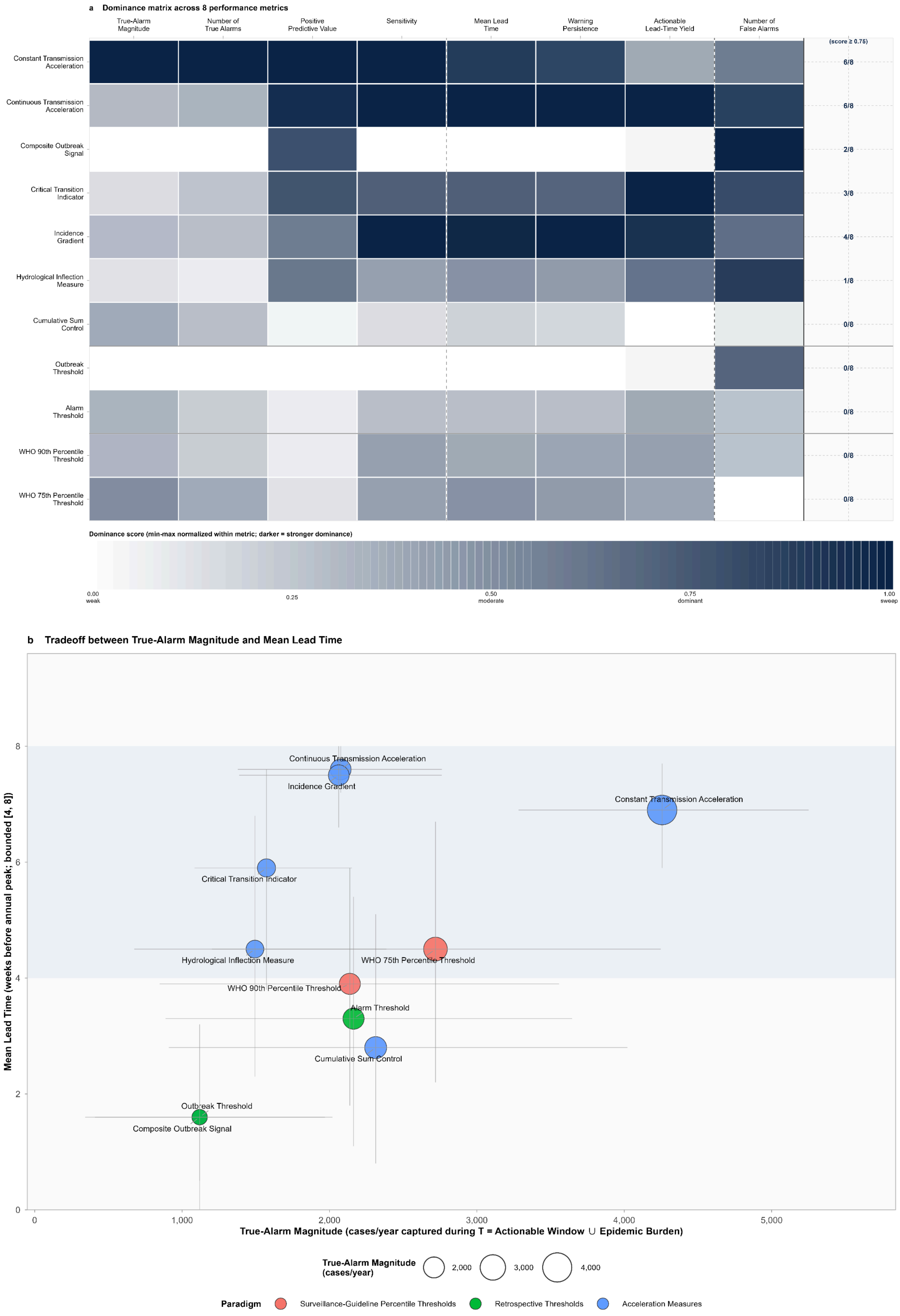

a, Dominance matrix for the 11 detectors (rows) across eight performance metrics (columns), grouped as epidemic burden and alarm accuracy (true-alarm magnitude, number of true alarms, positive predictive value, sensitivity), early-warning timeliness (mean lead time, warning persistence, actionable lead-time yield) and false alarms (number of false alarms). Cell shading shows the within-metric dominance score on a single-hue blue scale from white (lowest, score 0) to dark navy (highest, score 1). The rightmost column gives the dominance count, the number of metrics out of eight for which a detector scores at or above 0.75. b, Trade-off between true-alarm magnitude (x axis, cases per year captured by true alarms) and mean lead time (y axis, weeks of warning per year). Each marker is one detector and marker area scales linearly with true-alarm magnitude. Horizontal and vertical bars are year-cluster bootstrap 95% confidence intervals (1,000 replicates). The pale blue horizontal band marks the actionable window, 4 to 8 weeks before the peak. Marker colour denotes detection paradigm: national-standard methods (red), retrospective thresholds (green) and acceleration measures (blue). Ten seasons are evaluable; 2020, 2021 and 2025 are excluded.

### Fig. 3 | Constant transmission acceleration detection timing in Quezon City, 2013-2025.

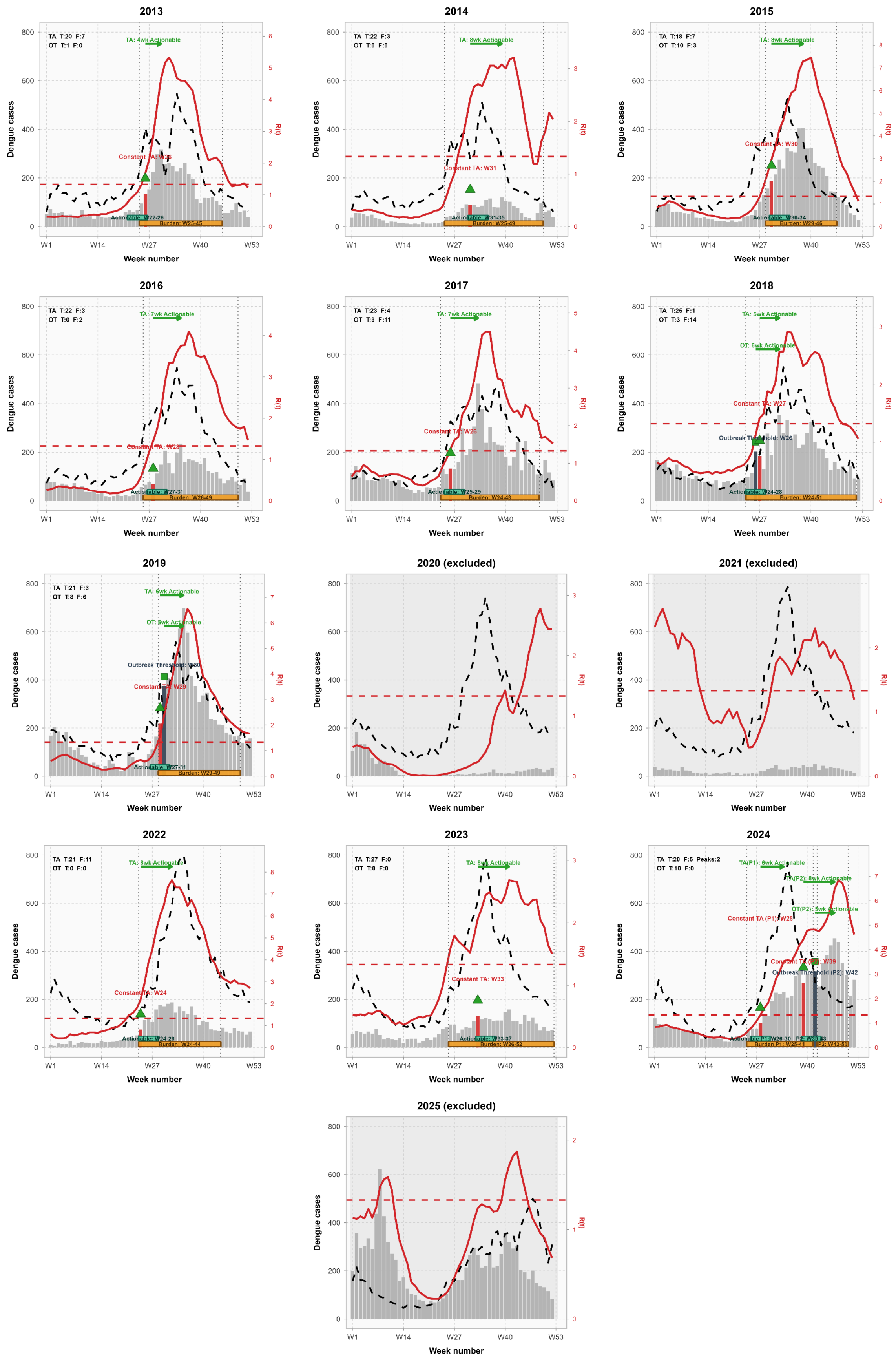

Each panel is one season. Grey bars are weekly dengue case counts (left axis). The dashed black line is the rolling outbreak threshold and the solid red line is the constant transmission acceleration ratio (right axis), with the horizontal dashed red line marking its activation level. Detector triggers are plotted as markers coded by detector: upward green triangles for constant transmission acceleration and green squares for the outbreak threshold; for clarity only the first within-window trigger per detector per season is drawn, and the full inventory is given in Supplementary Table 5. Two anticipatory anchor bands run along the foot of each panel: a wider amber band for the epidemic-burden anchor and a narrower teal band for the actionable window; a trigger inside either band is a true alarm, otherwise a false alarm. Per-panel labels report within-season true (T) and false (F) alarm counts for constant transmission acceleration (TA) and the outbreak threshold (OT). The 2020 and 2021 seasons are shown but excluded from analysis owing to COVID-19 surveillance disruption, and 2025 is excluded as out of distribution; these three panels are labelled “excluded”. The 2024 panel uses an illustrative two-peak layout for display only; all quantitative results use the single annual-maximum peak. Bars are observed weekly counts and carry no error bars.

**Fig. 4 | Paired-year comparison of constant transmission acceleration and the outbreak threshold across eight operational performance metrics, Quezon City, Philippines, 2013-2024.**

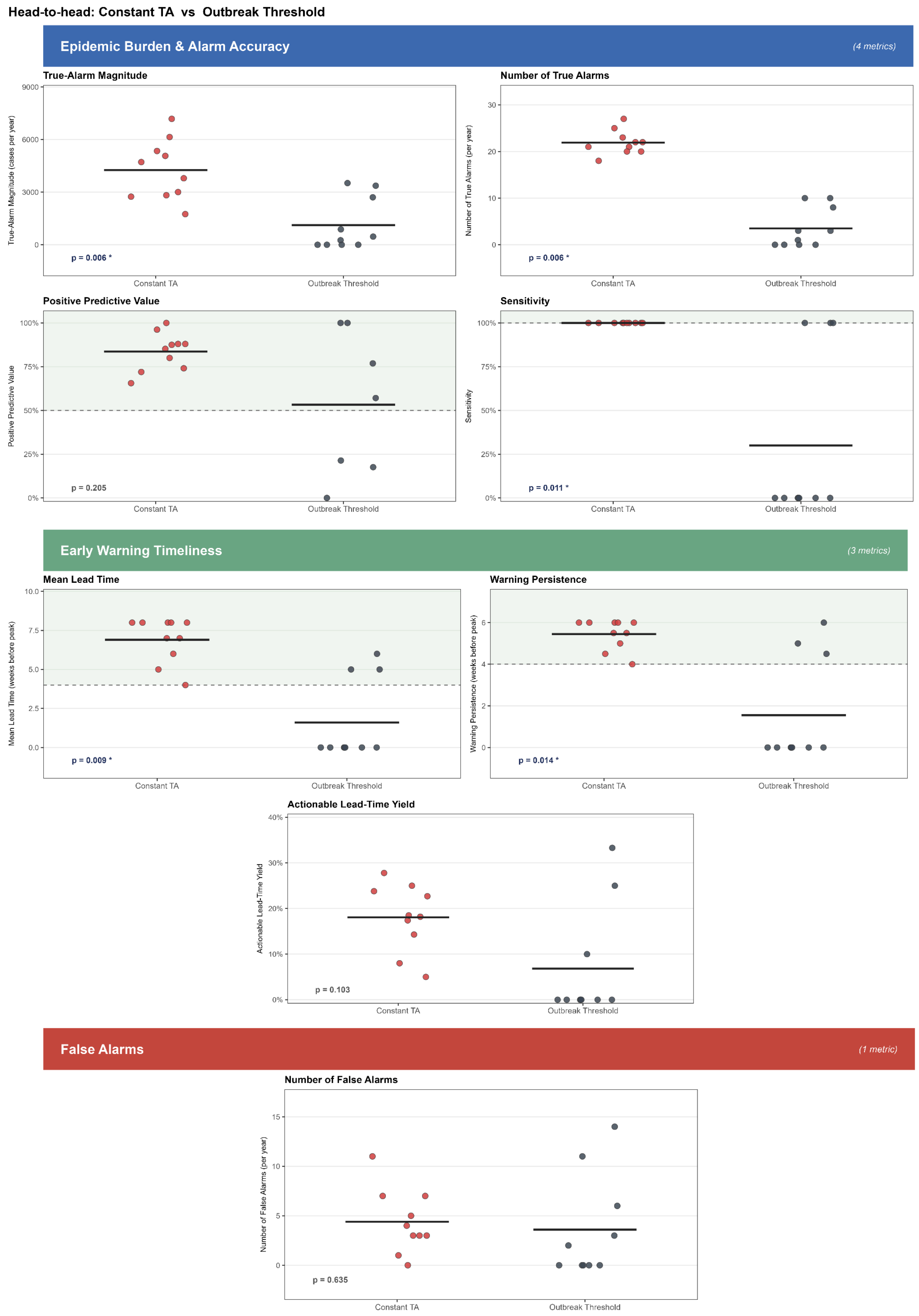

Constant transmission acceleration is contrasted with the outbreak threshold across the eight headline metrics, grouped under three banner headers: epidemic burden and alarm accuracy (blue header), early-warning timeliness (green header) and false alarms (red header). In each subpanel one dot is one evaluable season (n = 10; 2020, 2021 and 2025 excluded), jittered horizontally and coloured by detector (constant transmission acceleration, red; outbreak threshold, slate grey). The horizontal crossbar is the across-season mean for each detector. The pale-green region marks the operationally favourable zone, lying above the dashed reference for higher-is-better metrics and below it for the single lower-is-better metric (number of false alarms). The two-sided Wilcoxon paired signed-rank P value, paired by season, is printed at the lower left of each subpanel, with an asterisk when $P < 0.05$. Effect sizes (median paired differences) with year-cluster bootstrap 95% confidence intervals (1,000 replicates) are the primary effect-size statement and are given in Supplementary Table 6. Five of eight metrics reach $P < 0.05$: true-alarm magnitude, number of true alarms, sensitivity, mean lead time and warning persistence; positive predictive value, actionable lead-time yield and number of false alarms do not.

**Fig. 5 | Bootstrap-supported regional dominance and consensus of outbreak detectors across 17 Philippine regions.**

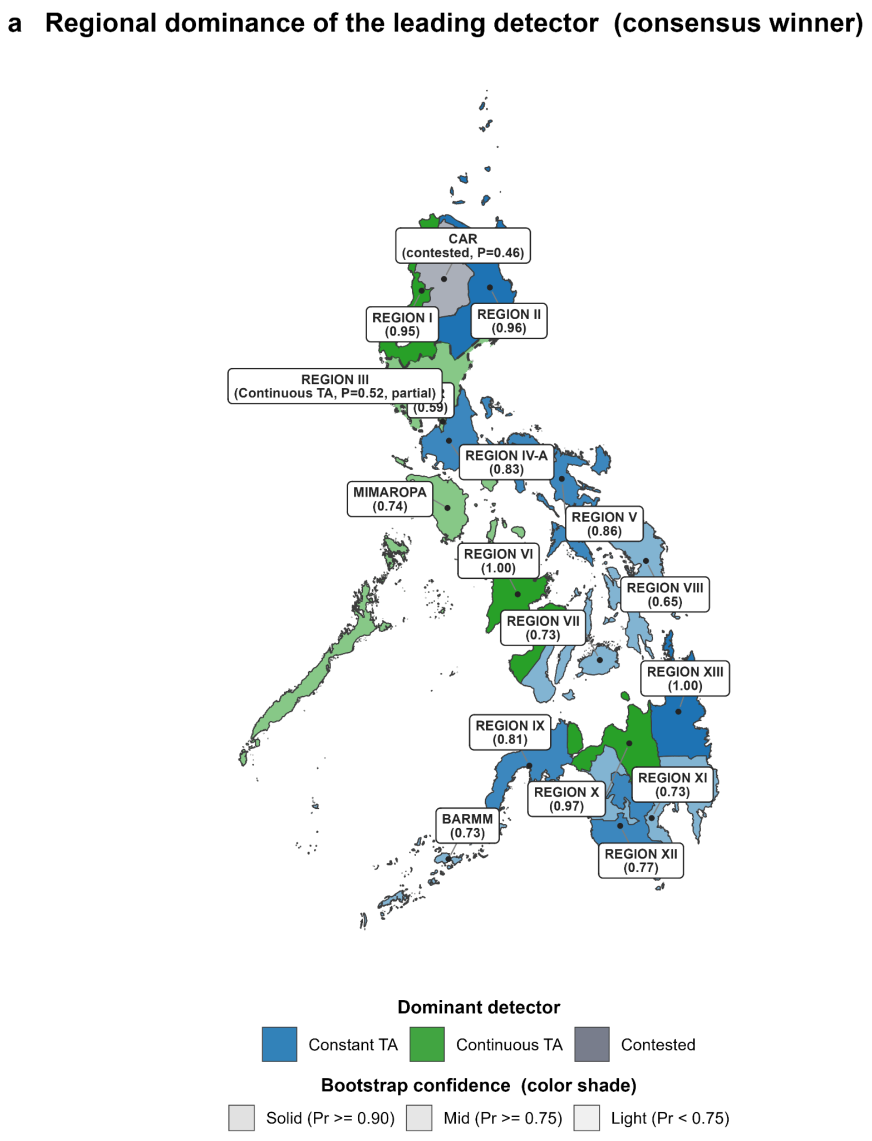

**b Bootstrap-winning detector and its per-metric significance, by region (paired bootstrap, Bonf k = 5)**

| Region | Detector | Dominance probability | TAM | N true alarms | Sensitivity | Mean lead time (wk) | WP (wk) |
|---|---|---|---|---|---|---|---|
| BARMM | Constant TA | 0.73 (***) | 4,037 (**) | 24.7 (**) | 83% (ns) | 6.5 (ns) | 4.9 (ns) |
| CAR | Contested | 0.46 (*) | *5,538* (**) | *10.1* (**) | *86%* (*) | *6.6* (**) | *5.0* (**) |
| MIMAROPA | Continuous TA | 0.74 (***) | 3,682 (ns) | 10.0 (ns) | 100% (ns) | 6.9 (ns) | 5.4 (ns) |
| NCR | Continuous TA | 0.59 (***) | 9,517 (ns) | 10.0 (ns) | 100% (ns) | 7.9 (ns) | 6.2 (**) |
| REGION I | Continuous TA | 0.95 (***) | 5,017 (ns) | 11.3 (ns) | 100% (ns) | 7.9 (**) | 5.9 (**) |
| REGION II | Constant TA | 0.96 (***) | 7,782 (ns) | 18.0 (ns) | 71% (ns) | 5.7 (ns) | 4.3 (ns) |
| REGION III | Continuous TA (partial) | 0.52 (ns) | *12,224* (ns) | *10.4* (ns) | *86%* (ns) | *6.9* (ns) | *5.1* (ns) |
| REGION IV-A | Constant TA | 0.83 (***) | 24,458 (**) | 21.7 (**) | 86% (ns) | 6.0 (ns) | 4.7 (ns) |
| REGION IX | Constant TA | 0.81 (***) | 7,641 (**) | 22.3 (**) | 71% (ns) | 5.4 (ns) | 4.1 (ns) |
| REGION V | Constant TA | 0.86 (***) | 2,446 (**) | 21.9 (**) | 86% (ns) | 5.7 (ns) | 4.6 (ns) |
| REGION VI | Continuous TA | 1.00 (***) | 9,655 (ns) | 11.3 (ns) | 100% (ns) | 7.6 (**) | 5.8 (**) |
| REGION VII | Constant TA | 0.73 (***) | 10,337 (*) | 20.9 (*) | 43% (ns) | 3.4 (ns) | 2.6 (ns) |
| REGION VIII | Constant TA | 0.65 (***) | 8,120 (**) | 16.9 (**) | 86% (ns) | 5.3 (ns) | 4.4 (ns) |
| REGION X | Continuous TA | 0.97 (***) | 4,311 (ns) | 6.6 (ns) | 86% (ns) | 6.1 (ns) | 5.1 (*) |
| REGION XI | Constant TA | 0.73 (***) | 7,585 (**) | 21.1 (**) | 57% (ns) | 4.3 (ns) | 3.3 (ns) |
| REGION XII | Constant TA | 0.77 (***) | 8,650 (*) | 23.1 (ns) | 71% (ns) | 5.4 (ns) | 4.1 (ns) |
| REGION XIII | Constant TA | 1.00 (***) | 4,377 (**) | 15.0 (**) | 57% (ns) | 4.6 (ns) | 3.4 (ns) |

**c Per-detector regional dominance probabilities**

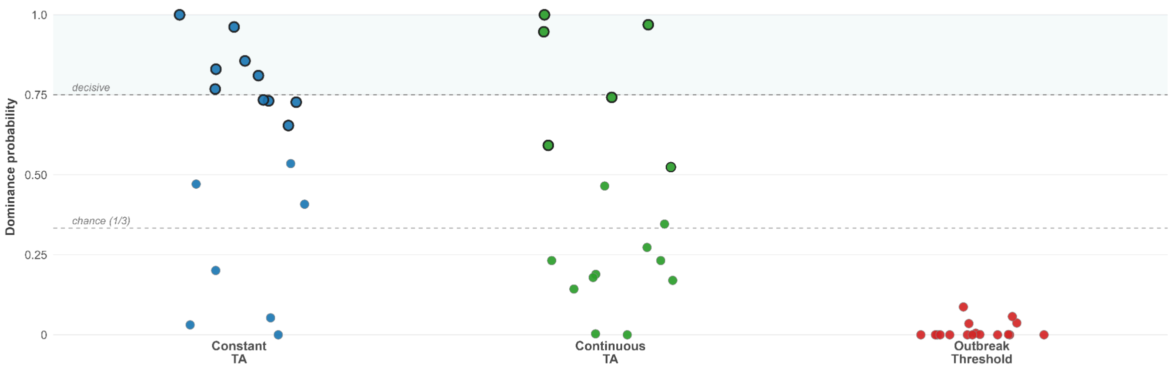


Performance of three target detectors (constant transmission acceleration, continuous transmission acceleration and the outbreak threshold) across 17 regions. a, Map of the regional consensus winner. Each region is filled by its winning detector (constant transmission acceleration, blue; continuous transmission acceleration, green; contested, grey). Fill saturation encodes the bootstrap dominance probability Pr: solid for $Pr \geq 0.90$, mid shade for $0.75 \leq Pr < 0.90$ and light shade for $Pr < 0.75$. Border style encodes the consensus tier, with a solid border for a strong winner, a dashed border for a partial winner and grey fill for a contested region. b, Table of the bootstrap-winning detector and its per-metric significance for each region (n = 17 rows), reporting true-alarm magnitude, number of true alarms, sensitivity, mean lead time and warning persistence; the significance column gives the weakest-link pairwise-adjusted P value. c, Per-detector regional dominance probabilities (51 points, being 17 regions × 3 detectors); the crossbar is the median per detector and the shaded band marks the decisive zone $Pr \geq 0.75$. Of the 17 regions, 15 reached a strong consensus (10 constant transmission acceleration, 5 continuous transmission acceleration), Region III was partial and CAR was contested. Centre values are bootstrap point estimates and intervals derive from B = 1,000 year-cluster resamples. See Supplementary Tables 10, 13 and 14.

**Fig. 6 | Bootstrap-supported country-level dominance and consensus of outbreak detectors across eight dengue-endemic countries.**

**a Bootstrap-winning detector and its per-metric significance, by country**

| Country | Detector | Dominance probability | Sig. | TAM | N true alarms | Sensitivity | Mean lead time (wk) | WP (wk) |
|---|---|---|---|---|---|---|---|---|
| BRAZIL | Constant TA | 1.00 | *** | 1,022,892 (**) | 16.5 (**) | 83% (ns) | 6.7 (ns) | 5.0 (ns) |
| COLOMBIA | Outbreak Threshold | 0.66 | *** | 25,689 (ns) | 11.4 (ns) | 60% (ns) | 4.2 (ns) | 3.4 (ns) |
| MEXICO | Continuous TA (partial) | *0.50* | *** | *69,176 (ns)* | *8.8 (ns)* | *100% (ns)* | *6.7 (ns)* | *5.3 (ns)* |
| PERU | Constant TA | 0.94 | *** | 54,435 (ns) | 15.8 (ns) | 67% (ns) | 5.3 (ns) | 4.0 (ns) |
| PHILIPPINES | Continuous TA | 0.87 | *** | 79,697 (ns) | 10.7 (ns) | 83% (ns) | 6.3 (ns) | 4.8 (ns) |
| SINGAPORE | Constant TA | 0.98 | *** | 7,413 (**) | 19.4 (**) | 71% (ns) | 5.1 (ns) | 4.0 (ns) |
| SRI LANKA | Constant TA | 1.00 | *** | 43,229 (**) | 21.1 (**) | 57% (ns) | 4.6 (ns) | 3.4 (ns) |
| TAIWAN | Continuous TA | 0.70 | *** | 3,425 (ns) | 8.4 (ns) | 60% (ns) | 4.4 (ns) | 3.4 (ns) |

**b Per-detector country dominance probabilities**

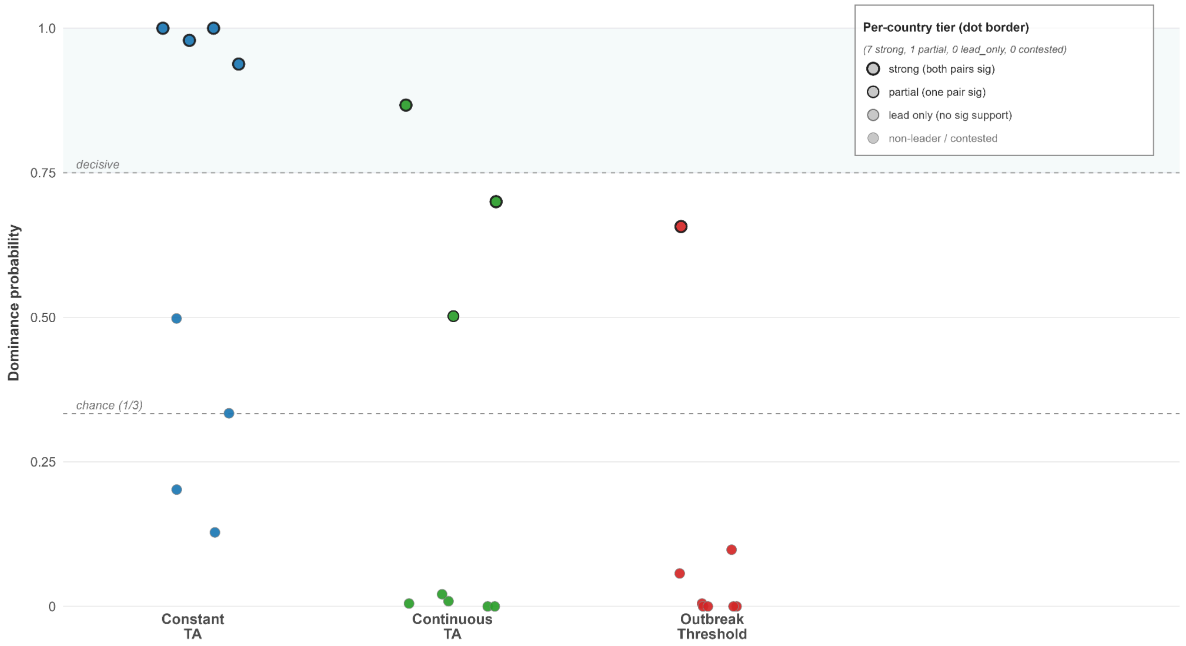


Performance of three target detectors (constant transmission acceleration, continuous transmission acceleration and the outbreak threshold) across eight dengue-endemic countries. a, Table of the bootstrap-winning detector and its per-metric significance by country (n = 8 rows, alphabetical), reporting true-alarm magnitude, number of true alarms, sensitivity, mean lead time and warning persistence. Detector cells are coloured by consensus winner (full colour for a strong winner, faded for a partial winner, grey for a lead-only or contested result); the significance column encodes the weakest-link pairwise-adjusted P value from one asterisk to three. b, Per-detector country dominance probabilities (24 points, being 8 countries × 3 detectors); the crossbar is the median across countries and the shaded band marks the decisive zone Pr ≥ 0.75. Point border thickness encodes the consensus tier, with a thick black border for strong, a medium black border for partial and a grey border for lead-only or non-winning. All eight countries returned an all-pairs consensus winner: constant transmission acceleration in four (Brazil, Peru, Singapore and Sri Lanka), continuous transmission acceleration in three (the Philippines and Taiwan at the strong tier and Mexico at the partial tier) and the outbreak threshold in one (Colombia, Pr = 0.66). Centre values are bootstrap point estimates and Pr derives from B = 1,000 country-cluster resamples. See Supplementary Tables 18 and 19.

#### Tables

Supplementary Table 1 | Headline performance summary across 11 dengue outbreak-detection methods, Quezon City, Philippines, 2013–2024.

| | *Epidemic burden & alarm accuracy* | | | | *Early-warning timeliness* | | | *False Alarms* | |
|---|---|---|---|---|---|---|---|---|---|
| **Detector** | **TAM (cases/yr)** | **True alarms (n yr$^{-1}$)** | **PPV** | **Sensitivity** | **Mean lead time (wks)** | **Warning persistence (wks)** | **ALY** | **False alarms (n yr$^{-1}$)** | **Years w/ true alarm** |
| Constant Transmission Acceleration | **4,257** [3,284, 5,252] | **21.9** [20.5, 23.6] | **83.3%** [76.9%, 90.0%] | **100.0%** [100.0%, 100.0%] | **6.9** [5.9, 7.7] | **5.5** [5.0, 5.8] | **0.18** [0.13, 0.22] | **4.4** [2.6, 6.3] | **10 / 10** |
| Continuous Transmission Acceleration | **2,076** [1,379, 2,763] | **9.8** [8.2, 11.1] | **81.7%** [70.9%, 92.0%] | **100.0%** [100.0%, 100.0%] | **7.6** [7.2, 8.0] | **6.2** [5.8, 6.7] | **0.38** [0.30, 0.44] | **2.2** [0.9, 3.8] | **10 / 10** |
| Incidence Gradient | **2,063** [1,387, 2,762] | **8.8** [7.6, 9.9] | **69.3%** [59.5%, 80.0%] | **100.0%** [100.0%, 100.0%] | **7.5** [6.6, 8.0] | **6.2** [5.5, 6.8] | **0.36** [0.26, 0.46] | **3.9** [2.0, 6.3] | **10 / 10** |
| Hydrological Inflection Measure | **1,494** [675, 2,387] | **4.9** [2.6, 7.4] | **70.0%** [61.9%, 78.5%] | **60.0%** [30.0%, 90.0%] | **4.5** [2.3, 6.8] | **3.6** [1.8, 5.5] | **0.26** [0.11, 0.45] | **2.1** [1.0, 3.3] | **6 / 10** |
| Critical Transition Indicator | **1,573** [1,084, 2,152] | **8.3** [6.6, 9.9] | **75.5%** [61.4%, 90.1%] | **80.0%** [50.0%, 100.0%] | **5.9** [3.8, 7.6] | **4.8** [3.0, 6.2] | **0.38** [0.22, 0.53] | **2.7** [1.0, 4.5] | **8 / 10** |
| Cumulative Sum Control | **2,314** [910, 4,023] | **8.8** [3.4, 15.2] | **50.9%** [43.5%, 62.4%] | **40.0%** [10.0%, 70.0%] | **2.8** [0.8, 5.1] | **2.4** [0.6, 4.3] | **0.06** [0.01, 0.10] | **8.5** [2.6, 15.7] | **4 / 10** |
| Composite Outbreak Signal | **1,119** [343, 2,021] | **3.5** [1.3, 6.0] | **76.1%** [56.0%, 90.6%] | **30.0%** [0.0%, 60.0%] | **1.6** [0.0, 3.2] | **1.6** [0.0, 3.1] | **0.07** [0.00, 0.15] | **1.1** [0.3, 2.0] | **3 / 10** |
| Outbreak Threshold | **1,119** [409, 1,970] | **3.5** [1.5, 6.0] | **49.3%** [26.7%, 80.0%] | **30.0%** [10.0%, 60.0%] | **1.6** [0.5, 3.2] | **1.6** [0.5, 3.0] | **0.07** [0.01, 0.15] | **3.6** [0.8, 6.9] | **3 / 10** |
| Alarm Threshold | **2,163** [887, 3,646] | **7.6** [3.5, 12.0] | **52.1%** [40.1%, 68.8%] | **50.0%** [20.0%, 80.0%] | **3.3** [1.1, 5.4] | **2.8** [1.0, 4.6] | **0.18** [0.03, 0.38] | **7.0** [2.4, 12.4] | **5 / 10** |
| WHO 75th Percentile Threshold | **2,719** [1,202, 4,248] | **10.5** [5.4, 15.3] | **53.3%** [47.0%, 62.0%] | **60.0%** [30.0%, 90.0%] | **4.5** [2.2, 6.7] | **3.6** [1.7, 5.4] | **0.19** [0.05, 0.38] | **9.2** [4.4, 14.2] | **6 / 10** |
| WHO 90th Percentile Threshold | **2,138** [848, 3,557] | **7.6** [3.6, 11.9] | **52.1%** [38.5%, 67.8%] | **60.0%** [30.0%, 90.0%] | **3.9** [1.8, 5.9] | **3.4** [1.7, 5.2] | **0.19** [0.05, 0.38] | **7.0** [2.8, 12.4] | **6 / 10** |

Each cell gives the point estimate (bold) above its 95% bias-corrected bootstrap confidence interval (in brackets), computed from B = 2,000 year-cluster resamples across the 10 evaluable seasons. The eight metrics match those in the dominance matrix in Fig. 2a. TAM, true-alarm magnitude (cases captured by true alarms, summed across the season); true alarms per year and false alarms per year, the season-averaged counts; PPV, positive predictive value; sensitivity, the proportion of evaluable seasons with at least one true actionable alarm; mean lead time, mean weeks between the first true actionable alarm and the peak; warning persistence, mean run length of consecutive true-alarm weeks; ALY, actionable lead-time yield. "Years with true alarm" gives the numerator and denominator of sensitivity. A missing observation is shown as an em dash (—).

Supplementary Table 2 | True-alarm magnitude and mean lead time by detector and season.

| Detector | 2013 | 2014 | 2015 | 2016 | 2017 | 2018 | 2019 | 2022 | 2023 | 2024 |
|---|---|---|---|---|---|---|---|---|---|---|
| Constant Transmission Acceleration | **3,794**<br>(4.0 wk) | **1,748**<br>(8.0 wk) | **4,716**<br>(8.0 wk) | **3,005**<br>(7.0 wk) | **5,069**<br>(7.0 wk) | **5,347**<br>(5.0 wk) | **7,181**<br>(6.0 wk) | **2,742**<br>(8.0 wk) | **2,824**<br>(8.0 wk) | **6,144**<br>(8.0 wk) |
| Continuous Transmission Acceleration | **2,116**<br>(7.0 wk) | **913**<br>(8.0 wk) | **3,139**<br>(8.0 wk) | **1,539**<br>(8.0 wk) | **2,768**<br>(8.0 wk) | **2,207**<br>(6.0 wk) | **4,504**<br>(8.0 wk) | **1,446**<br>(8.0 wk) | **368**<br>(7.0 wk) | **1,762**<br>(8.0 wk) |
| Incidence Gradient | **1,126**<br>(4.0 wk) | **510**<br>(8.0 wk) | **2,818**<br>(8.0 wk) | **1,159**<br>(8.0 wk) | **2,582**<br>(8.0 wk) | **2,327**<br>(7.0 wk) | **4,088**<br>(8.0 wk) | **1,112**<br>(8.0 wk) | **1,200**<br>(8.0 wk) | **3,708**<br>(8.0 wk) |
| Hydrological Inflection Measure | **872**<br>— | **96**<br>— | **1,912**<br>(7.0 wk) | **0**<br>— | **2,038**<br>(8.0 wk) | **2,150**<br>(8.0 wk) | **3,758**<br>(7.0 wk) | **273**<br>(7.0 wk) | **136**<br>— | **3,708**<br>(8.0 wk) |
| Critical Transition Indicator | **1,867**<br>(7.0 wk) | **645**<br>(8.0 wk) | **1,072**<br>(8.0 wk) | **1,381**<br>(8.0 wk) | **2,950**<br>(8.0 wk) | **2,207**<br>(6.0 wk) | **3,216**<br>(6.0 wk) | **929**<br>(8.0 wk) | **723**<br>— | **736**<br>— |
| Cumulative Sum Control | **519**<br>— | **0**<br>— | **2,303**<br>— | **0**<br>— | **3,654**<br>(8.0 wk) | **5,763**<br>(8.0 wk) | **7,126**<br>(7.0 wk) | **0**<br>— | **0**<br>— | **3,770**<br>(5.0 wk) |
| Composite Outbreak Signal | **257**<br>— | **0**<br>— | **2,706**<br>— | **0**<br>— | **878**<br>— | **464**<br>(6.0 wk) | **3,367**<br>(5.0 wk) | **0**<br>— | **0**<br>— | **3,517**<br>(5.0 wk) |
| Outbreak Threshold | **257**<br>— | **0**<br>— | **2,706**<br>— | **0**<br>— | **878**<br>— | **464**<br>(6.0 wk) | **3,367**<br>(5.0 wk) | **0**<br>— | **0**<br>— | **3,517**<br>(5.0 wk) |
| Alarm Threshold | **1,099**<br>— | **0**<br>— | **3,010**<br>— | **0**<br>— | **2,803**<br>(4.0 wk) | **2,745**<br>(7.0 wk) | **6,891**<br>(7.0 wk) | **114**<br>(7.0 wk) | **0**<br>— | **4,970**<br>(8.0 wk) |
| WHO 75th Percentile Threshold | **1,477**<br>— | **96**<br>— | **3,810**<br>(7.0 wk) | **477**<br>— | **3,872**<br>(8.0 wk) | **4,015**<br>(8.0 wk) | **6,891**<br>(7.0 wk) | **273**<br>(7.0 wk) | **136**<br>— | **6,144**<br>(8.0 wk) |
| WHO 90th Percentile Threshold | **1,099**<br>— | **96**<br>— | **2,982**<br>(6.0 wk) | **0**<br>— | **2,803**<br>(4.0 wk) | **2,425**<br>(7.0 wk) | **6,891**<br>(7.0 wk) | **114**<br>(7.0 wk) | **0**<br>— | **4,970**<br>(8.0 wk) |

Each cell stacks the season-level true-alarm magnitude (cases captured, top, bold) above the mean lead time of the first true actionable alarm in parentheses (weeks before the peak). Values are season-level point estimates with no interval, as each cell is a single season; n = 10 seasons. The pandemic seasons 2020 and 2021 and the partial 2025 season are excluded from the primary specification (see Supplementary Table 3). A zero magnitude with a dash for lead time means the detector fired no alarm that season.

Supplementary Table 3 | Sensitivity of headline metrics to year-inclusion rules, by detector.

| Specification | Detector | True-alarm magnitude (cases) | Number of true alarms | Positive Predictive Value (%) | Sensitivity (%) | Mean lead (wk) | Warning persistence (wk) | Actionable lead-time yield | Number of false alarms | Number of evaluable years |
|---|---|---|---|---|---|---|---|---|---|---|
| **Primary (10 yr)** | **Constant Transmission Acceleration** | 4,257 | 21.9 | 83.3% | 100.0% | 6.9 | 5.5 | 0.18 | 4.4 | 10 |
| | **Continuous Transmission Acceleration** | 2,076 | 9.8 | 81.7% | 100.0% | 7.6 | 6.2 | 0.38 | 2.2 | 10 |
| | **Incidence Gradient** | 2,063 | 8.8 | 69.3% | 100.0% | 7.5 | 6.2 | 0.36 | 3.9 | 10 |
| | **Hydrological Inflection Measure** | 1,494 | 4.9 | 70.0% | 60.0% | 4.5 | 3.6 | 0.26 | 2.1 | 10 |
| | **Critical Transition Indicator** | 1,573 | 8.3 | 75.5% | 80.0% | 5.9 | 4.8 | 0.38 | 2.7 | 10 |
| | **Cumulative Sum Control** | 2,314 | 8.8 | 50.9% | 40.0% | 2.8 | 2.4 | 0.06 | 8.5 | 10 |
| | **Composite Outbreak Signal** | 1,119 | 3.5 | 76.1% | 30.0% | 1.6 | 1.6 | 0.07 | 1.1 | 10 |
| | **Outbreak Threshold** | 1,119 | 3.5 | 49.3% | 30.0% | 1.6 | 1.6 | 0.07 | 3.6 | 10 |
| | **Alarm Threshold** | 2,163 | 7.6 | 52.1% | 50.0% | 3.3 | 2.8 | 0.18 | 7.0 | 10 |
| | **WHO 75th Percentile Threshold** | 2,719 | 10.5 | 53.3% | 60.0% | 4.5 | 3.6 | 0.19 | 9.2 | 10 |
| | **WHO 90th Percentile Threshold** | 2,138 | 7.6 | 52.1% | 60.0% | 3.9 | 3.4 | 0.19 | 7.0 | 10 |
| *+2020/2021 (12 yr)* | Constant Transmission Acceleration | 3,613 | 20.5 | 76.6% | 91.7% | 6.4 | 5.0 | 0.17 | 6.3 | 12 |
| | Continuous Transmission Acceleration | 1,745 | 8.7 | 71.7% | 91.7% | 7.0 | 5.8 | 0.33 | 3.4 | 12 |
| | Incidence Gradient | 1,719 | 7.3 | 69.3% | 83.3% | 6.2 | 5.1 | 0.30 | 3.3 | 12 |
| | Hydrological Inflection Measure | 1,261 | 4.2 | 70.4% | 50.0% | 3.8 | 3.0 | 0.22 | 1.8 | 12 |
| | Critical Transition Indicator | 1,324 | 7.3 | 67.2% | 75.0% | 5.6 | 4.6 | 0.33 | 3.6 | 12 |
| | Cumulative Sum Control | 1,928 | 7.3 | 50.9% | 33.3% | 2.3 | 2.0 | 0.05 | 7.1 | 12 |
| | Composite Outbreak Signal | 932 | 2.9 | 76.1% | 25.0% | 1.3 | 1.3 | 0.06 | 0.9 | 12 |
| | Outbreak Threshold | 932 | 2.9 | 49.3% | 25.0% | 1.3 | 1.3 | 0.06 | 3.0 | 12 |
| | Alarm Threshold | 1,803 | 6.3 | 52.1% | 41.7% | 2.8 | 2.4 | 0.15 | 5.8 | 12 |

| Specification | Detector | True-alarm magnitude (cases) | Number of true alarms | Positive Predictive Value (%) | Sensitivity (%) | Mean lead (wk) | Warning persistence (wk) | Actionable lead-time yield | Number of false alarms | Number of evaluable years |
|---|---|---|---|---|---|---|---|---|---|---|
| | WHO 75th Percentile Threshold | 2,281 | 8.8 | 53.5% | 50.0% | 3.8 | 3.0 | 0.16 | 7.7 | 12 |
| | WHO 90th Percentile Threshold | 1,782 | 6.3 | 52.1% | 50.0% | 3.2 | 2.9 | 0.16 | 5.8 | 12 |
| *+2025 (11 yr)* | Constant Transmission Acceleration | 4,072 | 20.5 | 80.1% | 90.9% | 6.3 | 5.0 | 0.16 | 5.1 | 11 |
| | Continuous Transmission Acceleration | 2,170 | 9.9 | 83.2% | 90.9% | 6.9 | 5.6 | 0.34 | 2.0 | 11 |
| | Incidence Gradient | 2,231 | 9.3 | 70.8% | 100.0% | 7.4 | 6.1 | 0.34 | 3.8 | 11 |
| | Hydrological Inflection Measure | 1,747 | 5.9 | 73.0% | 63.6% | 4.6 | 3.8 | 0.25 | 2.2 | 11 |
| | Critical Transition Indicator | 1,769 | 8.7 | 76.2% | 81.8% | 6.0 | 4.9 | 0.36 | 2.7 | 11 |
| | Cumulative Sum Control | 2,809 | 11.3 | 55.4% | 45.5% | 3.1 | 2.6 | 0.06 | 9.1 | 11 |
| | Composite Outbreak Signal | 1,357 | 4.4 | 78.7% | 36.4% | 2.1 | 2.0 | 0.08 | 1.2 | 11 |
| | Outbreak Threshold | 1,508 | 5.3 | 60.4% | 36.4% | 2.1 | 1.9 | 0.08 | 3.5 | 11 |
| | Alarm Threshold | 2,662 | 10.1 | 59.7% | 54.5% | 3.6 | 3.1 | 0.17 | 6.8 | 11 |
| | WHO 75th Percentile Threshold | 3,196 | 12.9 | 57.0% | 63.6% | 4.7 | 3.8 | 0.18 | 9.7 | 11 |
| | WHO 90th Percentile Threshold | 2,634 | 10.1 | 59.7% | 63.6% | 4.2 | 3.6 | 0.18 | 6.8 | 11 |
| *All (13 yr)* | Constant Transmission Acceleration | 3,506 | 19.5 | 74.4% | 84.6% | 5.9 | 4.7 | 0.15 | 6.7 | 13 |
| | Continuous Transmission Acceleration | 1,850 | 8.8 | 73.7% | 84.6% | 6.5 | 5.3 | 0.30 | 3.2 | 13 |
| | Incidence Gradient | 1,887 | 7.8 | 70.8% | 84.6% | 6.2 | 5.1 | 0.29 | 3.2 | 13 |
| | Hydrological Inflection Measure | 1,492 | 5.1 | 73.3% | 53.8% | 3.9 | 3.2 | 0.21 | 1.8 | 13 |
| | Critical Transition Indicator | 1,510 | 7.8 | 68.7% | 76.9% | 5.7 | 4.7 | 0.32 | 3.5 | 13 |
| | Cumulative Sum Control | 2,377 | 9.5 | 55.4% | 38.5% | 2.6 | 2.2 | 0.05 | 7.7 | 13 |
| | Composite Outbreak Signal | 1,148 | 3.7 | 78.7% | 30.8% | 1.8 | 1.7 | 0.07 | 1.0 | 13 |
| | Outbreak Threshold | 1,276 | 4.5 | 60.4% | 30.8% | 1.8 | 1.6 | 0.07 | 2.9 | 13 |
| | Alarm Threshold | 2,252 | 8.5 | 59.7% | 46.2% | 3.1 | 2.6 | 0.15 | 5.8 | 13 |

| Specification | Detector | True-alarm magnitude (cases) | Number of true alarms | Positive Predictive Value (%) | Sensitivity (%) | Mean lead (wk) | Warning persistence (wk) | Actionable lead-time yield | Number of false alarms | Number of evaluable years |
|---|---|---|---|---|---|---|---|---|---|---|
| | WHO 75th Percentile Threshold | 2,719 | 11.0 | 57.2% | 53.8% | 4.0 | 3.2 | 0.15 | 8.2 | 13 |
| | WHO 90th Percentile Threshold | 2,229 | 8.5 | 59.7% | 53.8% | 3.5 | 3.1 | 0.16 | 5.8 | 13 |

The primary specification (detector name in bold) excludes the pandemic seasons 2020 and 2021 and the partial 2025 season. Three further specifications add subsets of those years back to the evaluable set. Point estimates only are tabulated for compactness; the paired bootstrap confidence intervals for the primary specification are in Supplementary Table 1. The number of evaluable years per specification is given in the last column. Stable detector ranking across specifications supports the primary choice.

Supplementary Table 4 | Constant transmission acceleration versus the outbreak threshold pooled across 10 evaluable seasons, Quezon City, 2013–2024.

| Detector | Total triggers (n) | True alarms (n) | False alarms (n) | Positive Predictive Value (%) | Sensitivity (%) | Years with True Alarm (AW) | Years with any true alarm in AW ∪ EB | Mean lead time (wks) | Actionable lead-time yield (%) | Warning persistence (wks) |
|---|---|---|---|---|---|---|---|---|---|---|
| **Constant TA** | 263 | 219 | 44 | 83.3% | 100.0% | 10 / 10 | *10 / 10* | 6.9 | 18.1% | 5.45 |
| **Outbreak threshold** | 71 | 35 | 36 | 49.3% | 30.0% | 3 / 10 | *6 / 10* | 1.6 | 9.8% | 2.21 |

Performance pooled across the 10 evaluable seasons (2013 to 2019 and 2022 to 2024; 2020, 2021 and 2025 excluded). Values are pooled point estimates; n = 10 seasons. A trigger is a true alarm if it falls within the actionable window (peak−8 to peak−4 weeks) or within the 70% epidemic-burden block, and a false alarm otherwise. PPV is true alarms divided by total triggers. The headline sensitivity is the proportion of seasons with at least one true alarm in the actionable window; the relaxed column counting any true alarm in the actionable window or epidemic-burden block is given for context only. Mean lead time is bounded to 4 to 8 weeks by construction. ALY is true actionable triggers divided by total true alarms; warning persistence is the mean across seasons of the run length of consecutive alarm-on weeks.

Supplementary Table 5 | Per-year detector performance with lead-time compartment metrics, 2013–2024.

| Year | Detector | Peak (wk) | Actionable Window | Epidemic Week Coverage | Total triggers (n) | True alarms (n) | False alarms (n) | Positive Predictive Value (%) | First true alarm week | Lead time (wks) | Compartment | True-Alarm Magnitude | Actionable lead-time yield (%) | Warning persistence (wks) |
|---|---|---|---|---|---|---|---|---|---|---|---|---|---|---|
| **2013** | Constant TA | 30 | Wk 22–26 | Wk 25–45 | 27 | 20 | 7 | 74.1% | Wk 26 | 4 | Actionable | 3794 | 5.0% | 4.0 |
| | Outbreak threshold | 30 | Wk 22–26 | Wk 25–45 | 1 | 1 | 0 | 100.0% | — | — | — | 257 | 0.0% | — |
| **2014** | Constant TA | 39 | Wk 31–35 | Wk 25–49 | 25 | 22 | 3 | 88.0% | Wk 31 | 8 | Actionable | 1748 | 22.7% | 6.0 |

| | | | | | | | | | | | | | | |
|---|---|---|---|---|---|---|---|---|---|---|---|---|---|---|
| | Outbreak threshold | 39 | Wk 31–35 | Wk 25–49 | 0 | 0 | 0 | — | — | — | — | 0 | — | — |
| **2015** | Constant TA | 38 | Wk 30–34 | Wk 29–46 | 25 | 18 | 7 | 72.0% | Wk 30 | 8 | Actionable | 4716 | 27.8% | 6.0 |
| | Outbreak threshold | 38 | Wk 30–34 | Wk 29–46 | 13 | 10 | 3 | 76.9% | — | — | — | 2706 | 0.0% | — |
| **2016** | Constant TA | 35 | Wk 27–31 | Wk 26–49 | 25 | 22 | 3 | 88.0% | Wk 28 | 7 | Actionable | 3005 | 18.2% | 5.5 |
| | Outbreak threshold | 35 | Wk 27–31 | Wk 26–49 | 2 | 0 | 2 | 0.0% | — | — | — | 0 | — | — |
| **2017** | Constant TA | 33 | Wk 25–29 | Wk 24–48 | 27 | 23 | 4 | 85.2% | Wk 26 | 7 | Actionable | 5069 | 17.4% | 5.5 |
| | Outbreak threshold | 33 | Wk 25–29 | Wk 24–48 | 14 | 3 | 11 | 21.4% | — | — | — | 878 | 0.0% | — |
| **2018** | Constant TA | 32 | Wk 24–28 | Wk 24–51 | 26 | 25 | 1 | 96.2% | Wk 27 | 5 | Actionable | 5347 | 8.0% | 4.5 |
| | Outbreak threshold | 32 | Wk 24–28 | Wk 24–51 | 17 | 3 | 14 | 17.6% | Wk 26 | 6 | Actionable | 464 | 33.3% | 6.0 |
| **2019** | Constant TA | 35 | Wk 27–31 | Wk 29–49 | 24 | 21 | 3 | 87.5% | Wk 29 | 6 | Actionable | 7181 | 14.3% | 5.0 |
| | Outbreak threshold | 35 | Wk 27–31 | Wk 29–49 | 14 | 8 | 6 | 57.1% | Wk 30 | 5 | Actionable | 3367 | 25.0% | 4.5 |
| **2022** | Constant TA | 32 | Wk 24–28 | Wk 24–44 | 32 | 21 | 11 | 65.6% | Wk 24 | 8 | Actionable | 2742 | 23.8% | 6.0 |
| | Outbreak threshold | 32 | Wk 24–28 | Wk 24–44 | 0 | 0 | 0 | — | — | — | — | 0 | — | — |
| **2023** | Constant TA | 41 | Wk 33–37 | Wk 26–52 | 27 | 27 | 0 | 100.0% | Wk 33 | 8 | Actionable | 2824 | 18.5% | 6.0 |
| | Outbreak threshold | 41 | Wk 33–37 | Wk 26–52 | 0 | 0 | 0 | — | — | — | — | 0 | — | — |
| **2024** | Constant TA | 47 | Wk 39–43 | Wk 33–52 | 25 | 20 | 5 | 80.0% | Wk 39 | 8 | Actionable | 6144 | 25.0% | 6.0 |
| | Outbreak threshold | 47 | Wk 39–43 | Wk 33–52 | 10 | 10 | 0 | 100.0% | Wk 42 | 5 | Actionable | 3517 | 10.0% | 5.0 |

One row per season and detector. Values are single-season point estimates with no interval (n = 10 seasons × 3 detectors = 30 rows). The two anchor rules (actionable window and 70% epidemic-burden block) are shared across detectors within a season. PPV per year is true alarms divided by total triggers; a dash marks no qualifying observation. Lead time is computed for the first true alarm in the actionable window and is bounded to 4 to 8 weeks by construction. The compartment column classifies that first alarm as actionable (peak−8 to peak−4 weeks) or reactive (at or after the peak).

Supplementary Table 6 | Head-to-head paired Wilcoxon comparisons across operational performance metrics.

| Metric | Comparison | *n* yrs | Median paired Δ (95% CI) | *V* | *P* (Wilcoxon, two-sided) |
|---|---|---|---|---|---|
| **Epidemic Burden and Alarm Accuracy** | | | | | |
| True-alarm magnitude (cases) | **Constant TA vs OT** | **10** | **2,914.5 (2,376, 4,002.5)** | **55.0** | **0.0059 *** |
| | Continuous TA vs OT | 10 | 1,291.5 (433, 1,743) | 47.0 | 0.053 |
| | Constant TA vs Continuous TA | 10 | 1,989.5 (1,436.5, 2,722.4) | 55.0 | 0.0059 * |
| Number of true alarms | **Constant TA vs OT** | **10** | **20.5 (13, 22)** | **55.0** | **0.0058 *** |
| | Continuous TA vs OT | 10 | 7.5 (2.5, 10.5) | 52.5 | 0.012 * |
| | Constant TA vs Continuous TA | 10 | 11.0 (9.5, 13.5) | 55.0 | 0.0058 * |
| Positive predictive value | **Constant TA vs OT** | **7** | **0.304 (-0.200, 0.786)** | **22.0** | **0.205** |
| | Continuous TA vs OT | 7 | 0.275 (-0.122, 0.824) | 17.0 | 0.208 |
| | Constant TA vs Continuous TA | 10 | -0.038 (-0.113, 0.132) | 25.0 | 0.839 |
| Sensitivity | **Constant TA vs OT** | **10** | **1.000 (0.000, 1.000)** | **28.0** | **0.011 *** |
| | Continuous TA vs OT | 10 | 1.000 (0.000, 1.000) | 28.0 | 0.011 * |
| | Constant TA vs Continuous TA | 10 | 0.000 (0.000, 0.000) | 0.0 | n.a. |
| **Early-Warning Timeliness** | | | | | |
| Mean lead time (wks) | **Constant TA vs OT** | **10** | **7.0 (2.0, 8.0)** | **53.5** | **0.0088 *** |
| | Continuous TA vs OT | 10 | 7.5 (3.0, 8.0) | 45.0 | 0.0079 * |
| | Constant TA vs Continuous TA | 10 | -0.5 (-1.5, 0.0) | 2.5 | 0.106 |
| Warning persistence (wks) | **Constant TA vs OT** | **10** | **5.5 (0.75, 6.0)** | **52.0** | **0.014 *** |
| | Continuous TA vs OT | 10 | 6.0 (2.25, 6.0) | 54.0 | 0.0072 * |
| | Constant TA vs Continuous TA | 10 | -0.5 (-1.25, 0.0) | 0.0 | 0.021 * |
| Actionable lead-time yield | **Constant TA vs OT** | **10** | **0.178 (-0.029, 0.227)** | **44.0** | **0.103** |
| | Continuous TA vs OT | 10 | 0.390 (0.202, 0.455) | 45.0 | 0.0090 * |
| | Constant TA vs Continuous TA | 10 | -0.248 (-0.283, -0.128) | 2.0 | 0.011 * |
| **False Alarms** | | | | | |
| Number of false alarms | **Constant TA vs OT** | **10** | **2.0 (-5, 5)** | **27.0** | **0.635** |
| | Continuous TA vs OT | 10 | 0.0 (-7, 3) | 15.0 | 0.726 |
| | Constant TA vs Continuous TA | 10 | 1.5 (-0.5, 4.5) | 45.0 | 0.081 |

Comparisons are paired by season. For each metric and pair the table reports the median paired difference (Detector A minus Detector B) with its 95% confidence interval from B = 1,000 year-cluster bootstrap resamples, the Wilcoxon signed-rank statistic V, and the two-sided P value. Three pairwise contrasts are reported per metric (constant transmission acceleration versus the outbreak threshold; continuous transmission acceleration versus the outbreak threshold; constant versus continuous transmission acceleration). n is 10 seasons except for positive predictive value, where two seasons have no triggers in either arm and n falls to 7. Each contrast is tested at alpha = 0.05; an asterisk marks P < 0.05. The bootstrap confidence intervals are the primary effect-size statement and the Wilcoxon P values are a sensitivity diagnostic.

Supplementary Table 7 | Sensitivity of headline metrics to the actionable-window width.

| AW specification (wks pre-peak) | Detector | Total triggers (n) | True alarms (n) | False alarms (n) | Positive Predictive Value | Sensitivity | Mean lead time (wks) |
|---|---|---|---|---|---|---|---|
| 3-6 weeks pre-peak | Constant TA | 263 | 219 | 44 | 83.3% | 100.0% | 5.7 |
| 3-6 weeks pre-peak | Outbreak threshold | 71 | 35 | 36 | 49.3% | 30.0% | 1.6 |
| **4-8 weeks pre-peak** *(primary)* | Constant TA | 263 | 219 | 44 | 83.3% | 100.0% | 6.9 |
| **4-8 weeks pre-peak** *(primary)* | Outbreak threshold | 71 | 35 | 36 | 49.3% | 30.0% | 1.6 |
| 5-10 weeks pre-peak | Constant TA | 263 | 222 | 41 | 84.4% | 90.0% | 7.5 |
| 5-10 weeks pre-peak | Outbreak threshold | 71 | 35 | 36 | 49.3% | 30.0% | 1.6 |

Table 7 varies the actionable window across narrow (3 to 6 weeks), primary (4 to 8 weeks) and wide (5 to 10 weeks) definitions. Values are point estimates; year inclusion and detector parameters match the primary analysis (n = 10 seasons). Wider windows raise the share of in-window triggers and so raise positive predictive value; narrower ones lower it.

Supplementary Table 8 | Sensitivity to the epidemic-burden cumulative-case threshold.

| A2 burden specification | Detector | Total triggers (n) | True alarms (n) | False alarms (n) | Positive Predictive Value | Sensitivity | Mean lead time (wks) |
|---|---|---|---|---|---|---|---|
| 60% cumulative burden | Constant TA | 263 | 180 | 83 | 68.4% | 100.0% | 6.9 |
| 60% cumulative burden | Outbreak threshold | 71 | 27 | 44 | 38.0% | 30.0% | 1.6 |
| **70% cumulative burden** *(primary)* | Constant TA | 263 | 219 | 44 | 83.3% | 100.0% | 6.9 |
| **70% cumulative burden** *(primary)* | Outbreak threshold | 71 | 35 | 36 | 49.3% | 30.0% | 1.6 |

| | | | | | | | |
|---|---|---|---|---|---|---|---|
| 80% cumulative burden | Constant TA | 263 | 251 | 12 | 95.4% | 100.0% | 6.9 |
| 80% cumulative burden | Outbreak threshold | 71 | 51 | 20 | 71.8% | 30.0% | 1.6 |

Table 8 varies the epidemic-burden envelope across 60%, primary 70% and 80% cumulative cases. Values are point estimates; year inclusion and detector parameters match the primary analysis (n = 10 seasons). Wider envelopes raise the share of in-window triggers and so raise positive predictive value; narrower ones lower it.

Supplementary Table 9 | Sensitivity of headline metrics to year-inclusion rules.

| **Year-inclusion specification** | **Detector** | **Total triggers (n)** | **True alarms (n)** | **False alarms (n)** | **Positive Predictive Value** | **Sensitivity** | **Mean lead time (wks)** |
|---|---|---|---|---|---|---|---|
| **10 seasons (primary; excludes 2020, 2021, 2025)** *(primary)* | Constant TA | 263 | 219 | 44 | 83.3% | 100.0% | 6.9 |
| **10 seasons (primary; excludes 2020, 2021, 2025)** *(primary)* | Outbreak threshold | 71 | 35 | 36 | 49.3% | 30.0% | 1.6 |
| 11 seasons (primary + 2020) | Constant TA | 276 | 219 | 57 | 79.3% | 90.9% | 6.3 |
| 11 seasons (primary + 2020) | Outbreak threshold | 71 | 35 | 36 | 49.3% | 27.3% | 1.4 |
| 11 seasons (primary + 2021) | Constant TA | 308 | 246 | 62 | 79.9% | 100.0% | 7.0 |
| 11 seasons (primary + 2021) | Outbreak threshold | 71 | 35 | 36 | 49.3% | 27.3% | 1.4 |
| 11 seasons (primary + 2025) | Constant TA | 282 | 226 | 56 | 80.1% | 90.9% | 6.3 |
| 11 seasons (primary + 2025) | Outbreak threshold | 96 | 58 | 38 | 60.4% | 36.4% | 2.1 |
| 13 seasons (all available) | Constant TA | 340 | 253 | 87 | 74.4% | 84.6% | 5.9 |
| 13 seasons (all available) | Outbreak threshold | 96 | 58 | 38 | 60.4% | 30.8% | 1.8 |

Each row reports detector performance under the indicated inclusion set. The primary set is 10 seasons (2013 to 2019 and 2022 to 2024); values are point estimates. Adding 2020 lowers sensitivity for all detectors because that season produced no qualifying true alarm for at least one detector under the anchor rule.

Supplementary Table 10 | Per-region operational performance of the 11 outbreak-detection methods across the 17 Philippine regions.

| Region | Detector | TAM | True alarms | Sens. | Lead (wks) | WP (wks) | Yrs w/ TA |
|---|---|---|---|---|---|---|---|
| **BARMM** | **Constant Transmission Acceleration** | **4,037** [2,405, 5,663] | **24.7** [20.7, 27.5] | **83.3%** [33.3%, 83.3%] | **6.50** [3.83, 8.00] | **4.92** [2.92, 6.00] | **5 / 6** |
| | Continuous Transmission Acceleration | 1,132 [779, 1,593] | 7.3 [4.8, 9.8] | 100.0% [50.0%, 83.3%] | 6.67 [5.67, 7.50] | 5.50 [4.91, 6.08] | 6 / 6 |
| | Incidence Gradient | 1,609 [839, 2,269] | 8.8 [4.5, 12] | 83.3% [33.3%, 83.3%] | 6.67 [4.00, 8.00] | 5.29 [3.12, 6.67] | 5 / 6 |
| | Hydrological Inflection Measure | 1,604 [783, 2,338] | 8.7 [4.3, 12.3] | 66.7% [16.7%, 66.7%] | 5.33 [1.33, 8.00] | 4.12 [1.17, 6.29] | 4 / 6 |
| | Critical Transition Indicator | 910 [195, 1,698] | 5 [1.5, 9] | 66.7% [16.7%, 66.7%] | 5.00 [2.33, 7.67] | 4.08 [1.83, 6.25] | 4 / 6 |
| | Cumulative Sum Control | 3,716 [1,830, 5,861] | 21.2 [11.5, 28.3] | 66.7% [16.7%, 66.7%] | 5.33 [1.33, 8.00] | 4.00 [1.00, 6.00] | 4 / 6 |
| | Composite Outbreak Signal | 2,234 [662, 3,943] | 10.5 [4.3, 16.7] | 66.7% [16.7%, 66.7%] | 5.17 [1.33, 7.83] | 4.38 [1.17, 6.67] | 4 / 6 |
| | Outbreak Threshold | 2,342 [662, 4,170] | 11.2 [4.3, 17.7] | 66.7% [16.7%, 66.7%] | 5.17 [1.33, 7.83] | 4.38 [1.17, 6.67] | 4 / 6 |
| | Alarm Threshold | 3,320 [1,405, 5,387] | 17.8 [9.3, 24.5] | 66.7% [16.7%, 66.7%] | 5.33 [1.33, 8.00] | 4.00 [1.00, 6.00] | 4 / 6 |
| | WHO 75th Percentile Threshold | 3,738 [1,779, 5,751] | 20.8 [11.3, 27.3] | 66.7% [16.7%, 66.7%] | 5.33 [1.33, 8.00] | 4.00 [1.00, 6.00] | 4 / 6 |
| | WHO 90th Percentile Threshold | 3,511 [1,530, 5,510] | 19.3 [10.5, 25.8] | 66.7% [16.7%, 66.7%] | 5.33 [1.33, 8.00] | 4.00 [1.00, 6.00] | 4 / 6 |
| **CAR** | Constant Transmission Acceleration | 8,528 [4,403, 13,872] | 18.7 [13.7, 22.4] | 85.7% [28.6%, 71.4%] | 6.14 [3.86, 7.86] | 4.79 [3.07, 5.93] | 6 / 7 |
| | Continuous Transmission Acceleration | 5,538 [2,578, 9,716] | 10.1 [8.6, 11.3] | 85.7% [28.6%, 71.4%] | 6.57 [4.29, 8.00] | 5.00 [3.29, 6.00] | 6 / 7 |
| | Incidence Gradient | 4,708 [1,456, 9,218] | 7.6 [4.6, 9.9] | 85.7% [28.6%, 71.4%] | 5.86 [3.71, 7.43] | 4.79 [3.14, 5.86] | 6 / 7 |
| | Hydrological Inflection Measure | 3,842 [370, 8,693] | 4 [1, 7.4] | 28.6% [0.0%, 28.6%] | 2.29 [0.00, 4.57] | 1.71 [0.00, 3.43] | 2 / 7 |
| | Critical Transition Indicator | 2,532 [1,156, 4,100] | 6.6 [3.9, 9.3] | 71.4% [28.6%, 71.4%] | 5.29 [2.29, 8.00] | 4.07 [1.71, 6.00] | 5 / 7 |
| | Cumulative Sum Control | 5,382 [581, 11,440] | 7 [2.6, 11.9] | 42.9% [14.3%, 42.9%] | 3.14 [0.86, 6.29] | 2.64 [0.71, 5.14] | 3 / 7 |

| Region | Detector | TAM | True alarms | Sens. | Lead (wks) | WP (wks) | Yrs w/ TA |
|---|---|---|---|---|---|---|---|
| | Composite Outbreak Signal | 5,361<br>[559, 11,440] | 7.1<br>[2.6, 11.9] | 28.6%<br>[0.0%, 28.6%] | 2.00<br>[0.00, 4.29] | 1.57<br>[0.00, 3.43] | 2 / 7 |
| | Outbreak Threshold | 5,361<br>[559, 11,440] | 7.1<br>[2.6, 11.9] | 28.6%<br>[0.0%, 28.6%] | 2.00<br>[0.00, 4.29] | 1.57<br>[0.00, 3.43] | 2 / 7 |
| | Alarm Threshold | 5,949<br>[1,031, 12,189] | 8.6<br>[4.4, 12.9] | 28.6%<br>[0.0%, 28.6%] | 2.29<br>[0.00, 4.57] | 1.71<br>[0.00, 3.43] | 2 / 7 |
| | WHO 75th Percentile Threshold | 6,355<br>[1,479, 12,434] | 10.1<br>[5.9, 14.1] | 28.6%<br>[0.0%, 28.6%] | 2.29<br>[0.00, 4.57] | 1.71<br>[0.00, 3.43] | 2 / 7 |
| | WHO 90th Percentile Threshold | 6,088<br>[1,212, 12,327] | 9.1<br>[4.9, 13.3] | 28.6%<br>[0.0%, 28.6%] | 2.29<br>[0.00, 4.57] | 1.71<br>[0.00, 3.43] | 2 / 7 |
| **MIMAROPA** | Constant Transmission Acceleration | 6,527<br>[3,701, 9,376] | 21.6<br>[19.3, 24.1] | 85.7%<br>[28.6%, 85.7%] | 5.57<br>[3.43, 7.29] | 4.50<br>[2.86, 5.64] | 6 / 7 |
| | **Continuous Transmission Acceleration** | **3,682**<br>[2,158, 5,268] | **10**<br>[9, 11] | **100.0%**<br>[42.9%, 85.7%] | **6.86**<br>[6.00, 7.71] | **5.43**<br>[5.00, 5.86] | **7 / 7** |
| | Incidence Gradient | 3,122<br>[1,573, 4,794] | 7.7<br>[4.7, 10] | 85.7%<br>[28.6%, 85.7%] | 6.14<br>[3.85, 8.00] | 4.79<br>[3.07, 6.00] | 6 / 7 |
| | Hydrological Inflection Measure | 3,133<br>[1,522, 4,693] | 7.6<br>[4.4, 10.3] | 85.7%<br>[28.6%, 85.7%] | 6.00<br>[3.71, 7.71] | 4.71<br>[3.00, 5.86] | 6 / 7 |
| | Critical Transition Indicator | 2,386<br>[986, 3,812] | 5.6<br>[3.1, 7.9] | 71.4%<br>[28.6%, 71.4%] | 5.43<br>[2.84, 7.71] | 4.29<br>[2.27, 6.14] | 5 / 7 |
| | Cumulative Sum Control | 5,806<br>[2,635, 8,711] | 15.9<br>[7.4, 23.3] | 57.1%<br>[14.3%, 57.1%] | 4.57<br>[1.14, 6.86] | 3.43<br>[0.86, 5.14] | 4 / 7 |
| | Composite Outbreak Signal | 3,557<br>[1,342, 5,818] | 9.3<br>[4.6, 14.9] | 85.7%<br>[28.6%, 85.7%] | 5.43<br>[3.14, 7.43] | 4.43<br>[2.85, 5.71] | 6 / 7 |
| | Outbreak Threshold | 3,565<br>[1,358, 5,826] | 9.4<br>[4.9, 14.9] | 85.7%<br>[28.6%, 85.7%] | 5.71<br>[3.71, 7.43] | 4.57<br>[3.00, 5.71] | 6 / 7 |
| | Alarm Threshold | 4,774<br>[2,228, 7,297] | 12.7<br>[6.9, 18.4] | 85.7%<br>[28.6%, 85.7%] | 6.00<br>[3.71, 7.71] | 4.71<br>[3.00, 5.86] | 6 / 7 |
| | WHO 75th Percentile Threshold | 5,562<br>[2,560, 8,608] | 14.9<br>[8, 21.1] | 85.7%<br>[28.6%, 85.7%] | 6.29<br>[4.00, 7.71] | 4.86<br>[3.14, 5.86] | 6 / 7 |
| | WHO 90th Percentile Threshold | 5,261<br>[2,436, 8,064] | 13.9<br>[7.4, 19.9] | 85.7%<br>[28.6%, 85.7%] | 6.14<br>[3.86, 7.71] | 4.79<br>[3.07, 5.86] | 6 / 7 |
| **NCR** | Constant Transmission Acceleration | 20,925<br>[16,343, 25,474] | 22<br>[19.9, 24.0] | 85.7%<br>[28.6%, 71.4%] | 6.00<br>[3.71, 7.57] | 4.71<br>[3.00, 5.79] | 6 / 7 |
| | **Continuous Transmission Acceleration** | **9,517**<br>[6,496, 12,986] | **10**<br>[8.4, 11.4] | **100.0%**<br>[42.9%, 85.7%] | **7.86**<br>[7.57, 8.00] | **6.21**<br>[6.00, 6.50] | **7 / 7** |
| | Incidence Gradient | 8,685<br>[4,423, 12,615] | 7.9<br>[4.3, 10.9] | 71.4%<br>[28.6%, 71.4%] | 5.71<br>[2.29, 8.00] | 4.36<br>[1.79, 6.14] | 5 / 7 |

| Region | Detector | TAM | True alarms | Sens. | Lead (wks) | WP (wks) | Yrs w/ TA |
|---|---|---|---|---|---|---|---|
| | Hydrological Inflection Measure | 7,542<br>[3,656, 11,052] | 6.4<br>[3.1, 9] | 71.4%<br>[28.6%, 71.4%] | 5.29<br>[2.29, 7.71] | 4.12<br>[1.79, 5.90] | 5 / 7 |
| | Critical Transition Indicator | 5,500<br>[2,661, 8,726] | 6.4<br>[3.1, 9.7] | 71.4%<br>[28.6%, 71.4%] | 5.29<br>[2.29, 7.57] | 4.64<br>[2.29, 6.86] | 5 / 7 |
| | Cumulative Sum Control | 10,117<br>[3,676, 17,672] | 7.9<br>[2.4, 13.4] | 42.9%<br>[14.3%, 42.9%] | 2.29<br>[0.57, 4.57] | 2.00<br>[0.57, 3.71] | 3 / 7 |
| | Composite Outbreak Signal | 10,215<br>[4,157, 17,121] | 8.7<br>[3.4, 15.1] | 57.1%<br>[14.3%, 57.1%] | 3.43<br>[1.29, 5.86] | 2.86<br>[1.21, 4.79] | 4 / 7 |
| | Outbreak Threshold | 10,261<br>[4,157, 17,214] | 8.9<br>[3.4, 15.6] | 57.1%<br>[14.3%, 57.1%] | 3.57<br>[1.29, 6.29] | 2.93<br>[1.21, 4.86] | 4 / 7 |
| | Alarm Threshold | 13,002<br>[5,754, 19,933] | 11.4<br>[5.1, 17.7] | 71.4%<br>[28.6%, 71.4%] | 4.57<br>[2.28, 6.86] | 3.79<br>[1.71, 5.50] | 5 / 7 |
| | WHO 75th Percentile Threshold | 14,225<br>[6,572, 21,429] | 12.9<br>[6, 19.1] | 71.4%<br>[28.6%, 71.4%] | 5.29<br>[2.29, 7.71] | 4.05<br>[1.71, 5.81] | 5 / 7 |
| | WHO 90th Percentile Threshold | 13,620<br>[5,983, 20,761] | 12.1<br>[5.3, 18.6] | 71.4%<br>[28.6%, 71.4%] | 5.00<br>[2.29, 7.29] | 3.90<br>[1.71, 5.62] | 5 / 7 |
| **REGION I** | Constant Transmission Acceleration | 8,053<br>[5,773, 10,481] | 19.4<br>[17.1, 22] | 85.7%<br>[28.6%, 85.7%] | 6.00<br>[3.86, 7.43] | 4.71<br>[3.07, 5.71] | 6 / 7 |
| | **Continuous Transmission Acceleration** | **5,017**<br>[3,459, 6,787] | **11.3**<br>[10.7, 11.9] | **100.0%**<br>[42.9%, 85.7%] | **7.86**<br>[7.57, 8.00] | **5.93**<br>[5.79, 6.00] | **7 / 7** |
| | Incidence Gradient | 3,767<br>[1,769, 5,765] | 7.6<br>[4.4, 9.9] | 85.7%<br>[28.6%, 85.7%] | 6.14<br>[3.43, 7.86] | 4.86<br>[3.00, 6.07] | 6 / 7 |
| | Hydrological Inflection Measure | 2,869<br>[816, 5,162] | 4.9<br>[1.4, 8.4] | 42.9%<br>[14.3%, 42.9%] | 3.43<br>[1.14, 6.86] | 2.64<br>[0.86, 5.14] | 3 / 7 |
| | Critical Transition Indicator | 2,994<br>[1,415, 4,506] | 6.9<br>[4.4, 8.7] | 85.7%<br>[28.6%, 85.7%] | 6.57<br>[4.29, 7.86] | 5.00<br>[3.29, 5.93] | 6 / 7 |
| | Cumulative Sum Control | 3,050<br>[0, 6,100] | 4.7<br>[0, 10.7] | 14.3%<br>[0.0%, 14.3%] | 1.14<br>[0.00, 3.43] | 0.86<br>[0.00, 2.57] | 1 / 7 |
| | Composite Outbreak Signal | 3,234<br>[581, 6,272] | 6.4<br>[1.4, 12.4] | 28.6%<br>[0.0%, 28.6%] | 2.29<br>[0.00, 4.57] | 1.79<br>[0.00, 3.71] | 2 / 7 |
| | Outbreak Threshold | 3,234<br>[581, 6,272] | 6.4<br>[1.4, 12.4] | 28.6%<br>[0.0%, 28.6%] | 2.29<br>[0.00, 4.57] | 1.79<br>[0.00, 3.71] | 2 / 7 |
| | Alarm Threshold | 4,162<br>[581, 8,337] | 7.9<br>[2.3, 15.1] | 42.9%<br>[14.3%, 42.9%] | 3.43<br>[1.14, 6.86] | 2.64<br>[0.86, 5.14] | 3 / 7 |
| | WHO 75th Percentile Threshold | 5,104<br>[1,483, 9,261] | 9.4<br>[2.9, 16.1] | 42.9%<br>[14.3%, 42.9%] | 3.43<br>[1.14, 6.86] | 2.64<br>[0.86, 5.14] | 3 / 7 |
| | WHO 90th Percentile Threshold | 4,316<br>[581, 8,389] | 8.1<br>[2.3, 15.3] | 42.9%<br>[14.3%, 42.9%] | 3.43<br>[1.14, 6.86] | 2.64<br>[0.86, 5.14] | 3 / 7 |

| Region | Detector | TAM | True alarms | Sens. | Lead (wks) | WP (wks) | Yrs w/ TA |
|---|---|---|---|---|---|---|---|
| **REGION II** | **Constant Transmission Acceleration** | **7,782** [4,094, 11,718] | **18** [11.0, 23.9] | **71.4%** [28.6%, 71.4%] | **5.71** [3.40, 8.00] | **4.29** [2.55, 6.00] | **5 / 7** |
| | Continuous Transmission Acceleration | 4,416 [2,390, 6,213] | 9.9 [8.6, 10.9] | 71.4% [28.6%, 71.4%] | 5.14 [2.29, 7.43] | 4.00 [1.71, 5.71] | 5 / 7 |
| | Incidence Gradient | 4,047 [1,866, 5,986] | 8.3 [4.1, 11.6] | 57.1% [14.3%, 57.1%] | 4.57 [2.29, 6.86] | 3.46 [1.71, 5.29] | 4 / 7 |
| | Hydrological Inflection Measure | 3,843 [1,519, 6,040] | 8 [3.1, 12.4] | 42.9% [14.3%, 42.9%] | 3.43 [1.14, 6.86] | 2.61 [0.86, 5.18] | 3 / 7 |
| | Critical Transition Indicator | 2,444 [807, 4,072] | 4.9 [2.1, 7.3] | 57.1% [14.3%, 57.1%] | 4.57 [2.29, 6.86] | 3.43 [1.71, 5.14] | 4 / 7 |
| | Cumulative Sum Control | 6,524 [1,967, 10,965] | 12.6 [3.4, 21.0] | 28.6% [0.0%, 28.6%] | 2.29 [0.00, 4.57] | 1.71 [0.00, 3.43] | 2 / 7 |
| | Composite Outbreak Signal | 4,593 [1,639, 8,123] | 9.9 [3.7, 16.7] | 42.9% [14.3%, 42.9%] | 3.43 [1.14, 6.86] | 2.57 [0.86, 5.14] | 3 / 7 |
| | Outbreak Threshold | 5,409 [2,107, 9,081] | 12.9 [4.7, 20.9] | 42.9% [14.3%, 42.9%] | 3.43 [1.14, 6.86] | 2.57 [0.86, 5.14] | 3 / 7 |
| | Alarm Threshold | 6,801 [2,908, 10,351] | 15 [6.6, 22.6] | 42.9% [14.3%, 42.9%] | 3.43 [1.14, 6.86] | 2.57 [0.86, 5.14] | 3 / 7 |
| | WHO 75th Percentile Threshold | 7,245 [3,011, 10,958] | 15.7 [7, 23.9] | 42.9% [14.3%, 42.9%] | 3.43 [1.14, 6.86] | 2.57 [0.86, 5.14] | 3 / 7 |
| | WHO 90th Percentile Threshold | 6,894 [2,908, 10,535] | 15.1 [6.6, 22.9] | 42.9% [14.3%, 42.9%] | 3.43 [1.14, 6.86] | 2.57 [0.86, 5.14] | 3 / 7 |
| **REGION III** | Constant Transmission Acceleration | 23,612 [18,161, 29,327] | 22.3 [20.7, 23.6] | 85.7% [28.6%, 71.8%] | 5.43 [3.14, 7.14] | 4.43 [2.71, 5.57] | 6 / 7 |
| | **Continuous Transmission Acceleration** | **12,224** [7,846, 16,905] | **10.4** [8.7, 11.6] | **85.7%** [28.6%, 71.4%] | **6.86** [4.57, 8.00] | **5.14** [3.43, 6.00] | **6 / 7** |
| | Incidence Gradient | 9,862 [4,677, 15,036] | 7.1 [3.7, 9.9] | 71.4% [28.6%, 71.4%] | 5.29 [2.29, 7.43] | 4.07 [1.71, 5.71] | 5 / 7 |
| | Hydrological Inflection Measure | 8,469 [3,419, 13,555] | 6 [2.6, 9.1] | 57.1% [14.3%, 57.1%] | 4.43 [1.14, 6.86] | 3.31 [0.86, 5.10] | 4 / 7 |
| | Critical Transition Indicator | 8,682 [5,071, 12,154] | 7 [4.4, 8.7] | 71.4% [28.6%, 71.4%] | 4.86 [2.29, 7.00] | 3.86 [1.71, 5.50] | 5 / 7 |
| | Cumulative Sum Control | 12,922 [4,292, 23,021] | 9.7 [3.3, 17.1] | 28.6% [0.0%, 28.6%] | 2.29 [0.00, 4.57] | 1.71 [0.00, 3.43] | 2 / 7 |
| | Composite Outbreak Signal | 11,189 [5,058, 16,715] | 8.6 [3.9, 12.9] | 28.6% [0.0%, 28.6%] | 2.00 [0.00, 4.29] | 1.57 [0.00, 3.29] | 2 / 7 |
| | Outbreak Threshold | 11,330 [5,178, 16,957] | 9 [4, 13.9] | 42.9% [14.3%, 42.9%] | 3.14 [0.86, 6.29] | 2.57 [0.86, 5.14] | 3 / 7 |

| Region | Detector | TAM | True alarms | Sens. | Lead (wks) | WP (wks) | Yrs w/ TA |
|---|---|---|---|---|---|---|---|
| | Alarm Threshold | 14,280<br>[6,351, 21,552] | 11.6<br>[5, 17.6] | 57.1%<br>[14.3%, 57.1%] | 4.14<br>[1.14, 6.86] | 3.21<br>[0.86, 5.14] | 4 / 7 |
| | WHO 75th Percentile Threshold | 15,561<br>[7,044, 23,832] | 12.7<br>[5.4, 19.4] | 57.1%<br>[14.3%, 57.1%] | 4.57<br>[1.14, 6.86] | 3.38<br>[0.86, 5.14] | 4 / 7 |
| | WHO 90th Percentile Threshold | 14,665<br>[6,677, 22,037] | 12<br>[5, 18.3] | 57.1%<br>[14.3%, 57.1%] | 4.57<br>[1.14, 6.86] | 3.38<br>[0.86, 5.14] | 4 / 7 |
| **REGION IV-A** | **Constant Transmission Acceleration** | **24,458**<br>[15,718, 36,037] | **21.7**<br>[19.6, 24] | **85.7%**<br>[28.6%, 85.7%] | **6.00**<br>[3.86, 7.71] | **4.71**<br>[3.07, 5.86] | **6 / 7** |
| | Continuous Transmission Acceleration | 11,149<br>[5,491, 20,485] | 8.7<br>[6.6, 10.4] | 71.4%<br>[28.6%, 71.4%] | 5.71<br>[2.29, 8.00] | 4.29<br>[1.71, 6.00] | 5 / 7 |
| | Incidence Gradient | 10,271<br>[3,641, 19,384] | 6.9<br>[4, 9.3] | 71.4%<br>[28.6%, 71.4%] | 5.71<br>[3.43, 8.00] | 4.36<br>[2.57, 6.14] | 5 / 7 |
| | Hydrological Inflection Measure | 9,255<br>[2,910, 18,387] | 5.9<br>[2.7, 8.6] | 71.4%<br>[28.6%, 71.4%] | 5.57<br>[3.14, 7.86] | 4.32<br>[2.57, 6.07] | 5 / 7 |
| | Critical Transition Indicator | 5,125<br>[1,572, 10,035] | 4.3<br>[1.9, 6.6] | 42.9%<br>[14.3%, 42.9%] | 3.29<br>[1.00, 5.72] | 2.50<br>[0.79, 4.30] | 3 / 7 |
| | Cumulative Sum Control | 12,623<br>[2,082, 28,174] | 6.7<br>[1.1, 13.6] | 28.6%<br>[0.0%, 28.6%] | 1.71<br>[0.00, 4.00] | 1.43<br>[0.00, 3.43] | 2 / 7 |
| | Composite Outbreak Signal | 13,065<br>[2,947, 28,169] | 7.6<br>[2.6, 12.6] | 42.9%<br>[14.3%, 42.9%] | 2.86<br>[0.57, 5.71] | 2.29<br>[0.57, 4.29] | 3 / 7 |
| | Outbreak Threshold | 13,065<br>[2,947, 28,169] | 7.6<br>[2.6, 12.6] | 42.9%<br>[14.3%, 42.9%] | 2.86<br>[0.57, 5.71] | 2.29<br>[0.57, 4.29] | 3 / 7 |
| | Alarm Threshold | 14,298<br>[3,547, 29,142] | 8.9<br>[3.6, 14] | 71.4%<br>[28.6%, 71.4%] | 4.71<br>[2.00, 7.00] | 4.07<br>[1.86, 6.07] | 5 / 7 |
| | WHO 75th Percentile Threshold | 16,019<br>[4,669, 30,926] | 10.7<br>[4.6, 16.9] | 71.4%<br>[28.6%, 71.4%] | 5.57<br>[3.14, 7.86] | 4.25<br>[2.43, 6.00] | 5 / 7 |
| | WHO 90th Percentile Threshold | 15,189<br>[3,877, 30,591] | 9.9<br>[3.7, 16.1] | 71.4%<br>[28.6%, 71.4%] | 5.43<br>[2.85, 7.71] | 4.43<br>[2.29, 6.43] | 5 / 7 |
| **REGION IX** | **Constant Transmission Acceleration** | **7,641**<br>[4,398, 11,390] | **22.3**<br>[18.1, 25.9] | **71.4%**<br>[28.6%, 71.4%] | **5.43**<br>[3.14, 7.71] | **4.14**<br>[2.42, 5.86] | **5 / 7** |
| | Continuous Transmission Acceleration | 3,194<br>[1,464, 5,424] | 7.6<br>[5.7, 9.3] | 57.1%<br>[14.3%, 57.1%] | 4.29<br>[1.14, 6.71] | 3.29<br>[0.86, 5.07] | 4 / 7 |
| | Incidence Gradient | 3,137<br>[1,473, 5,275] | 7.3<br>[4.7, 9.7] | 85.7%<br>[28.6%, 85.7%] | 6.00<br>[4.00, 7.71] | 5.00<br>[3.36, 6.29] | 6 / 7 |
| | Hydrological Inflection Measure | 2,194<br>[378, 4,633] | 4.6<br>[1.6, 7.9] | 57.1%<br>[14.3%, 57.1%] | 4.00<br>[1.42, 6.57] | 3.36<br>[1.42, 5.36] | 4 / 7 |
| | Critical Transition Indicator | 1,910<br>[786, 3,093] | 6.4<br>[3.1, 9.6] | 28.6%<br>[0.0%, 28.6%] | 2.29<br>[0.00, 4.57] | 1.71<br>[0.00, 3.43] | 2 / 7 |

| Region | Detector | TAM | True alarms | Sens. | Lead (wks) | WP (wks) | Yrs w/ TA |
|---|---|---|---|---|---|---|---|
| | Cumulative Sum Control | 2,938<br>[252, 7,828] | 5.1<br>[0.7, 11.9] | 14.3%<br>[0.0%, 14.3%] | 1.14<br>[0.00, 3.43] | 0.86<br>[0.00, 2.57] | 1 / 7 |
| | Composite Outbreak Signal | 2,749<br>[138, 7,673] | 4.6<br>[0.6, 11.1] | 14.3%<br>[0.0%, 14.3%] | 1.14<br>[0.00, 3.43] | 0.86<br>[0.00, 2.57] | 1 / 7 |
| | Outbreak Threshold | 2,811<br>[217, 7,714] | 4.9<br>[0.9, 11.4] | 28.6%<br>[0.0%, 28.6%] | 2.29<br>[0.00, 4.57] | 2.00<br>[0.00, 4.29] | 2 / 7 |
| | Alarm Threshold | 3,503<br>[612, 8,250] | 7.4<br>[2.3, 13.9] | 42.9%<br>[14.3%, 42.9%] | 3.29<br>[1.00, 5.73] | 2.61<br>[0.86, 4.45] | 3 / 7 |
| | WHO 75th Percentile Threshold | 4,090<br>[896, 8,699] | 9.3<br>[3.7, 15.3] | 57.1%<br>[14.3%, 57.1%] | 4.14<br>[1.42, 6.86] | 3.21<br>[1.27, 5.14] | 4 / 7 |
| | WHO 90th Percentile Threshold | 3,594<br>[649, 8,306] | 7.9<br>[2.4, 14.4] | 42.9%<br>[14.3%, 42.9%] | 3.29<br>[1.00, 5.73] | 2.50<br>[0.79, 4.30] | 3 / 7 |
| **REGION V** | **Constant Transmission Acceleration** | **2,446**<br>[1,250, 4,325] | **21.9**<br>[19.1, 24.6] | **85.7%**<br>[28.6%, 71.8%] | **5.71**<br>[3.57, 7.29] | **4.57**<br>[2.93, 5.64] | **6 / 7** |
| | Continuous Transmission Acceleration | 1,306<br>[433, 2,759] | 8.3<br>[6.0, 10.3] | 71.4%<br>[28.6%, 71.4%] | 5.57<br>[3.28, 7.86] | 4.21<br>[2.50, 5.93] | 5 / 7 |
| | Incidence Gradient | 1,262<br>[313, 2,735] | 6.7<br>[3.3, 10.1] | 71.4%<br>[28.6%, 71.4%] | 4.86<br>[2.29, 7.14] | 3.82<br>[2.00, 5.54] | 5 / 7 |
| | Hydrological Inflection Measure | 1,125<br>[136, 2,655] | 5<br>[1.4, 9.1] | 57.1%<br>[14.3%, 57.1%] | 4.00<br>[1.70, 6.29] | 3.39<br>[1.39, 5.64] | 4 / 7 |
| | Critical Transition Indicator | 678<br>[223, 1,269] | 5.7<br>[2.9, 8.4] | 42.9%<br>[14.3%, 42.9%] | 3.14<br>[0.86, 5.71] | 2.43<br>[0.71, 4.29] | 3 / 7 |
| | Cumulative Sum Control | 1,425<br>[0, 3,428] | 5.9<br>[0, 12.4] | 14.3%<br>[0.0%, 14.3%] | 1.14<br>[0.00, 3.43] | 0.86<br>[0.00, 2.57] | 1 / 7 |
| | Composite Outbreak Signal | 1,544<br>[94, 3,735] | 6.9<br>[0.9, 14.6] | 42.9%<br>[14.3%, 42.9%] | 2.86<br>[0.57, 5.71] | 2.29<br>[0.57, 4.29] | 3 / 7 |
| | Outbreak Threshold | 1,562<br>[112, 3,736] | 7.1<br>[1.1, 14.6] | 42.9%<br>[14.3%, 42.9%] | 2.86<br>[0.57, 5.71] | 2.29<br>[0.57, 4.29] | 3 / 7 |
| | Alarm Threshold | 1,661<br>[210, 3,822] | 8.3<br>[2.3, 15.4] | 57.1%<br>[14.3%, 57.1%] | 4.00<br>[1.70, 6.29] | 3.43<br>[1.42, 5.71] | 4 / 7 |
| | WHO 75th Percentile Threshold | 1,694<br>[244, 3,888] | 8.7<br>[2.7, 15.6] | 57.1%<br>[14.3%, 57.1%] | 4.00<br>[1.70, 6.29] | 3.43<br>[1.42, 5.71] | 4 / 7 |
| | WHO 90th Percentile Threshold | 1,671<br>[221, 3,842] | 8.4<br>[2.4, 15.4] | 57.1%<br>[14.3%, 57.1%] | 4.00<br>[1.70, 6.29] | 3.43<br>[1.42, 5.71] | 4 / 7 |
| **REGION VI** | Constant Transmission Acceleration | 11,630<br>[5,362, 19,567] | 18.4<br>[14.3, 22.3] | 85.7%<br>[28.6%, 71.8%] | 5.71<br>[3.71, 7.29] | 4.57<br>[3.07, 5.64] | 6 / 7 |
| | **Continuous Transmission Acceleration** | **9,655**<br>[3,912, 16,657] | **11.3**<br>[10.1, 12] | **100.0%**<br>[42.9%, 85.7%] | **7.57**<br>[7.00, 8.00] | **5.79**<br>[5.50, 6.00] | **7 / 7** |

| Region | Detector | TAM | True alarms | Sens. | Lead (wks) | WP (wks) | Yrs w/ TA |
|---|---|---|---|---|---|---|---|
| | Incidence Gradient | 5,984<br>[1,296, 11,901] | 6.4<br>[3, 10] | 57.1%<br>[14.3%, 57.1%] | 4.43<br>[2.14, 6.86] | 3.36<br>[1.64, 5.14] | 4 / 7 |
| | Hydrological Inflection Measure | 4,129<br>[65, 10,461] | 2<br>[0.3, 4.4] | 28.6%<br>[0.0%, 28.6%] | 2.14<br>[0.00, 4.43] | 1.86<br>[0.00, 4.00] | 2 / 7 |
| | Critical Transition Indicator | 5,749<br>[936, 12,161] | 6.4<br>[3.9, 8.7] | 85.7%<br>[28.6%, 71.8%] | 6.43<br>[4.29, 7.86] | 5.00<br>[3.29, 6.07] | 6 / 7 |
| | Cumulative Sum Control | 5,250<br>[0, 14,494] | 2.9<br>[0, 6.9] | 14.3%<br>[0.0%, 14.3%] | 1.14<br>[0.00, 3.43] | 0.86<br>[0.00, 2.57] | 1 / 7 |
| | Composite Outbreak Signal | 3,615<br>[0, 10,845] | 1.4<br>[0, 4.3] | 14.3%<br>[0.0%, 14.3%] | 1.14<br>[0.00, 3.43] | 0.86<br>[0.00, 2.57] | 1 / 7 |
| | Outbreak Threshold | 3,615<br>[0, 10,845] | 1.4<br>[0, 4.3] | 14.3%<br>[0.0%, 14.3%] | 1.14<br>[0.00, 3.43] | 0.86<br>[0.00, 2.57] | 1 / 7 |
| | Alarm Threshold | 5,923<br>[0, 14,404] | 3.1<br>[0, 6.9] | 14.3%<br>[0.0%, 14.3%] | 1.14<br>[0.00, 3.43] | 0.86<br>[0.00, 2.57] | 1 / 7 |
| | WHO 75th Percentile Threshold | 6,640<br>[65, 15,855] | 3.9<br>[0.3, 8] | 28.6%<br>[0.0%, 28.6%] | 2.14<br>[0.00, 4.43] | 1.86<br>[0.00, 4.00] | 2 / 7 |
| | WHO 90th Percentile Threshold | 5,952<br>[29, 14,433] | 3.3<br>[0.1, 6.9] | 14.3%<br>[0.0%, 14.3%] | 1.14<br>[0.00, 3.43] | 0.86<br>[0.00, 2.57] | 1 / 7 |
| **REGION VII** | **Constant Transmission Acceleration** | **10,337**<br>[5,511, 15,466] | **20.9**<br>[17, 25.1] | **42.9%**<br>[14.3%, 42.9%] | **3.43**<br>[1.14, 6.86] | **2.57**<br>[0.86, 5.14] | **3 / 7** |
| | Continuous Transmission Acceleration | 5,631<br>[2,652, 8,818] | 9.4<br>[6.9, 11.4] | 42.9%<br>[14.3%, 42.9%] | 3.14<br>[0.86, 6.00] | 2.43<br>[0.71, 4.71] | 3 / 7 |
| | Incidence Gradient | 5,288<br>[2,099, 8,424] | 8<br>[3.7, 12.3] | 42.9%<br>[14.3%, 42.9%] | 2.43<br>[0.71, 4.29] | 2.07<br>[0.64, 3.58] | 3 / 7 |
| | Hydrological Inflection Measure | 5,201<br>[2,106, 8,227] | 7.7<br>[3.6, 12.1] | 42.9%<br>[14.3%, 42.9%] | 2.29<br>[0.57, 4.29] | 2.00<br>[0.57, 3.58] | 3 / 7 |
| | Critical Transition Indicator | 3,339<br>[896, 6,753] | 4.7<br>[1.9, 7.4] | 14.3%<br>[0.0%, 14.3%] | 0.86<br>[0.00, 2.57] | 0.71<br>[0.00, 2.14] | 1 / 7 |
| | Cumulative Sum Control | 8,799<br>[2,845, 14,403] | 14.3<br>[5, 25.4] | 28.6%<br>[0.0%, 28.6%] | 2.29<br>[0.00, 5.71] | 1.71<br>[0.00, 4.29] | 2 / 7 |
| | Composite Outbreak Signal | 8,023<br>[3,027, 13,095] | 12.4<br>[5.0, 20.9] | 14.3%<br>[0.0%, 14.3%] | 1.14<br>[0.00, 3.43] | 0.86<br>[0.00, 2.57] | 1 / 7 |
| | Outbreak Threshold | 8,777<br>[3,812, 13,707] | 14.6<br>[6.6, 22.1] | 28.6%<br>[0.0%, 28.6%] | 2.29<br>[0.00, 5.71] | 1.71<br>[0.00, 4.29] | 2 / 7 |
| | Alarm Threshold | 9,758<br>[4,104, 15,221] | 17<br>[7.4, 26.0] | 42.9%<br>[14.3%, 42.9%] | 2.86<br>[0.57, 5.71] | 2.29<br>[0.57, 4.29] | 3 / 7 |
| | WHO 75th Percentile Threshold | 9,912<br>[4,174, 15,384] | 17.9<br>[7.9, 27.9] | 42.9%<br>[14.3%, 42.9%] | 2.86<br>[0.57, 5.71] | 2.29<br>[0.57, 4.29] | 3 / 7 |

| Region | Detector | TAM | True alarms | Sens. | Lead (wks) | WP (wks) | Yrs w/ TA |
|---|---|---|---|---|---|---|---|
| | WHO 90th Percentile Threshold | 9,774 [4,104, 15,253] | 17.1 [7.6, 26.4] | 42.9% [14.3%, 42.9%] | 2.86 [0.57, 5.71] | 2.29 [0.57, 4.29] | 3 / 7 |
| **REGION VIII** | **Constant Transmission Acceleration** | **8,120** [3,316, 13,727] | **16.9** [11, 21.6] | **85.7%** [28.6%, 85.7%] | **5.29** [3.43, 7.00] | **4.36** [2.86, 5.50] | **6 / 7** |
| | Continuous Transmission Acceleration | 4,489 [1,493, 8,059] | 8.3 [6, 10.4] | 71.4% [28.6%, 71.4%] | 5.71 [3.43, 8.00] | 4.43 [2.57, 6.29] | 5 / 7 |
| | Incidence Gradient | 2,622 [476, 5,807] | 4.9 [1.9, 7.9] | 57.1% [14.3%, 57.1%] | 4.14 [1.14, 6.71] | 3.57 [1.14, 6.00] | 4 / 7 |
| | Hydrological Inflection Measure | 1,795 [0, 5,090] | 2.1 [0, 5] | 28.6% [0.0%, 28.6%] | 2.14 [0.00, 4.57] | 1.64 [0.00, 3.43] | 2 / 7 |
| | Critical Transition Indicator | 2,240 [270, 4,607] | 6.4 [1.9, 11.7] | 28.6% [0.0%, 28.6%] | 1.86 [0.00, 4.14] | 1.50 [0.00, 3.21] | 2 / 7 |
| | Cumulative Sum Control | 2,906 [0, 8,281] | 3.4 [0, 8] | 28.6% [0.0%, 28.6%] | 2.00 [0.00, 4.29] | 1.57 [0.00, 3.43] | 2 / 7 |
| | Composite Outbreak Signal | 1,524 [0, 4,573] | 1 [0, 3] | 14.3% [0.0%, 14.3%] | 0.71 [0.00, 2.14] | 0.64 [0.00, 1.93] | 1 / 7 |
| | Outbreak Threshold | 1,702 [0, 4,927] | 1.9 [0, 4.4] | 28.6% [0.0%, 28.6%] | 2.14 [0.00, 4.57] | 1.82 [0.00, 4.00] | 2 / 7 |
| | Alarm Threshold | 2,506 [0, 7,170] | 3 [0, 7] | 28.6% [0.0%, 28.6%] | 2.29 [0.00, 4.57] | 1.79 [0.00, 3.71] | 2 / 7 |
| | WHO 75th Percentile Threshold | 2,837 [62, 7,907] | 3.7 [0.1, 8.4] | 28.6% [0.0%, 28.6%] | 2.29 [0.00, 4.57] | 1.71 [0.00, 3.43] | 2 / 7 |
| | WHO 90th Percentile Threshold | 2,642 [0, 7,508] | 3.3 [0, 7.6] | 28.6% [0.0%, 28.6%] | 2.29 [0.00, 4.57] | 1.71 [0.00, 3.43] | 2 / 7 |
| **REGION X** | Constant Transmission Acceleration | 11,045 [7,041, 14,755] | 23.4 [21.4, 25.4] | 71.4% [28.6%, 71.4%] | 4.57 [2.29, 6.72] | 3.71 [1.93, 5.36] | 5 / 7 |
| | **Continuous Transmission Acceleration** | **4,311** [2,000, 6,535] | **6.6** [4.1, 8.6] | **85.7%** [28.6%, 85.7%] | **6.14** [4.00, 7.57] | **5.14** [3.36, 6.57] | **6 / 7** |
| | Incidence Gradient | 4,397 [1,693, 7,232] | 7.1 [3.4, 10.9] | 71.4% [28.6%, 71.4%] | 5.57 [3.14, 7.86] | 4.64 [2.57, 6.64] | 5 / 7 |
| | Hydrological Inflection Measure | 3,288 [828, 6,088] | 4.4 [1, 8.6] | 42.9% [14.3%, 42.9%] | 2.86 [0.57, 5.71] | 2.29 [0.57, 4.29] | 3 / 7 |
| | Critical Transition Indicator | 3,177 [877, 5,648] | 5.3 [2.1, 8.3] | 42.9% [14.3%, 42.9%] | 3.14 [0.86, 5.72] | 2.50 [0.71, 4.64] | 3 / 7 |
| | Cumulative Sum Control | 6,265 [1,096, 12,412] | 10.6 [2.9, 19.3] | 28.6% [0.0%, 28.6%] | 2.29 [0.00, 4.57] | 1.71 [0.00, 3.43] | 2 / 7 |
| | Composite Outbreak Signal | 2,065 [0, 4,227] | 2.7 [0, 6.1] | 28.6% [0.0%, 28.6%] | 1.86 [0.00, 4.14] | 1.57 [0.00, 3.43] | 2 / 7 |

| Region | Detector | TAM | True alarms | Sens. | Lead (wks) | WP (wks) | Yrs w/ TA |
|---|---|---|---|---|---|---|---|
| | Outbreak Threshold | 2,065<br>[0, 4,227] | 2.7<br>[0, 6.1] | 28.6%<br>[0.0%, 28.6%] | 1.86<br>[0.00, 4.14] | 1.57<br>[0.00, 3.43] | 2 / 7 |
| | Alarm Threshold | 4,039<br>[432, 8,435] | 5.3<br>[0.4, 10.6] | 28.6%<br>[0.0%, 28.6%] | 2.29<br>[0.00, 4.57] | 1.71<br>[0.00, 3.43] | 2 / 7 |
| | WHO 75th Percentile Threshold | 6,521<br>[1,538, 12,341] | 9.6<br>[2.3, 17.9] | 42.9%<br>[14.3%, 42.9%] | 2.86<br>[0.57, 5.71] | 2.29<br>[0.57, 4.29] | 3 / 7 |
| | WHO 90th Percentile Threshold | 4,850<br>[624, 9,534] | 6.6<br>[0.7, 13.4] | 28.6%<br>[0.0%, 28.6%] | 2.29<br>[0.00, 4.57] | 1.71<br>[0.00, 3.43] | 2 / 7 |
| **REGION XI** | **Constant Transmission Acceleration** | **7,585**<br>[3,543, 11,934] | **21.1**<br>[13.1, 27.3] | **57.1%**<br>[14.3%, 57.1%] | **4.29**<br>[1.14, 7.14] | **3.29**<br>[0.86, 5.57] | **4 / 7** |
| | Continuous Transmission Acceleration | 2,572<br>[675, 5,113] | 5.1<br>[2.1, 8.1] | 57.1%<br>[14.3%, 57.1%] | 3.57<br>[1.14, 6.00] | 3.14<br>[1.14, 5.21] | 4 / 7 |
| | Incidence Gradient | 3,129<br>[1,025, 5,671] | 7.1<br>[3.4, 10.4] | 57.1%<br>[14.3%, 57.1%] | 3.57<br>[1.14, 6.00] | 3.00<br>[1.14, 4.86] | 4 / 7 |
| | Hydrological Inflection Measure | 2,717<br>[557, 5,364] | 5.3<br>[2, 8.6] | 42.9%<br>[14.3%, 42.9%] | 2.71<br>[0.57, 5.29] | 2.29<br>[0.57, 4.29] | 3 / 7 |
| | Critical Transition Indicator | 1,164<br>[307, 2,208] | 3.3<br>[1.1, 5.7] | 28.6%<br>[0.0%, 28.6%] | 1.57<br>[0.00, 3.57] | 1.36<br>[0.00, 3.07] | 2 / 7 |
| | Cumulative Sum Control | 6,050<br>[1,103, 11,759] | 13.7<br>[4.1, 23.1] | 57.1%<br>[14.3%, 57.1%] | 4.57<br>[2.26, 6.86] | 3.64<br>[1.70, 5.79] | 4 / 7 |
| | Composite Outbreak Signal | 4,405<br>[448, 8,388] | 8<br>[1.7, 15.9] | 42.9%<br>[14.3%, 42.9%] | 3.14<br>[0.86, 5.71] | 2.71<br>[0.71, 5.14] | 3 / 7 |
| | Outbreak Threshold | 4,405<br>[448, 8,388] | 8<br>[1.7, 15.9] | 42.9%<br>[14.3%, 42.9%] | 3.14<br>[0.86, 5.71] | 2.71<br>[0.71, 5.14] | 3 / 7 |
| | Alarm Threshold | 5,467<br>[820, 10,718] | 11.1<br>[3.4, 20.0] | 42.9%<br>[14.3%, 42.9%] | 3.43<br>[1.14, 5.71] | 2.57<br>[0.86, 4.29] | 3 / 7 |
| | WHO 75th Percentile Threshold | 6,121<br>[1,275, 11,469] | 13.7<br>[5.0, 22.3] | 57.1%<br>[14.3%, 57.1%] | 4.43<br>[1.14, 7.57] | 3.36<br>[0.86, 5.79] | 4 / 7 |
| | WHO 90th Percentile Threshold | 5,805<br>[851, 11,172] | 12.4<br>[3.9, 21.1] | 57.1%<br>[14.3%, 57.1%] | 4.43<br>[1.14, 7.57] | 3.36<br>[0.86, 5.79] | 4 / 7 |
| **REGION XII** | **Constant Transmission Acceleration** | **8,650**<br>[4,739, 12,642] | **23.1**<br>[13.7, 29.9] | **71.4%**<br>[28.6%, 71.4%] | **5.43**<br>[2.29, 8.00] | **4.14**<br>[1.71, 6.00] | **5 / 7** |
| | Continuous Transmission Acceleration | 2,592<br>[1,116, 4,193] | 6<br>[3, 8.9] | 71.4%<br>[28.6%, 71.4%] | 4.71<br>[2.42, 7.00] | 3.79<br>[2.07, 5.50] | 5 / 7 |
| | Incidence Gradient | 3,140<br>[1,027, 5,631] | 7<br>[3.6, 10.6] | 71.4%<br>[28.6%, 71.4%] | 5.43<br>[2.29, 7.71] | 4.33<br>[1.76, 6.10] | 5 / 7 |
| | Hydrological Inflection Measure | 2,363<br>[393, 4,955] | 4.3<br>[1.1, 8.6] | 42.9%<br>[14.3%, 42.9%] | 3.14<br>[0.86, 5.71] | 2.39<br>[0.71, 4.25] | 3 / 7 |

| Region | Detector | TAM | True alarms | Sens. | Lead (wks) | WP (wks) | Yrs w/ TA |
|---|---|---|---|---|---|---|---|
| | Critical Transition Indicator | 2,167<br>[654, 3,780] | 5.7<br>[1.3, 10.1] | 57.1%<br>[14.3%, 57.1%] | 4.43<br>[1.14, 7.43] | 3.43<br>[0.86, 5.71] | 4 / 7 |
| | Cumulative Sum Control | 4,691<br>[329, 10,198] | 9.6<br>[1.1, 20.0] | 28.6%<br>[0.0%, 28.6%] | 2.29<br>[0.00, 4.60] | 1.71<br>[0.00, 3.45] | 2 / 7 |
| | Composite Outbreak Signal | 1,927<br>[294, 4,001] | 4.1<br>[1, 7.9] | 14.3%<br>[0.0%, 14.3%] | 1.14<br>[0.00, 3.43] | 0.93<br>[0.00, 2.79] | 1 / 7 |
| | Outbreak Threshold | 2,534<br>[882, 4,273] | 6.7<br>[2.6, 11.9] | 14.3%<br>[0.0%, 14.3%] | 1.14<br>[0.00, 3.43] | 0.93<br>[0.00, 2.79] | 1 / 7 |
| | Alarm Threshold | 4,233<br>[1,213, 7,638] | 9.9<br>[4.3, 16.1] | 28.6%<br>[0.0%, 28.6%] | 1.86<br>[0.00, 4.14] | 1.57<br>[0.00, 3.29] | 2 / 7 |
| | WHO 75th Percentile Threshold | 5,196<br>[1,493, 9,234] | 12<br>[5.3, 19.0] | 42.9%<br>[14.3%, 42.9%] | 3.14<br>[0.86, 5.71] | 2.39<br>[0.71, 4.25] | 3 / 7 |
| | WHO 90th Percentile Threshold | 4,401<br>[1,281, 7,715] | 10.4<br>[4.7, 16.6] | 28.6%<br>[0.0%, 28.6%] | 2.29<br>[0.00, 4.57] | 1.67<br>[0.00, 3.38] | 2 / 7 |
| **REGION XIII** | **Constant Transmission Acceleration** | **4,377**<br>[1,922, 7,349] | **15**<br>[7.7, 22] | **57.1%**<br>[14.3%, 57.1%] | **4.57**<br>[1.14, 6.86] | **3.43**<br>[0.86, 5.14] | **4 / 7** |
| | Continuous Transmission Acceleration | 1,432<br>[173, 2,812] | 3.1<br>[0.6, 6] | 14.3%<br>[0.0%, 14.3%] | 0.57<br>[0.00, 1.71] | 0.57<br>[0.00, 1.71] | 1 / 7 |
| | Incidence Gradient | 2,195<br>[841, 3,885] | 6.3<br>[3.1, 9.9] | 42.9%<br>[14.3%, 42.9%] | 3.14<br>[0.86, 6.00] | 2.43<br>[0.71, 4.71] | 3 / 7 |
| | Hydrological Inflection Measure | 2,089<br>[551, 3,885] | 5<br>[1.4, 9.3] | 42.9%<br>[14.3%, 42.9%] | 3.14<br>[0.86, 6.00] | 2.71<br>[0.71, 5.29] | 3 / 7 |
| | Critical Transition Indicator | 983<br>[201, 1,945] | 3.7<br>[1.6, 5.7] | 0.0%<br>[0.0%, 0.0%] | 0.00<br>[0.00, 0.00] | 0.00<br>[0.00, 0.00] | 0 / 7 |
| | Cumulative Sum Control | 3,945<br>[999, 7,512] | 12.7<br>[3.7, 22.7] | 42.9%<br>[14.3%, 42.9%] | 3.43<br>[1.14, 6.86] | 2.71<br>[0.86, 5.29] | 3 / 7 |
| | Composite Outbreak Signal | 2,940<br>[629, 6,103] | 6.7<br>[1.1, 13.9] | 28.6%<br>[0.0%, 28.6%] | 2.29<br>[0.00, 4.57] | 1.75<br>[0.00, 3.57] | 2 / 7 |
| | Outbreak Threshold | 2,992<br>[680, 6,103] | 7.1<br>[1.6, 13.9] | 28.6%<br>[0.0%, 28.6%] | 2.29<br>[0.00, 4.57] | 1.75<br>[0.00, 3.57] | 2 / 7 |
| | Alarm Threshold | 3,696<br>[989, 7,091] | 10.4<br>[3.6, 18.4] | 42.9%<br>[14.3%, 42.9%] | 3.43<br>[1.14, 6.86] | 2.86<br>[0.86, 5.43] | 3 / 7 |
| | WHO 75th Percentile Threshold | 3,905<br>[1,100, 7,308] | 11.7<br>[4.0, 20.6] | 42.9%<br>[14.3%, 42.9%] | 3.43<br>[1.14, 6.86] | 2.86<br>[0.86, 5.43] | 3 / 7 |
| | WHO 90th Percentile Threshold | 3,827<br>[1,046, 7,260] | 11.1<br>[3.8, 19.3] | 42.9%<br>[14.3%, 42.9%] | 3.43<br>[1.14, 6.86] | 2.86<br>[0.86, 5.43] | 3 / 7 |

Within each region the 11 detectors are listed in the paradigm-grouped order used throughout the paper. Each metric cell gives the point estimate (top) above its 95% percentile bootstrap confidence interval (bottom), computed from B = 1,000 year-cluster resamples within the region. The four-tier consensus winner is shown in bold so it can be read against the other 10 methods. Confidence intervals are not given for the years-with-true-alarm column, which is a discrete year-count fraction. Year inclusion is n = 7 evaluable years per region (2016 to 2024 excluding 2020, 2021 and 2025); regions are retained where the series has at least 12 annual peak cases. NCR contains Quezon City and is therefore not independent of the city-scale derivation cohort; the remaining 16 regions form the out-of-sample validation set. TAM, true-alarm magnitude; Sens., sensitivity; Lead, mean weeks from the first true actionable alarm to the peak; WP, warning persistence; Yrs w/ TA, evaluable years with at least one actionable-window true alarm over the region total.

Supplementary Table 11 | Regional headline performance of the 11 outbreak-detection methods, pooled across 17 Philippine regions.

| | *Epidemic burden & alarm accuracy* | | | *Early-warning timeliness* | | *False Alarms* | |
|---|---|---|---|---|---|---|---|
| **Detector** | **TAM (cases/yr)** | **True alarms ($n\ yr^{-1}$)** | **Sensitivity** | **Mean lead time (wks)** | **Warning persistence (wks)** | **False alarms ($n\ yr^{-1}$)** | **Regions w/ true alarm** |
| **Constant Transmission Acceleration** | **10,338** [7,525, 13,530] | **20.7** [19.4, 21.8] | **76.3%** [69.7%, 82.1%] | **5.40** [5.03, 5.73] | **4.23** [3.91, 4.49] | **6.9** [5.7, 8.1] | **17 / 17** |
| **Continuous Transmission Acceleration** | **5,168** [3,627, 6,840] | **8.4** [7.3, 9.4] | **75.6%** [64.7%, 85.7%] | **5.58** [4.67, 6.40] | **4.37** [3.67, 4.98] | **2.0** [1.6, 2.4] | **17 / 17** |
| **Incidence Gradient** | **4,543** [3,385, 5,850] | **7.2** [6.8, 7.6] | **68.8%** [62.0%, 75.1%] | **5.04** [4.47, 5.56] | **4.01** [3.60, 4.42] | **3.1** [2.6, 3.6] | **17 / 17** |
| **WHO 75th Percentile Threshold** | **7,102** [5,251, 9,254] | **11.6** [9.7, 13.7] | **51.0%** [43.4%, 58.8%] | **3.85** [3.29, 4.44] | **2.99** [2.56, 3.43] | **8.7** [7.2, 10.4] | **17 / 17** |
| **WHO 90th Percentile Threshold** | **6,592** [4,888, 8,561] | **10.6** [8.7, 12.6] | **47.6%** [38.7%, 56.0%] | **3.62** [3.01, 4.24] | **2.81** [2.30, 3.30] | **7.5** [6.0, 9.1] | **17 / 17** |
| **Cumulative Sum Control** | **6,024** [4,601, 7,679] | **9.6** [7.5, 11.9] | **33.3%** [26.6%, 40.6%] | **2.53** [1.94, 3.12] | **1.97** [1.53, 2.45] | **7.2** [5.8, 8.6] | **17 / 17** |
| **Alarm Threshold** | **6,316** [4,578, 8,325] | **10.0** [8.0, 11.9] | **46.8%** [38.1%, 55.5%] | **3.44** [2.87, 4.02] | **2.73** [2.25, 3.23] | **6.7** [5.4, 8.1] | **17 / 17** |
| **Critical Transition Indicator** | **3,058** [2,152, 4,092] | **5.5** [5.0, 6.0] | **51.0%** [39.5%, 61.9%] | **3.76** [2.81, 4.58] | **2.97** [2.28, 3.68] | **3.7** [3.0, 4.5] | **16 / 17** |
| **Hydrological Inflection Measure** | **3,850** [2,849, 5,046] | **5.4** [4.5, 6.3] | **50.1%** [42.9%, 58.3%] | **3.66** [3.08, 4.25] | **2.91** [2.47, 3.37] | **2.4** [2.0, 2.9] | **17 / 17** |
| **Outbreak Threshold** | **5,002** [3,449, 6,789] | **7.5** [5.8, 9.1] | **38.4%** [30.3%, 47.3%] | **2.78** [2.25, 3.37] | **2.25** [1.81, 2.75] | **4.6** [3.6, 5.7] | **17 / 17** |
| **Composite Outbreak Signal** | **4,838** [3,335, 6,566] | **6.8** [5.3, 8.2] | **35.0%** [26.1%, 45.1%] | **2.47** [1.88, 3.12] | **1.99** [1.50, 2.56] | **2.3** [1.8, 2.9] | **17 / 17** |

Each cell gives the cross-region point estimate above its 95% percentile bootstrap confidence interval, from B = 1,000 region-cluster resamples of the 17 regions. Detector order follows the paradigm-grouped convention. Year inclusion is n = 7 evaluable years per region (2016 to 2024 excluding 2020, 2021 and 2025). Regions-with-true-alarm counts the regions in which the detector fired at least one actionable-window true alarm in at least one year, over 17.

Supplementary Table 12 | Detector-paired Wilcoxon signed-rank tests by operational metric, regional analysis.

| **Metric** | **Comparison** | ***n* regions** | **Median paired Δ (A − B)** | ***V*** | ***P*** | ***P* pairwise** | **Sig.** |
|---|---|---|---|---|---|---|---|
| ***Epidemic burden & alarm accuracy*** | | | | | | | |
| **True-alarm magnitude (cases)** | Constant TA vs Outbreak Threshold | 17 | 4,819 | 153.0 | <0.001 | **<0.001*** | ✓ |

| Metric | Comparison | *n* regions | Median paired Δ (A − B) | *V* | *P* | *P* pairwise | Sig. |
|---|---|---|---|---|---|---|---|
| | Continuous TA vs Outbreak Threshold | 17 | 57.571 | 75.0 | 0.962 | 0.962 | |
| | Constant TA vs Continuous TA | 17 | 3,632 | 153.0 | <0.001 | **<0.001*** | ✓ |
| **Number of true alarms** | Constant TA vs Outbreak Threshold | 17 | 13.286 | 153.0 | <0.001 | **<0.001*** | ✓ |
| | Continuous TA vs Outbreak Threshold | 17 | 1.143 | 94.0 | 0.421 | 0.421 | |
| | Constant TA vs Continuous TA | 17 | 11.857 | 153.0 | <0.001 | **<0.001*** | ✓ |
| **Sensitivity** | Constant TA vs Outbreak Threshold | 17 | 0.429 | 136.0 | <0.001 | **<0.001*** | ✓ |
| | Continuous TA vs Outbreak Threshold | 17 | 0.333 | 151.0 | <0.001 | **<0.001*** | ✓ |
| | Constant TA vs Continuous TA | 17 | 0.000 | 25.5 | 0.528 | 0.528 | |
| ***Early-warning timeliness*** | | | | | | | |
| **Mean lead time (wks)** | Constant TA vs Outbreak Threshold | 17 | 2.714 | 152.0 | <0.001 | **<0.001*** | ✓ |
| | Continuous TA vs Outbreak Threshold | 17 | 2.857 | 148.0 | <0.001 | **<0.001*** | ✓ |
| | Constant TA vs Continuous TA | 17 | -0.167 | 59.0 | 0.421 | 0.421 | |
| **Warning persistence (wks)** | Constant TA vs Outbreak Threshold | 17 | 2.143 | 152.0 | <0.001 | **<0.001*** | ✓ |
| | Continuous TA vs Outbreak Threshold | 17 | 2.000 | 148.0 | <0.001 | **<0.001*** | ✓ |
| | Constant TA vs Continuous TA | 17 | -0.071 | 59.0 | 0.421 | 0.421 | |

Comparisons are paired by region (n = 17). Each row reports the median paired difference (A minus B) with its 95% percentile interval from B = 1,000 year-cluster bootstrap resamples within each region, the Wilcoxon signed-rank statistic V, the raw two-sided P, and the Bonferroni-adjusted pairwise P across the three contrasts within the metric (corrected alpha = 0.0167). A pair is flagged significant where the interval on the median excludes zero. The Wilcoxon P values are a sensitivity diagnostic and the bootstrap intervals are the primary effect-size statement.

Supplementary Table 13 | All-pairs head-to-head consensus check with test statistics, regional analysis.

| | | | ***Strict cross-region Bonferroni (k = 51)*** | | | ***Within-region only Bonferroni (k = 3)*** | | | ***Aggregate consensus statistics*** | | | |
|---|---|---|---|---|---|---|---|---|---|---|---|---|
| **Region** | **Consensus winner** | ***Pr* (Sig.)** | **Const TA vs OT** | **Cont TA vs OT** | **Const TA vs Cont TA** | **Const TA vs OT** | **Cont TA vs OT** | **Const TA vs Cont TA** | ***n* dom.** | **Weakest-link *P* (k=51)** | ***n* sig wins** | ***n* sig losses** |
| BARMM | Constant Transmission Acceleration | 0.731 (***) | CI excl. 0 | CI excl. 0 | CI excl. 0 | CI excl. 0 | CI excl. 0 | CI excl. 0 | 1 | CI excl. 0 | 2 | 0 |
| CAR | Contested | 0.465 (*) | CI excl. 0 | CI excl. 0 | CI excl. 0 | CI excl. 0 | CI excl. 0 | CI excl. 0 | 1 | 0.045 | 1 | 1 |

| | | | | | | | | | | | | |
|---|---|---|---|---|---|---|---|---|---|---|---|---|
| MIMAROPA | Continuous Transmission Acceleration | 0.742 (***) | CI excl. 0 | CI excl. 0 | CI excl. 0 | CI excl. 0 | CI excl. 0 | CI excl. 0 | 1 | CI excl. 0 | 2 | 0 |
| NCR | Continuous Transmission Acceleration | 0.592 (***) | CI excl. 0 | CI excl. 0 | CI excl. 0 | CI excl. 0 | CI excl. 0 | CI excl. 0 | 1 | CI excl. 0 | 2 | 0 |
| REGION I | Continuous Transmission Acceleration | 0.947 (***) | CI excl. 0 | CI excl. 0 | CI excl. 0 | CI excl. 0 | CI excl. 0 | CI excl. 0 | 1 | CI excl. 0 | 2 | 0 |
| REGION II | Constant Transmission Acceleration | 0.962 (***) | CI excl. 0 | CI excl. 0 | CI excl. 0 | CI excl. 0 | CI excl. 0 | CI excl. 0 | 1 | CI excl. 0 | 2 | 0 |
| REGION III | Continuous Transmission Acceleration (partial) | 0.524 (ns) | CI excl. 0 | CI excl. 0 | ns (CI incl. 0) | CI excl. 0 | CI excl. 0 | ns (CI incl. 0) | 0 | — | 1 | 0 |
| REGION IV-A | Constant Transmission Acceleration | 0.830 (***) | CI excl. 0 | CI excl. 0 | CI excl. 0 | CI excl. 0 | CI excl. 0 | CI excl. 0 | 1 | CI excl. 0 | 2 | 0 |
| REGION IX | Constant Transmission Acceleration | 0.810 (***) | CI excl. 0 | CI excl. 0 | CI excl. 0 | CI excl. 0 | CI excl. 0 | CI excl. 0 | 1 | CI excl. 0 | 2 | 0 |
| REGION V | Constant Transmission Acceleration | 0.856 (***) | CI excl. 0 | CI excl. 0 | CI excl. 0 | CI excl. 0 | CI excl. 0 | CI excl. 0 | 1 | CI excl. 0 | 2 | 0 |
| REGION VI | Continuous Transmission Acceleration | 1.000 (***) | ns (CI incl. 0) | CI excl. 0 | CI excl. 0 | ns (CI incl. 0) | CI excl. 0 | CI excl. 0 | 1 | CI excl. 0 | 2 | 0 |
| REGION VII | Constant Transmission Acceleration | 0.734 (***) | CI excl. 0 | CI excl. 0 | CI excl. 0 | CI excl. 0 | CI excl. 0 | CI excl. 0 | 1 | CI excl. 0 | 2 | 0 |
| REGION VIII | Constant Transmission Acceleration | 0.654 (***) | CI excl. 0 | CI excl. 0 | CI excl. 0 | CI excl. 0 | CI excl. 0 | CI excl. 0 | 1 | CI excl. 0 | 2 | 0 |
| REGION X | Continuous Transmission Acceleration | 0.969 (***) | CI excl. 0 | CI excl. 0 | CI excl. 0 | CI excl. 0 | CI excl. 0 | CI excl. 0 | 1 | CI excl. 0 | 2 | 0 |
| REGION XI | Constant Transmission Acceleration | 0.727 (***) | CI excl. 0 | CI excl. 0 | CI excl. 0 | CI excl. 0 | CI excl. 0 | CI excl. 0 | 1 | CI excl. 0 | 2 | 0 |
| REGION XII | Constant Transmission Acceleration | 0.768 (***) | CI excl. 0 | CI excl. 0 | CI excl. 0 | CI excl. 0 | CI excl. 0 | CI excl. 0 | 1 | CI excl. 0 | 2 | 0 |

| | | | | | | | | | | | | |
|---|---|---|---|---|---|---|---|---|---|---|---|---|
| REGION XIII | Constant Transmission Acceleration | 1.000 (***) | CI excl. 0 | ns (CI incl. 0) | CI excl. 0 | CI excl. 0 | ns (CI incl. 0) | CI excl. 0 | 1 | CI excl. 0 | 2 | 0 |

One row per region (n = 17, alphabetical). Cells where the 95% year-cluster bootstrap interval on the pair difference excludes zero read “CI excl. 0”; cells whose interval includes zero read “ns (CI incl. 0)”. The strict cross-region Bonferroni (k = 51, corrected alpha approximately 9.8 x 10^-4) is the primary correction; the within-region Bonferroni (k = 3) is a sensitivity check. No numeric bootstrap P values are reported, because they describe the precision of the bootstrap distribution rather than evidence against a null. The aggregate columns (n dominated, weakest-link, significant wins and losses) reconstruct the four consensus tiers.

Supplementary Table 14 | Per-detector cross-region dominance probability distribution.

| **Detector** | ***n* regions** | **Median *Pr*** | **IQR [Q1, Q3]** | **Min** | **Max** | **Strong + partial winner regions** | **Decisive (Pr ≥ 0.75)** | **Above chance (Pr > 1/3)** |
|---|---|---|---|---|---|---|---|---|
| **Constant Transmission Acceleration** | 17 | **0.727** | [0.408, 0.810] | 0.000 | 1.000 | 10 / 17 | 6 / 17 | 13 / 17 |
| **Continuous Transmission Acceleration** | 17 | **0.273** | [0.179, 0.592] | 0.000 | 1.000 | 6 / 17 | 3 / 17 | 8 / 17 |
| **Outbreak Threshold** | 17 | **0.000** | [0.000, 0.005] | 0.000 | 0.087 | 0 / 17 | 0 / 17 | 0 / 17 |

One row per target detector. Each row summarises the bootstrap dominance probability Pr across the 17 regions (the 51 points in Fig. 5c are 17 regions × 3 detectors), giving the median, the interquartile range, the minimum and the maximum. Strong-plus-partial winner regions count the regions where the consensus winner equals this detector at the strong or partial tier. The decisive count is regions with Pr ≥ 0.75; the above-chance count is regions with Pr > 1/3.

Supplementary Table 15 | Per-country operational performance of the 11 outbreak-detection methods across eight dengue-endemic countries.

| **Region** | **Detector** | **TAM** | **True alarms** | **Sens.** | **Lead (wks)** | **WP (wks)** | **Yrs w/ TA** |
|---|---|---|---|---|---|---|---|
| **BRAZIL** | **Constant Transmission Acceleration** | **1,022,892**<br>[383,745, 1,696,594] | **16.5**<br>[8.8, 22.0] | **83.3%**<br>[33.3%, 83.3%] | **6.67**<br>[4.00, 8.00] | **5.00**<br>[3.00, 6.00] | **5 / 6** |
| | Continuous Transmission Acceleration | 668,300<br>[239,815, 1,116,390] | 8.2<br>[4.5, 11.0] | 83.3%<br>[33.3%, 83.3%] | 6.67<br>[4.00, 8.00] | 5.00<br>[3.00, 6.00] | 5 / 6 |
| | Incidence Gradient | 631,196<br>[239,767, 1,018,686] | 8.5<br>[5.0, 10.8] | 83.3%<br>[33.3%, 83.3%] | 6.33<br>[3.67, 8.00] | 4.92<br>[2.83, 6.17] | 5 / 6 |
| | Hydrological Inflection Measure | 545,890<br>[154,632, 957,116] | 4.7<br>[1.3, 8.0] | 50.0%<br>[16.7%, 50.0%] | 3.83<br>[1.17, 6.50] | 2.92<br>[0.92, 4.92] | 3 / 6 |
| | Critical Transition Indicator | 254,298<br>[79,763, 481,020] | 7.0<br>[2.5, 11.5] | 66.7%<br>[16.7%, 66.7%] | 5.33<br>[2.67, 8.00] | 4.33<br>[2.00, 6.67] | 4 / 6 |
| | Cumulative Sum Control | 701,710<br>[146,758, 1,431,760] | 6.2<br>[1.3, 12.2] | 16.7%<br>[0.0%, 16.7%] | 1.33<br>[0.00, 4.00] | 1.00<br>[0.00, 3.00] | 1 / 6 |
| | Composite Outbreak Signal | 294,338<br>[0, 589,499] | 2.7<br>[0.0, 5.3] | 16.7%<br>[0.0%, 16.7%] | 1.33<br>[0.00, 4.00] | 1.00<br>[0.00, 3.00] | 1 / 6 |

| Region | Detector | TAM | True alarms | Sens. | Lead (wks) | WP (wks) | Yrs w/ TA |
|---|---|---|---|---|---|---|---|
| | Outbreak Threshold | 294,338 [0, 589,499] | 2.7 [0.0, 5.3] | 16.7% [0.0%, 16.7%] | 1.33 [0.00, 4.00] | 1.00 [0.00, 3.00] | 1 / 6 |
| | Alarm Threshold | 741,370 [185,282, 1,298,586] | 6.7 [1.7, 12.2] | 50.0% [16.7%, 50.0%] | 3.33 [0.67, 6.00] | 2.67 [0.67, 4.67] | 3 / 6 |
| | WHO 75th Percentile Threshold | 855,049 [232,719, 1,542,494] | 7.7 [2.2, 13.3] | 50.0% [16.7%, 50.0%] | 3.83 [1.17, 6.50] | 2.92 [0.92, 4.92] | 3 / 6 |
| | WHO 90th Percentile Threshold | 841,074 [218,743, 1,528,518] | 7.5 [2.0, 13.2] | 50.0% [16.7%, 50.0%] | 3.67 [1.00, 6.33] | 2.83 [0.83, 4.83] | 3 / 6 |
| **COLOMBIA** | Constant Transmission Acceleration | 31,581 [6,167, 63,078] | 16.6 [4.2, 28.2] | 40.0% [0.0%, 40.0%] | 3.20 [0.00, 6.40] | 2.40 [0.00, 4.80] | 2 / 5 |
| | Continuous Transmission Acceleration | 944 [0, 2,072] | 0.8 [0.0, 1.8] | 0.0% [0.0%, 0.0%] | 0.00 [0.00, 0.00] | 0.00 [0.00, 0.00] | 0 / 5 |
| | Incidence Gradient | 18,690 [5,289, 32,122] | 10.2 [4.0, 15.6] | 60.0% [20.0%, 60.0%] | 4.40 [1.20, 7.60] | 3.40 [1.00, 5.80] | 3 / 5 |
| | Hydrological Inflection Measure | 16,387 [3,540, 31,064] | 7.8 [2.4, 13.0] | 60.0% [20.0%, 60.0%] | 4.40 [1.20, 7.60] | 3.40 [1.00, 5.80] | 3 / 5 |
| | Critical Transition Indicator | 3,442 [456, 6,954] | 3.2 [0.6, 5.8] | 20.0% [0.0%, 20.0%] | 1.40 [0.00, 4.20] | 1.10 [0.00, 3.30] | 1 / 5 |
| | Cumulative Sum Control | 31,770 [2,246, 67,188] | 15.4 [1.4, 29.4] | 60.0% [20.0%, 60.0%] | 4.20 [1.00, 7.40] | 3.30 [0.90, 5.70] | 3 / 5 |
| | Composite Outbreak Signal | 21,444 [1,485, 55,939] | 8.4 [1.0, 19.8] | 40.0% [0.0%, 40.0%] | 2.60 [0.00, 5.80] | 2.20 [0.00, 4.60] | 2 / 5 |
| | **Outbreak Threshold** | **25,689** [1,970, 59,698] | **11.4** [1.2, 22.6] | **60.0%** [20.0%, 60.0%] | **4.20** [1.00, 7.40] | **3.40** [1.00, 5.80] | **3 / 5** |
| | Alarm Threshold | 28,918 [3,236, 63,346] | 13.6 [2.2, 25.0] | 60.0% [20.0%, 60.0%] | 4.80 [1.60, 8.00] | 3.60 [1.20, 6.00] | 3 / 5 |
| | WHO 75th Percentile Threshold | 30,405 [3,853, 64,215] | 14.8 [2.8, 26.4] | 60.0% [20.0%, 60.0%] | 4.80 [1.60, 8.00] | 3.60 [1.20, 6.00] | 3 / 5 |
| | WHO 90th Percentile Threshold | 29,390 [3,458, 63,596] | 14.0 [2.4, 25.6] | 60.0% [20.0%, 60.0%] | 4.80 [1.60, 8.00] | 3.60 [1.20, 6.00] | 3 / 5 |
| **MEXICO** | Constant Transmission Acceleration | 104,059 [57,787, 151,696] | 17.3 [15.7, 19.2] | 83.3% [33.3%, 83.3%] | 6.67 [4.00, 8.00] | 5.00 [3.00, 6.00] | 5 / 6 |
| | **Continuous Transmission Acceleration** | **69,176** [29,648, 110,548] | **8.8** [7.5, 10.2] | **100.0%** [49.6%, 83.3%] | **6.67** [5.50, 7.67] | **5.33** [4.75, 5.83] | **6 / 6** |
| | Incidence Gradient | 58,311 [15,362, 103,328] | 7.2 [4.0, 9.7] | 83.3% [33.3%, 83.3%] | 6.67 [4.00, 8.00] | 5.15 [3.04, 6.39] | 5 / 6 |
| | Hydrological Inflection Measure | 46,057 [0, 93,794] | 3.5 [0.0, 7.2] | 33.3% [0.0%, 33.3%] | 2.67 [0.00, 5.33] | 2.00 [0.00, 4.00] | 2 / 6 |
| | Critical Transition Indicator | 19,694 [6,269, 33,473] | 2.8 [1.7, 3.7] | 66.7% [16.7%, 66.7%] | 4.33 [1.83, 7.00] | 3.67 [1.58, 5.92] | 4 / 6 |
| | Cumulative Sum Control | 63,960 [0, 129,035] | 5.0 [0.0, 10.3] | 33.3% [0.0%, 33.3%] | 2.67 [0.00, 5.33] | 2.00 [0.00, 4.00] | 2 / 6 |

| Region | Detector | TAM | True alarms | Sens. | Lead (wks) | WP (wks) | Yrs w/ TA |
|---|---|---|---|---|---|---|---|
| | Composite Outbreak Signal | 34,014<br>[0, 96,885] | 3.2<br>[0.0, 7.8] | 16.7%<br>[0.0%, 16.7%] | 1.33<br>[0.00, 4.00] | 1.00<br>[0.00, 3.00] | 1 / 6 |
| | Outbreak Threshold | 34,014<br>[0, 96,885] | 3.2<br>[0.0, 7.8] | 16.7%<br>[0.0%, 16.7%] | 1.33<br>[0.00, 4.00] | 1.00<br>[0.00, 3.00] | 1 / 6 |
| | Alarm Threshold | 63,140<br>[2,579, 126,279] | 5.5<br>[0.8, 10.2] | 33.3%<br>[0.0%, 33.3%] | 2.67<br>[0.00, 5.33] | 2.00<br>[0.00, 4.00] | 2 / 6 |
| | WHO 75th Percentile Threshold | 67,246<br>[3,286, 131,615] | 6.0<br>[1.0, 11.2] | 33.3%<br>[0.0%, 33.3%] | 2.67<br>[0.00, 5.33] | 2.00<br>[0.00, 4.00] | 2 / 6 |
| | WHO 90th Percentile Threshold | 63,846<br>[3,286, 126,716] | 5.7<br>[1.0, 10.3] | 33.3%<br>[0.0%, 33.3%] | 2.67<br>[0.00, 5.33] | 2.00<br>[0.00, 4.00] | 2 / 6 |
| **PERU** | **Constant Transmission Acceleration** | **54,435**<br>[8,572, 120,873] | **15.8**<br>[5.6, 26.2] | **66.7%**<br>[16.7%, 66.7%] | **5.33**<br>[1.33, 8.00] | **4.00**<br>[1.00, 6.00] | **4 / 6** |
| | Continuous Transmission Acceleration | 35,948<br>[4,875, 79,439] | 7.0<br>[3.3, 10.2] | 50.0%<br>[16.7%, 50.0%] | 3.83<br>[1.17, 6.50] | 2.92<br>[0.92, 4.92] | 3 / 6 |
| | Incidence Gradient | 33,172<br>[5,361, 71,805] | 6.8<br>[2.3, 10.3] | 50.0%<br>[16.7%, 50.0%] | 4.00<br>[1.33, 6.67] | 3.00<br>[1.00, 5.00] | 3 / 6 |
| | Hydrological Inflection Measure | 33,562<br>[5,892, 72,336] | 6.7<br>[2.3, 10.7] | 50.0%<br>[16.7%, 50.0%] | 4.00<br>[1.33, 6.67] | 3.00<br>[1.00, 5.00] | 3 / 6 |
| | Critical Transition Indicator | 25,879<br>[2,098, 54,857] | 7.0<br>[3.2, 10.8] | 33.3%<br>[0.0%, 33.3%] | 2.17<br>[0.00, 4.83] | 1.75<br>[0.00, 3.75] | 2 / 6 |
| | Cumulative Sum Control | 42,113<br>[1,426, 109,370] | 8.3<br>[1.8, 15.3] | 33.3%<br>[0.0%, 33.3%] | 2.67<br>[0.00, 5.33] | 2.00<br>[0.00, 4.00] | 2 / 6 |
| | Composite Outbreak Signal | 48,162<br>[4,361, 115,384] | 9.0<br>[3.3, 14.7] | 50.0%<br>[16.7%, 50.0%] | 4.00<br>[1.33, 6.67] | 3.00<br>[1.00, 5.00] | 3 / 6 |
| | Outbreak Threshold | 48,331<br>[4,535, 115,384] | 9.3<br>[3.8, 14.7] | 50.0%<br>[16.7%, 50.0%] | 4.00<br>[1.33, 6.67] | 3.00<br>[1.00, 5.00] | 3 / 6 |
| | Alarm Threshold | 50,751<br>[6,078, 117,710] | 10.7<br>[4.3, 16.3] | 50.0%<br>[16.7%, 50.0%] | 4.00<br>[1.33, 6.67] | 3.00<br>[1.00, 5.00] | 3 / 6 |
| | WHO 75th Percentile Threshold | 52,334<br>[7,615, 119,248] | 11.8<br>[4.5, 18.3] | 66.7%<br>[16.7%, 66.7%] | 5.00<br>[1.33, 7.67] | 4.00<br>[1.00, 6.00] | 4 / 6 |
| | WHO 90th Percentile Threshold | 50,941<br>[6,247, 117,880] | 11.0<br>[4.5, 17.0] | 66.7%<br>[16.7%, 66.7%] | 5.00<br>[1.33, 7.67] | 4.00<br>[1.00, 6.00] | 4 / 6 |
| **PHILIPPINES** | Constant Transmission Acceleration | 149,248<br>[83,559, 218,461] | 21.7<br>[19.7, 23.7] | 66.7%<br>[16.7%, 66.7%] | 3.83<br>[1.33, 6.17] | 3.25<br>[1.33, 5.08] | 4 / 6 |
| | **Continuous Transmission Acceleration** | **79,697**<br>[46,627, 122,048] | **10.7**<br>[10.2, 11.3] | **83.3%**<br>[33.3%, 83.3%] | **6.33**<br>[3.66, 7.83] | **4.83**<br>[2.83, 5.92] | **5 / 6** |
| | Incidence Gradient | 53,360<br>[20,120, 85,838] | 7.2<br>[3.5, 10.3] | 66.7%<br>[16.7%, 66.7%] | 5.17<br>[2.33, 7.83] | 3.92<br>[1.83, 5.92] | 4 / 6 |
| | Hydrological Inflection Measure | 43,614<br>[10,512, 82,879] | 5.0<br>[1.5, 8.8] | 50.0%<br>[16.7%, 50.0%] | 4.00<br>[1.33, 6.67] | 3.00<br>[1.00, 5.00] | 3 / 6 |
| | Critical Transition Indicator | 43,821<br>[16,895, 78,016] | 7.0<br>[3.8, 10.0] | 83.3%<br>[33.3%, 83.3%] | 5.83<br>[3.33, 7.67] | 4.67<br>[2.67, 6.00] | 5 / 6 |

| Region | Detector | TAM | True alarms | Sens. | Lead (wks) | WP (wks) | Yrs w/ TA |
|---|---|---|---|---|---|---|---|
| | Cumulative Sum Control | 84,558<br>[13,470, 179,462] | 8.2<br>[1.8, 15.7] | 33.3%<br>[0.0%, 33.3%] | 2.67<br>[0.00, 5.33] | 2.00<br>[0.00, 4.00] | 2 / 6 |
| | Composite Outbreak Signal | 68,814<br>[7,037, 156,598] | 6.2<br>[1.2, 12.0] | 33.3%<br>[0.0%, 33.3%] | 2.33<br>[0.00, 5.01] | 1.83<br>[0.00, 3.84] | 2 / 6 |
| | Outbreak Threshold | 70,574<br>[7,037, 161,878] | 6.5<br>[1.2, 12.7] | 33.3%<br>[0.0%, 33.3%] | 2.67<br>[0.00, 5.33] | 2.00<br>[0.00, 4.00] | 2 / 6 |
| | Alarm Threshold | 85,910<br>[11,814, 181,125] | 8.5<br>[1.7, 16.2] | 50.0%<br>[16.7%, 50.0%] | 3.67<br>[1.00, 6.67] | 2.92<br>[0.92, 5.00] | 3 / 6 |
| | WHO 75th Percentile Threshold | 98,914<br>[13,470, 202,146] | 10.3<br>[1.8, 19.3] | 50.0%<br>[16.7%, 50.0%] | 4.00<br>[1.33, 6.67] | 3.00<br>[1.00, 5.00] | 3 / 6 |
| | WHO 90th Percentile Threshold | 90,660<br>[11,814, 184,800] | 9.3<br>[1.7, 17.0] | 50.0%<br>[16.7%, 50.0%] | 4.00<br>[1.33, 6.67] | 3.00<br>[1.00, 5.00] | 3 / 6 |
| **SINGAPORE** | **Constant Transmission Acceleration** | **7,413**<br>[2,123, 12,828] | **19.4**<br>[10.3, 27.9] | **71.4%**<br>[28.6%, 71.4%] | **5.14**<br>[2.29, 7.43] | **4.00**<br>[2.00, 5.71] | **5 / 7** |
| | Continuous Transmission Acceleration | 3,265<br>[701, 6,079] | 6.4<br>[3.4, 9.1] | 57.1%<br>[14.3%, 57.1%] | 3.57<br>[1.14, 5.86] | 2.93<br>[0.86, 4.64] | 4 / 7 |
| | Incidence Gradient | 3,595<br>[1,094, 6,351] | 9.3<br>[5.7, 12.0] | 57.1%<br>[14.3%, 57.1%] | 4.14<br>[1.14, 6.71] | 3.17<br>[0.86, 4.95] | 4 / 7 |
| | Hydrological Inflection Measure | 3,159<br>[422, 6,223] | 5.6<br>[1.6, 9.6] | 57.1%<br>[14.3%, 57.1%] | 3.71<br>[1.14, 6.00] | 3.00<br>[0.86, 4.71] | 4 / 7 |
| | Critical Transition Indicator | 972<br>[322, 1,681] | 4.6<br>[1.4, 8.0] | 42.9%<br>[0.0%, 42.9%] | 3.00<br>[0.00, 5.71] | 2.43<br>[0.00, 4.50] | 3 / 7 |
| | Cumulative Sum Control | 5,622<br>[683, 11,590] | 9.3<br>[1.7, 16.9] | 28.6%<br>[0.0%, 28.6%] | 2.14<br>[0.00, 4.57] | 1.64<br>[0.00, 3.43] | 2 / 7 |
| | Composite Outbreak Signal | 3,547<br>[332, 6,849] | 6.3<br>[0.9, 13.0] | 28.6%<br>[0.0%, 28.6%] | 2.29<br>[0.00, 4.57] | 1.71<br>[0.00, 3.43] | 2 / 7 |
| | Outbreak Threshold | 3,547<br>[332, 6,849] | 6.3<br>[0.9, 13.0] | 28.6%<br>[0.0%, 28.6%] | 2.29<br>[0.00, 4.57] | 1.71<br>[0.00, 3.43] | 2 / 7 |
| | Alarm Threshold | 4,696<br>[595, 9,307] | 8.6<br>[2.0, 16.0] | 57.1%<br>[14.3%, 57.1%] | 3.86<br>[1.14, 6.14] | 3.07<br>[0.86, 4.86] | 4 / 7 |
| | WHO 75th Percentile Threshold | 5,667<br>[602, 11,717] | 9.7<br>[2.0, 17.7] | 57.1%<br>[14.3%, 57.1%] | 3.86<br>[1.14, 6.14] | 3.07<br>[0.86, 4.86] | 4 / 7 |
| | WHO 90th Percentile Threshold | 4,644<br>[543, 9,255] | 8.4<br>[2.0, 16.0] | 57.1%<br>[14.3%, 57.1%] | 3.86<br>[1.14, 6.14] | 3.07<br>[0.86, 4.86] | 4 / 7 |
| **SRI LANKA** | **Constant Transmission Acceleration** | **43,229**<br>[17,810, 72,632] | **21.1**<br>[13.4, 29.1] | **57.1%**<br>[14.3%, 57.1%] | **4.57**<br>[2.26, 6.86] | **3.43**<br>[1.69, 5.14] | **4 / 7** |
| | Continuous Transmission Acceleration | 21,504<br>[7,183, 38,672] | 6.7<br>[4.1, 9.3] | 14.3%<br>[0.0%, 14.3%] | 0.71<br>[0.00, 2.14] | 0.64<br>[0.00, 1.93] | 1 / 7 |
| | Incidence Gradient | 22,209<br>[5,104, 42,686] | 6.9<br>[2.9, 11.0] | 42.9%<br>[14.3%, 42.9%] | 3.43<br>[1.14, 5.71] | 2.75<br>[0.82, 4.93] | 3 / 7 |
| | Hydrological Inflection Measure | 21,286<br>[3,836, 42,217] | 6.1<br>[1.7, 10.6] | 42.9%<br>[14.3%, 42.9%] | 3.43<br>[1.14, 5.71] | 2.79<br>[0.86, 4.93] | 3 / 7 |

| Region | Detector | TAM | True alarms | Sens. | Lead (wks) | WP (wks) | Yrs w/ TA |
|---|---|---|---|---|---|---|---|
| | Critical Transition Indicator | 10,260<br>[4,099, 16,911] | 3.7<br>[2.1, 5.4] | 14.3%<br>[0.0%, 14.3%] | 0.57<br>[0.00, 1.71] | 0.57<br>[0.00, 1.71] | 1 / 7 |
| | Cumulative Sum Control | 16,092<br>[2,958, 31,862] | 9.1<br>[2.0, 18.0] | 28.6%<br>[0.0%, 28.6%] | 2.29<br>[0.00, 4.57] | 1.79<br>[0.00, 3.64] | 2 / 7 |
| | Composite Outbreak Signal | 27,546<br>[3,054, 60,869] | 6.9<br>[1.6, 13.0] | 28.6%<br>[0.0%, 28.6%] | 2.29<br>[0.00, 4.57] | 1.71<br>[0.00, 3.43] | 2 / 7 |
| | Outbreak Threshold | 27,834<br>[3,340, 60,876] | 7.0<br>[1.7, 13.1] | 28.6%<br>[0.0%, 28.6%] | 2.29<br>[0.00, 4.57] | 1.71<br>[0.00, 3.43] | 2 / 7 |
| | Alarm Threshold | 32,373<br>[7,840, 64,424] | 9.3<br>[3.7, 15.6] | 28.6%<br>[0.0%, 28.6%] | 2.29<br>[0.00, 4.57] | 1.71<br>[0.00, 3.43] | 2 / 7 |
| | WHO 75th Percentile Threshold | 37,015<br>[10,853, 68,871] | 12.9<br>[5.4, 20.6] | 42.9%<br>[14.3%, 42.9%] | 3.43<br>[1.14, 5.71] | 2.79<br>[0.86, 4.93] | 3 / 7 |
| | WHO 90th Percentile Threshold | 32,599<br>[8,253, 64,650] | 9.4<br>[3.9, 15.7] | 28.6%<br>[0.0%, 28.6%] | 2.29<br>[0.00, 4.57] | 1.71<br>[0.00, 3.43] | 2 / 7 |
| **TAIWAN** | Constant Transmission Acceleration | 4,383<br>[110, 12,818] | 6.4<br>[3.2, 10.6] | 40.0%<br>[0.0%, 40.0%] | 2.80<br>[0.00, 6.00] | 2.20<br>[0.00, 4.60] | 2 / 5 |
| | **Continuous Transmission Acceleration** | **3,425**<br>[121, 9,923] | **8.4**<br>[5.0, 11.8] | **60.0%**<br>[20.0%, 60.0%] | **4.40**<br>[1.20, 7.60] | **3.40**<br>[1.00, 5.80] | **3 / 5** |
| | Incidence Gradient | 2,995<br>[47, 8,811] | 6.0<br>[2.6, 9.0] | 40.0%<br>[0.0%, 40.0%] | 3.20<br>[0.00, 6.40] | 2.40<br>[0.00, 4.80] | 2 / 5 |
| | Hydrological Inflection Measure | 2,994<br>[45, 8,810] | 5.4<br>[2.2, 8.6] | 40.0%<br>[0.0%, 40.0%] | 3.20<br>[0.00, 6.40] | 2.40<br>[0.00, 4.80] | 2 / 5 |
| | Critical Transition Indicator | 60<br>[6, 128] | 2.8<br>[0.2, 5.8] | 20.0%<br>[0.0%, 20.0%] | 0.80<br>[0.00, 2.40] | 0.80<br>[0.00, 2.40] | 1 / 5 |
| | Cumulative Sum Control | 4,244<br>[1, 12,721] | 3.2<br>[0.2, 8.8] | 20.0%<br>[0.0%, 20.0%] | 1.60<br>[0.00, 4.80] | 1.20<br>[0.00, 3.60] | 1 / 5 |
| | Composite Outbreak Signal | 4,266<br>[0, 12,743] | 4.4<br>[0.0, 10.0] | 40.0%<br>[0.0%, 40.0%] | 2.40<br>[0.00, 5.60] | 2.00<br>[0.00, 4.40] | 2 / 5 |
| | Outbreak Threshold | 4,288<br>[26, 12,761] | 7.8<br>[2.0, 13.0] | 40.0%<br>[0.0%, 40.0%] | 2.40<br>[0.00, 5.60] | 2.00<br>[0.00, 4.40] | 2 / 5 |
| | Alarm Threshold | 4,324<br>[63, 12,774] | 8.0<br>[2.2, 13.0] | 40.0%<br>[0.0%, 40.0%] | 2.40<br>[0.00, 5.60] | 2.00<br>[0.00, 4.40] | 2 / 5 |
| | WHO 75th Percentile Threshold | 4,348<br>[81, 12,797] | 9.4<br>[3.0, 14.8] | 40.0%<br>[0.0%, 40.0%] | 3.20<br>[0.00, 6.40] | 2.40<br>[0.00, 4.80] | 2 / 5 |
| | WHO 90th Percentile Threshold | 4,325<br>[64, 12,774] | 8.8<br>[3.0, 13.8] | 40.0%<br>[0.0%, 40.0%] | 3.20<br>[0.00, 6.40] | 2.40<br>[0.00, 4.80] | 2 / 5 |

Within each country the 11 detectors are listed in the paradigm-grouped order used throughout the paper. Each metric cell gives the point estimate (top) above its 95% percentile bootstrap confidence interval (bottom), from B = 1,000 year-cluster resamples within the country. The consensus winner is in bold; Mexico's bold row is annotated "(partial)" because its leader reaches partial rather than strong consensus. Year inclusion is 2016 to 2024 excluding 2020, 2021 and 2025, giving 5 to 7 evaluable years per country. Abbreviations follow Supplementary Table 10.

Supplementary Table 16 | Cross-country headline performance of the 11 outbreak-detection methods, pooled across eight dengue-endemic countries.

| Detector | *Epidemic burden & alarm accuracy* | | | *Early-warning timeliness* | | *False Alarms* | |
|---|---|---|---|---|---|---|---|
| | TAM (cases/yr) | True alarms (n $yr^{-1}$) | Sensitivity | Mean lead time (wks) | Warning persistence (wks) | False alarms (n $yr^{-1}$) | Countries w/ true alarm |
| Constant Transmission Acceleration | 177,155 [31,144, 426,557] | 16.9 [13.2, 19.5] | 63.6% [52.1%, 74.3%] | 4.78 [3.81, 5.80] | 3.66 [2.98, 4.31] | 10.9 [7.8, 14.8] | 8 / 8 |
| Continuous Transmission Acceleration | 110,283 [15,213, 276,832] | 7.1 [5.1, 8.8] | 56.0% [31.6%, 77.2%] | 4.02 [2.29, 5.60] | 3.13 [1.78, 4.34] | 3.6 [2.4, 5.1] | 7 / 8 |
| WHO 75th Percentile Threshold | 143,872 [26,016, 354,991] | 10.3 [8.5, 12.2] | 50.0% [42.8%, 57.1%] | 3.85 [3.35, 4.34] | 2.97 [2.56, 3.38] | 9.0 [6.6, 11.8] | 8 / 8 |
| WHO 90th Percentile Threshold | 139,685 [24,379, 346,358] | 9.3 [7.8, 11.0] | 48.2% [39.2%, 56.5%] | 3.68 [3.07, 4.32] | 2.83 [2.33, 3.31] | 7.7 [5.3, 10.4] | 8 / 8 |
| Incidence Gradient | 102,941 [17,826, 258,728] | 7.8 [6.9, 8.7] | 60.4% [49.5%, 72.1%] | 4.67 [3.88, 5.54] | 3.59 [2.94, 4.19] | 4.2 [3.4, 5.0] | 8 / 8 |
| Alarm Threshold | 126,435 [23,581, 307,932] | 8.8 [7.3, 10.6] | 46.1% [38.4%, 53.0%] | 3.38 [2.82, 3.93] | 2.62 [2.22, 3.06] | 7.6 [5.3, 10.2] | 8 / 8 |
| Outbreak Threshold | 63,577 [18,880, 133,239] | 6.8 [4.9, 8.6] | 34.2% [25.3%, 44.4%] | 2.56 [1.92, 3.29] | 1.98 [1.46, 2.54] | 5.8 [4.0, 7.9] | 8 / 8 |
| Cumulative Sum Control | 118,758 [20,227, 292,244] | 8.1 [5.9, 10.7] | 31.7% [24.2%, 41.1%] | 2.45 [1.92, 3.03] | 1.87 [1.45, 2.36] | 6.5 [4.1, 9.3] | 8 / 8 |
| Hydrological Inflection Measure | 89,119 [14,679, 222,166] | 5.6 [4.7, 6.5] | 47.9% [42.1%, 53.8%] | 3.66 [3.29, 3.98] | 2.81 [2.51, 3.07] | 3.1 [2.2, 4.1] | 8 / 8 |
| Critical Transition Indicator | 44,803 [7,662, 106,668] | 4.8 [3.5, 6.0] | 43.4% [27.1%, 61.1%] | 2.93 [1.61, 4.22] | 2.41 [1.35, 3.45] | 4.9 [3.8, 6.2] | 8 / 8 |
| Composite Outbreak Signal | 62,766 [18,925, 131,662] | 5.9 [4.4, 7.4] | 31.7% [24.4%, 39.2%] | 2.32 [1.83, 2.92] | 1.81 [1.42, 2.23] | 5.0 [3.2, 7.0] | 8 / 8 |

Each cell gives the cross-country point estimate above its 95% percentile bootstrap confidence interval, from B = 1,000 country-cluster resamples of the eight countries. Year inclusion is 2016 to 2024 excluding 2020, 2021 and 2025, with 5 to 7 evaluable years per country (Colombia and Taiwan, 5; Brazil, Mexico, Peru and the Philippines, 6; Singapore and Sri Lanka, 7). Countries-with-true-alarm counts the countries in which the detector fired at least one actionable-window true alarm in at least one year, over eight.

Supplementary Table 17 | Detector-paired Wilcoxon signed-rank tests by operational metric, country analysis.

| Metric | Comparison | *n* countries | Median paired Δ (A − B) | *V* | *P* | *P* pairwise | Sig. |
|---|---|---|---|---|---|---|---|
| ***Epidemic burden & alarm accuracy*** | | | | | | | |
| **True-alarm magnitude (cases)** | Constant TA vs Outbreak Threshold | 8 | 10,749 | 36.0 | 0.014 | **0.014*** | ✓ |
| | Continuous TA vs Outbreak Threshold | 8 | -572 | 19.0 | 0.944 | 0.944 | |
| | Constant TA vs Continuous TA | 8 | 26,180 | 36.0 | 0.014 | **0.014*** | ✓ |
| **Number of true alarms** | Constant TA vs Outbreak Threshold | 8 | 13.488 | 35.0 | 0.021 | **0.021*** | ✓ |
| | Continuous TA vs Outbreak Threshold | 8 | 0.371 | 22.0 | 0.624 | 0.624 | |
| | Constant TA vs Continuous TA | 8 | 9.917 | 35.0 | 0.021 | **0.021*** | ✓ |

| Metric | Comparison | *n* countries | Median paired Δ (A − B) | *V* | *P* | *P* pairwise | Sig. |
|---|---|---|---|---|---|---|---|
| **Sensitivity** | Constant TA vs Outbreak Threshold | 8 | 0.310 | 26.0 | 0.051 | 0.051 | |
| | Continuous TA vs Outbreak Threshold | 8 | 0.243 | 22.0 | 0.205 | 0.205 | |
| | Constant TA vs Continuous TA | 8 | 0.071 | 16.5 | 0.735 | 0.735 | |
| ***Early-warning timeliness*** | | | | | | | |
| **Mean lead time (wks)** | Constant TA vs Outbreak Threshold | 8 | 1.810 | 34.0 | 0.030 | **0.030*** | ✓ |
| | Continuous TA vs Outbreak Threshold | 8 | 1.643 | 26.0 | 0.293 | 0.293 | |
| | Constant TA vs Continuous TA | 8 | 0.750 | 14.0 | 0.529 | 0.529 | |
| **Warning persistence (wks)** | Constant TA vs Outbreak Threshold | 8 | 1.482 | 33.5 | 0.035 | **0.035*** | ✓ |
| | Continuous TA vs Outbreak Threshold | 8 | 1.307 | 27.0 | 0.234 | 0.234 | |
| | Constant TA vs Continuous TA | 8 | 0.536 | 18.0 | 0.554 | 0.554 | |

Comparisons are paired by country (n = 8). Each row reports the median paired difference (A minus B), the Wilcoxon signed-rank statistic V, the raw two-sided P, and the Bonferroni-adjusted pairwise P across the three within-country contrasts (corrected alpha = 0.0167). An asterisk marks adjusted P < 0.05. The within-country correction is a sensitivity check; the primary correction at the country scale is the strict cross-country Bonferroni (k = 24, corrected alpha approximately 2.1 x 10^-3) applied at the consensus check. Six of 15 contrasts retain significance.

Supplementary Table 18 | All-pairs head-to-head consensus check with test statistics, country analysis.

| | | | ***Strict cross-region Bonferroni (k = 24)*** | | | ***Within-region only Bonferroni (k = 3)*** | | | ***Aggregate consensus statistics*** | | | |
|---|---|---|---|---|---|---|---|---|---|---|---|---|
| **Region** | **Consensus winner** | ***Pr* (Sig.)** | **Const TA vs OT** | **Cont TA vs OT** | **Const TA vs Cont TA** | **Const TA vs OT** | **Cont TA vs OT** | **Const TA vs Cont TA** | ***n* dom.** | **Weakest-link *P* (k=21)** | ***n* sig wins** | ***n* sig losses** |
| **BRAZIL** | Constant Transmission Acceleration | 1.000 (***) | CI excl. 0 | ns (CI incl. 0) | CI excl. 0 | CI excl. 0 | ns (CI incl. 0) | CI excl. 0 | 1 | CI excl. 0 | 2 | 0 |
| **COLOMBIA** | Outbreak Threshold | 0.657 (***) | CI excl. 0 | CI excl. 0 | CI excl. 0 | CI excl. 0 | CI excl. 0 | CI excl. 0 | 1 | CI excl. 0 | 2 | 0 |
| **MEXICO** | Continuous Transmission Acceleration *(partial)* | *0.502 (ns)* | CI excl. 0 | CI excl. 0 | ns (CI incl. 0) | CI excl. 0 | CI excl. 0 | ns (CI incl. 0) | 0 | 1.000 | 1 | 0 |

| | | | | | | | | | | | | |
|---|---|---|---|---|---|---|---|---|---|---|---|---|
| **PERU** | Constant Transmission Acceleration | 0.938 (***) | CI excl. 0 | CI excl. 0 | CI excl. 0 | CI excl. 0 | CI excl. 0 | CI excl. 0 | 1 | CI excl. 0 | 2 | 0 |
| **PHILIPPINES** | Continuous Transmission Acceleration | 0.867 (***) | CI excl. 0 | CI excl. 0 | CI excl. 0 | CI excl. 0 | CI excl. 0 | CI excl. 0 | 1 | CI excl. 0 | 2 | 0 |
| **SINGAPORE** | Constant Transmission Acceleration | 0.979 (***) | CI excl. 0 | CI excl. 0 | CI excl. 0 | CI excl. 0 | CI excl. 0 | CI excl. 0 | 1 | CI excl. 0 | 2 | 0 |
| **SRI LANKA** | Constant Transmission Acceleration | 1.000 (***) | CI excl. 0 | ns (CI incl. 0) | CI excl. 0 | CI excl. 0 | ns (CI incl. 0) | CI excl. 0 | 1 | CI excl. 0 | 2 | 0 |
| **TAIWAN** | Continuous Transmission Acceleration | 0.700 (***) | CI excl. 0 | CI excl. 0 | CI excl. 0 | CI excl. 0 | CI excl. 0 | CI excl. 0 | 1 | CI excl. 0 | 2 | 0 |

One row per country (n = 8, alphabetical). Cells where the 95% year-cluster bootstrap interval on the pair difference excludes zero read “CI excl. 0”; cells whose interval includes zero read “ns (CI incl. 0)”. The strict cross-country Bonferroni (k = 24, corrected alpha approximately 2.1 x 10^-3) is the primary correction; the within-country Bonferroni (k = 3) is a sensitivity check. The aggregate columns (n dominated, weakest-link, significant wins and losses) reconstruct the four consensus tiers, defined as strong (two significant wins, no losses), partial (one win, no losses), lead-only (no wins, no losses, Pr ≥ 0.50) and contested otherwise.

Supplementary Table 19 | Per-detector country dominance probability distribution.

| Detector | *n* countries | Median *Pr* | IQR [Q1, Q3] | Min | Max | Consensus-winner countries | Decisive (Pr ≥ 0.75) | Above chance (Pr > 1/3) |
|---|---|---|---|---|---|---|---|---|
| Constant Transmission Acceleration | 8 | 0.718 | [0.301, 0.984] | 0.128 | 1.000 | 4 / 8 | 4 / 8 | 6 / 8 |
| Continuous Transmission Acceleration | 8 | 0.015 | [0.004, 0.551] | 0.000 | 0.867 | 3 / 8 | 1 / 8 | 3 / 8 |
| Outbreak Threshold | 8 | 0.003 | [0.000, 0.067] | 0.000 | 0.657 | 1 / 8 | 0 / 8 | 1 / 8 |

One row per target detector. Each row summarises the bootstrap dominance probability Pr across the eight countries (the 24 points in Fig. 6b are 8 countries × 3 detectors), giving the median, the interquartile range (Type-7 quantiles), the minimum and the maximum. Consensus-winner countries count the countries where the consensus winner equals this detector, including Mexico’s partial-tier continuous transmission acceleration. The decisive count is countries with Pr ≥ 0.75; the above-chance count is countries with Pr > 1/3.

Supplementary Table 20 | Per-method estimates of the hysteresis thresholds.

| Derivation method | Activation threshold | Deactivation threshold | Notes |
|---|---|---|---|
| M1: Baseline-ratio asymmetric quantiles (90th / 50th percentiles) | 0.925 | 0.516 | Empirical Quezon City series; donor-year pool excluding 2020, 2021, 2025 |

| **Derivation method** | **Activation threshold** | **Deactivation threshold** | **Notes** |
|---|---|---|---|
| M2: ROC operating points (sensitivities 95% / 99%) | n.a. | n.a. | No convergent operating point under the primary specification (B = 2,000 bootstrap) |
| M3: $ARL_0$ = 12-week calibration (baseline-only fraction 30%) | 1.000 | n.a. | Achieved $ARL_0$ = 13.96 wks at activation threshold of 1.00 (closest to 12-wk target); secondary target $ARL_0$ = 26 wks reached at activation threshold of 1.14 (achieved 25.77 wks). M3 calibrates activation threshold only. |
| M4: Leave-one-year-out cross-validation; asymmetric utility favouring early triggering | 1.850 | 1.050 | Utility grid maximised across thirteen evaluable years (Quezon City series) |
| M5: Negative-binomial theoretical thresholds (±1σ) | 1.328 | 0.672 | Parametric (NB fitted to Quezon City weekly counts) |
| M6: Parametric bootstrap quantiles (90th / 25th percentiles) | 1.481 | 0.791 | B = 2,000 bootstrap |
| Range (min, max) across reportable methods | (0.925, 1.850) | (0.516, 1.050) | activation threshold: n = 5 reportable methods (M1, M3, M4, M5, M6); deactivation threshold: n = 4 reportable methods (M1, M4, M5, M6) |
| Median (adopted headline) | 1.33 | 0.73 | Raw median 1.328 and 0.731; rounded to 1.33 and 0.73 for the operational specification. Adoption rule: raw_median. |

Six derivation methods were applied to the constant transmission acceleration activation and deactivation thresholds on the Quezon City weekly series with 2020, 2021 and 2025 excluded (13 evaluable seasons). Two methods do not return a reportable pair: M2 (ROC operating points) finds no operating point under the primary specification, and M3 (ARL0 calibration) returns an activation threshold only. The headline thresholds are the raw median across the reportable values (n = 5 for activation, n = 4 for deactivation), giving an activation threshold of 1.33 and a deactivation threshold of 0.73. Across the reportable methods the activation estimates span 0.925 to 1.850 and the deactivation estimates span 0.516 to 1.050; this range is the empirical sensitivity envelope for the threshold choice. The raw-median rule was pre-specified for robustness across this range.
